# Polygenic predisposition modifies the associations of fish oil supplementation with circulating omega-3 fatty acids: a cross-sectional gene-diet interaction study in UK Biobank

**DOI:** 10.64898/2026.04.29.26352078

**Authors:** Huifang Xu, Ge Yu, Yueqi Lu, Harriett Fuller, Suhang Song, Ye Shen, Charleston W. K. Chiang, Burcu F. Darst, Kaixiong Ye

## Abstract

**Background:** Several genetic variants have been identified to modify the effects of fish oil supplementation (FOS) on increasing circulating omega-3 fatty acids, but it remains unexplored whether polygenic predisposition to low circulating omega-3 fatty acids modifies these effects.

**Objective:** To test if polygenic scores (PGS) for circulating omega-3 fatty acids modify the associations of FOS with corresponding circulating concentrations.

**Methods:** We developed PGS models for absolute circulating concentrations of total omega-3 fatty acids (Omega-3), docosahexaenoic acid (DHA), and their relative percentages in total fatty acids (Omega-3% and DHA%), using a multi-ethnic genome-wide association study (N=136,016). PGS models were validated in 437,803 UK Biobank participants of European (EUR), Central/South Asian (CSA), African, and East Asian genetic ancestries. Linear models tested PGS-by-FOS interactions on corresponding observed circulating concentrations. Discovery analysis was performed separately in 237,380 EUR participants and each non-EUR group. Replication analyses were performed using oily fish intake and in another 178,935 EUR participants.

**Results:** In EUR participants, PGS explained 5.3-11.1% of the phenotypic variance, and significant PGS-by-FOS interactions were detected across all four circulating omega-3 traits. Among participants in the bottom 5% of the PGS distribution, FOS was significantly associated with a 0.40 SD (95% CI: 0.39–0.44) increase in Omega-3. This association effect was 11.1% larger than the population average (β = 0.36; 95% CI: 0.35–0.37; P_Int_ = 0.016) and 42.8% larger than that in participants in the top 5% of the PGS distribution (β = 0.28 SD; 95% CI: 0.25–0.32; P_Int_ = 4.03X10^-10^). These interaction patterns were consistently observed in CSA ancestry and confirmed in replication and sensitivity analyses.

**Conclusions:** PGS modify the associations of FOS with circulating omega-3 fatty acids in EUR and CSA populations, with larger FOS effects in participants with lower PGS. These findings support the development of genome-informed precision nutrition.

## Introduction

Fish oil supplementation (FOS), a rich source of long-chain omega-3 polyunsaturated fatty acids (PUFAs), has raised global attention due to its potential benefits across a spectrum of health conditions, including cardiovascular events [1-4], breast cancer [5], colorectal cancer [6], Alzheimer’s disease and dementia [7, 8]. The beneficial effects of fish oil are mediated by elevated circulating omega-3 PUFA concentrations, which modulate a range of biochemical pathways [9-11]. Several randomized clinical trials, such as STRENGTH [12], VITAL [13], and PISCES [4], have reported that daily omega-3 supplementation for 3 months or longer, containing both eicosapentaenoic acid (EPA) and docosahexaenoic acid (DHA), significantly increased circulating concentrations of the corresponding omega-3 fatty acids. Compared with baseline, 1g of omega-3 fatty acid supplementation led to relative increases of 34% to 82% for EPA, 12% to 20% for DHA, and 55% to 60% for total omega-3 fatty acids. While these existing studies have quantified the effects of specific omega-3 supplements (e.g., fish oil) in raising circulating omega-3 concentrations, few have considered the possible modifying effects of genetic factors.

Like dietary intake or supplements, genetic factors can regulate circulating omega-3 PUFA concentrations. Genome-wide association studies (GWAS) have identified hundreds of genetic loci associated with circulating PUFA concentrations, such as variants near *FADS1/2*, *MYRF*, *APOA1*, *LIPC*, *APOE*, and *GCKR* genes [14-18]. In addition to direct genetic influence, gene-by-environment interactions (GEI) have also been observed, where genetic variants modify the association between dietary PUFA intake and circulating PUFA concentrations. For instance, participants carrying the TT genotype of rs174546 at *FADS1* showed a greater increase (7.8% increase) in circulating EPA levels in response to dietary EPA intake than those with the CC genotype (3.7% increase) [19]. Similarly, a clinical trial observed that participants carrying the *APOE* ε4/ε4 genotype who received PUFA supplementation for 18 months exhibited the smallest decrease in the ratio of arachidonic acid to highly unsaturated fatty acids, and a smaller increase in the ratio of EPA to highly unsaturated fatty acids, compared to participants with other *APOE* genotypes [20]. Our recent genome-wide GEI study also identified variants in the *FADS1-FADS2* gene cluster and *GPR12* as modifiers of the associations between FOS and circulating omega-3 fatty acids [21].

While pioneering studies have identified several GEIs for circulating fatty acids, they typically focused on individual genetic variants or genes, which failed to account for an individual’s overall genetic background and polygenic predisposition to either high or low circulating fatty acid concentrations. Moreover, because interaction effects are generally smaller than direct genetic effects, single-variant GEI studies often lack statistical power to detect subtle interaction signals without a large sample size [22]. To address these challenges, a polygenic score (PGS) can be employed to aggregate genetic effects across the genome. This approach boosts statistical power, enabling a more effective evaluation of how genetic composition interacts with environmental factors [23]. For example, our recent PGS-by-FOS interaction study showed that FOS attenuates the associations between genetically predicted and observed circulating levels of total cholesterol, low-density lipoprotein cholesterol, and triglycerides [24]. Despite these advances, no study has yet explored the effects of PGS-by-FOS interactions on circulating omega-3 concentrations.

This study aimed to examine the PGS-by-FOS interaction on circulating omega-3 fatty acid concentrations. First, we developed four PGS models for Omega-3, Omega-3%, DHA and DHA% based on a multi-ethnic, independent genome-wide association study (N = 136,016). We evaluated the associations between PGS and the corresponding observed concentrations across a diverse cohort from the UK Biobank (UKB), comprising participants of European (EUR, N = 237,380), Central/South Asian (CSA, N = 8,237), African (AFR, N = 6,323), and East Asian (EAS, N = 2,571) ancestries. Second, we tested for interactions between PGS and FOS on four omega-3 concentrations in each population. Third, we replicated our findings using oily fish intake (OFI) as a secondary dietary exposure and an additional set of UKB EUR participants (N = 178,935). Lastly, sensitivity analyses validated our findings across two dietary questionnaires, two ways of phenotypic transformation (e.g., raw versus rank-based inverse normal transformed (RINT)), and two PGS approaches. Our study aspires to characterize the joint, nonlinear effects of genetic composition and FOS on circulating omega-3 profiles. By identifying specific genetic subgroups that respond differently to fish oil intake, we support the transition from generalized dietary guidelines to targeted dietary interventions, maximizing the health benefits of omega-3 intake and advancing the implementation of personalized nutrition strategies.

## Methods

### Study population

The data used in this study were sourced from the UKB, a large prospective cohort study involving over 500,000 participants aged 40-69 at recruitment from 2006 to 2010 [25]. Each participant signed an informed consent form, and ethics approval was acquired from the North West Multi-Centre Research Ethics Committee. At baseline, participants completed touchscreen questionnaires capturing demographic, lifestyle, and health information. Blood samples were collected at recruitment for genotyping and biochemical marker measurements. The participant’s ancestry was derived from Pan-UKB [26]. This study was conducted under UKB application number 48818.

Among 502,180 participants, we conducted quality control (QC) to exclude participants with: 1) missing genetic ancestry group data from Pan-UKB (N = 54,314); 2) withdrawn consent (N = 203); 3) ten or more third-degree relatives (N = 179); 4) discrepancies between self-reported and genetic sex (N = 339); 4) sex chromosomes aneuploidy (N = 432); 5) outliers for heterozygosity and missing genotype rate > 5% (N = 521) [27]; 5) predominantly admixed American (N = 987) or Middle Eastern (N = 1,614) ancestry as defined by Pan-UKB; 6) lacking circulating omega-3 fatty acid measurements (N = 5,788). Following exclusion, 437,803 individuals were included in the analyses, including 419,832 EUR, 8,703 CSA, 6,583 AFR, and 2,685 EAS participants (**Supplementary Figure 1**).

### Measures of circulating omega-3 fatty acids

The four phenotypes of interest in our study are the absolute concentrations of total omega-3 fatty acids and DHA (referred to as Omega-3 and DHA), and their relative percentages in total fatty acids (expressed as Omega-3% and DHA%). They were measured in plasma, collected at recruitment, using nuclear magnetic resonance (NMR) spectroscopy on the Nightingale Health platform. In 2023, phase 2 NMR metabolomics data were released, involving approximately 300,000 participants. Phase 3 NMR metabolomics data of approximately 200,000 additional participants were released in 2025. Our discovery analysis focused on EUR participants with metabolomic data from UKB phase 2 and non-EUR ancestry participants with all phases’ metabolomic data, while the additional EUR participants in phase 3 served as a replication dataset (**Figure 1 and Supplementary Figure 1**).

**Figure 1.**
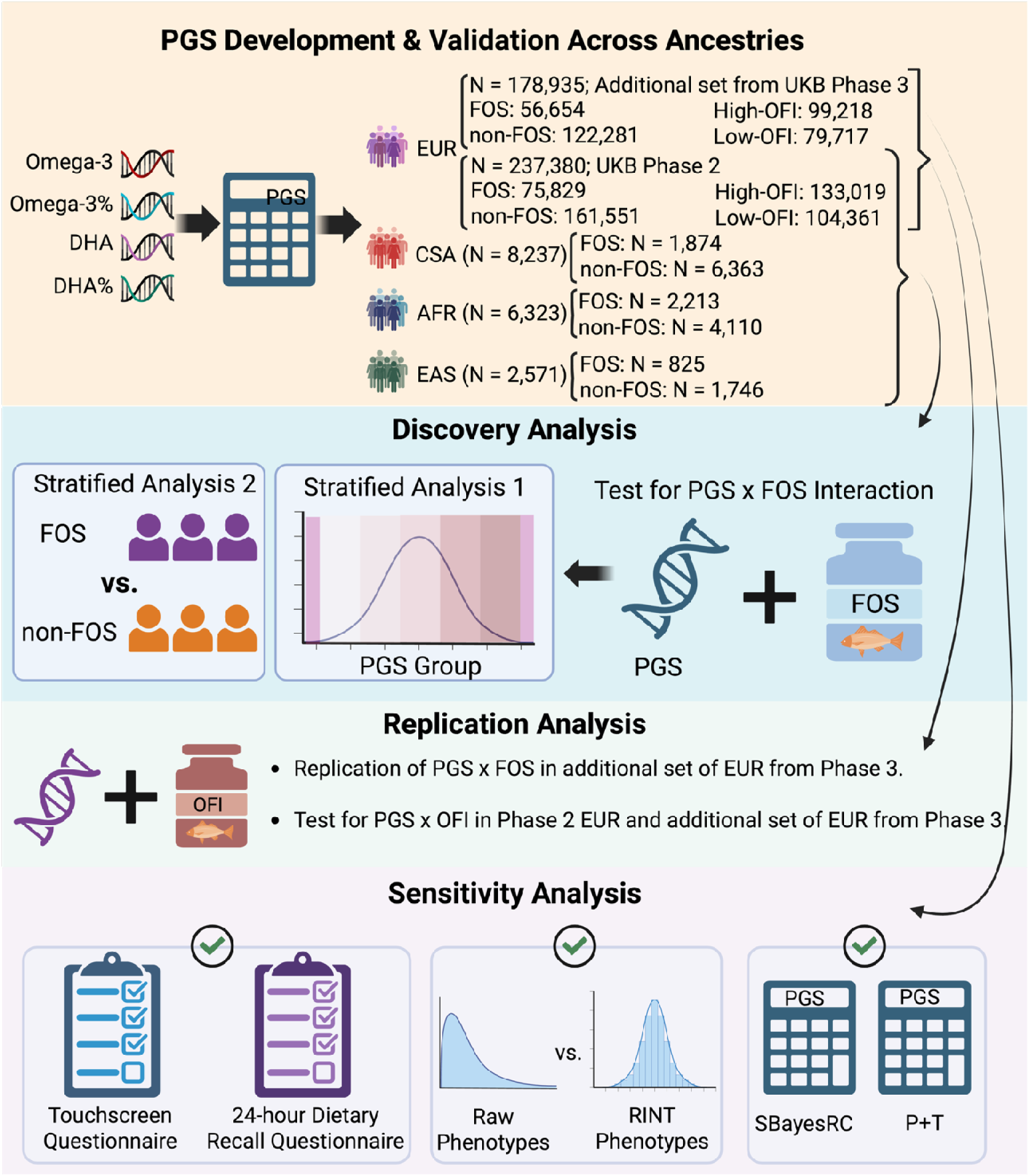
Schematic of study design. Fish oil supplementation (FOS) status was defined as FOS and non-FOS users based on self-reported intake from either the baseline touchscreen questionnaire or the 24-hour dietary recall. Oily fish intake (OFI) was categorized into high-OFI (consumption >= once per week) and low-OFI (consumption < once per week). Abbreviations: Omega-3, absolute concentration of total omega-3 fatty acids; Omega-3%, total omega-3 fatty acids to total fatty acids percentage; DHA, absolute concentration of docosahexaenoic acid; DHA%, docosahexaenoic acid to total fatty acids percentage; PGS, polygenic score; EUR, European; EAS, East Asian; AFR, African; CSA, Central/South Asian; UKB, UK Biobank; RINT, rank-based inverse normal transformation; FOS, fish oil supplementation; OFI, oily fish intake; P+T, pruning and thresholding.

### Dietary exposures

We examined fish oil supplementation (FOS) as our primary dietary exposure and oily fish intake (OFI) as a secondary dietary exposure **(Supplementary Tables 1 and 2**). We obtained FOS information from two questionnaires separately: the touchscreen questionnaire [28] at recruitment and the 24-hour dietary recall questionnaire at multiple follow-ups [29]. In the touchscreen questionnaire, participants were asked, “Do you regularly take any of the following? (You can select more than one answer)” (Data-field 6179). FOS users were defined as participants who answered yes on “Fish oil (including cod liver oil)”, while non-fish oil users (non-FOS) were defined as participants who selected only other supplements, chose “None of the above”, or did not provide a response. Responses of “Prefer not to answer” were excluded.

FOS status was also acquired from the online 24-hour dietary recall questionnaire, which was administered up to five times between 2009 and 2012. About 40% of the UKB participants completed at least one dietary recall [29]. To be eligible for this analysis, participants had to answer the question, “Did you have any vitamin or mineral supplements yesterday? (e.g., Vitamin C, multivitamins, fish oil, calcium supplement)” (data fields 104670 and 20084). Fish oil users were defined as those who had information on vitamin supplement use and answered yes to “Fish oil” for at least one recall. Non-FOS users were defined as participants who reported no supplement use or who reported supplements other than FOS in all returned assessments.

OFI status was obtained from the touchscreen questionnaire. Participants were asked, “How often do you eat oily fish? (e.g., sardines, salmon, mackerel, herring)” (data-field 1329). We divided participants into two groups. The high intake group of oily fish (high-OFI) was defined as participants who consumed oily fish once a week or more frequently, while the low intake group of oily fish (low-OFI) was defined as participants who consumed oily fish less than once a week. We excluded participants who answered, “Do not know” or “Prefer not to answer.”

### Genotype QC

Genotypic data used in the study were generated using UKB Axiom Array or UK BiLEVE array and imputed using Haplotype Reference Consortium and UK10K as reference panels, yielding approximately 96 million imputed variants [27]. Genetic data were further quality-controlled separately in each ancestry group to exclude variants with an imputation quality score < 0.3, minor allele frequency < 0.1%, genotype call rate < 0.95, and Hardy-Weinberg test P < 1X10^-8^. After QC, 13,083,408 autosomal genetic variants were retained for the EUR population, 10,973,292 for EAS, 12,484,527 for CSA, and 19,592,517 for AFR.

### Polygenic score construction

PGS for Omega-3, Omega-3%, DHA, and DHA% were constructed for UKB participants of EUR, CSA, AFR, and EAS ancestries. The training data is a multi-cohort GWAS meta-analysis encompassing 13,389,637 imputed autosomal single-nucleotide polymorphisms (SNPs) and up to 136,016 individuals, including 4,435 East Asians, 11,340 South Asians, and 120,241 Europeans. The phenotypes were measured in either serum or plasma [30]. The GWAS summary statistics for the four fatty acid traits were downloaded from the GWAS Catalog (Accession IDs: GCST90301955, GCST90301956, GCST90301959, GCST90301960) [30]. GWAS summary statistics and UKB genetic data were harmonized using MungSumstats [31] to ensure consistency in the reference genome build (i.e., GRCh37/hg19), SNP IDs, and allele information. Genetic variants shared between the GWAS summary statistics and the UKB cohort were extracted.

We constructed PGS using two approaches, SBayesRC [32] and pruning and thresholding (P+T) [33]. SBayesRC is a Bayesian multiple regression model that has been shown to improve genomic prediction by integrating GWAS summary statistics with linkage disequilibrium (LD) information and functional genomic annotations to estimate joint SNP effects for PGS calculations. We generated an in-sample LD reference from 239,268 UKB participants of EUR ancestry. Functional annotations, such as histone mark, coding and conserved regions, were obtained from the baseline model 2.2 developed by the Price group [34, 35]. PGS for each participant in the four ancestry groups was calculated as the sum of posterior SNP effects multiplied by effect allele dosages. For P+T, we selected genome-wide significant variants (P < 5X10^-8^) and pruned them (r^2^ = 0.1, window size = 250Kb) using in-sample EUR LD in PLINK. Across the four fatty acid traits, 110 to 214 independent, significant SNPs were obtained using the P+T approach, and 13,083,387 variants were included in the SBayesRC-derived PGS. PGS was calculated as the weighted sum of the effect allele dosages of these independent SNPs, with their effect sizes as weights. All PGS were standardized to a mean of 0 and a standard deviation (SD) of 1.

### Covariates

To control for potential confounding, we included in the analysis: sex, age at recruitment, interaction between sex and age, body mass index (BMI), Townsend index (TSI), smoking status, alcohol drinking status, physical activity, and statin use. Information on statin use was sourced from self-reported medications through verbal interview at recruitment. The remaining covariates were obtained from the touchscreen questionnaire at recruitment. TSI is a measure of socioeconomic status calculated from four census variables, including households without a car, overcrowded households, households not owner-occupied, and unemployed persons [36]. Smoking status and alcohol drinking status were defined as current, previous, or never. Physical activities were classified into high, moderate, and low. About 0.1% to 22.7% of participants had missing covariate information on TSI, BMI, smoking status, alcohol drinker status, and physical activity; we therefore imputed missing data using the random forest method with the mice R package [37]. After confirming the convergence and consistency across five imputation iterations, the first imputed dataset was selected for all downstream analyses.

### Statistical analysis

All statistical analyses and visualizations were conducted under R 4.3.1 version. Outliers for each fatty acid were defined as values that fell outside the 1.5 times interquartile ranges and were subsequently removed. Fatty acid concentrations were then rank-based inverse normal transformed (RINT). Accordingly, 1 unit change in the RINT-based fatty acid measure corresponds to a change of 1 SD in the standard normal distribution. We reported Pearson and Spearman correlation coefficients among covariates, between covariates and PGS, between PGS and observed circulating fatty acid levels, and between RINT-based and raw fatty acid levels (**Supplementary Table 3**).

Multivariable linear regression models with and without PGS-by-diet interaction terms were performed. Weak correlations between covariates were observed, indicating that they are largely independent (**Supplementary Figure 2**). Therefore, both models included the following covariates: sex, age, sex-by-age, BMI, TSI, smoking status, alcohol drinking status, physical activity, statin use, FOS, OFI, PGS, and the top 10 genetic PCs derived from the Pan-UKB. To assess whether PGS modifies the association of FOS or OFI with the omega-3 fatty acid levels, we conducted stratified analysis by PGS subgroups, including the bottom 5% (0–5%), five quintile-based bins (0–20%, 20–40%, 40–60%, 60–80%, and 80–100%), and the top 5% (95–100%) of the PGS distribution. We also performed stratified analysis by FOS or OFI status to examine whether the association effects of PGS with the observed omega-3 fatty acid levels vary in FOS and non-FOS groups, or in high-OFI and low-OFI groups. A p-value of 0.05 was set as a significant threshold. Partial R^2^ values were estimated using rsq R package [38] to assess the unique contribution of each variable on the phenotypic variance in fatty acid concentrations when controlling for all other covariates.

Our discovery analysis was restricted to (i) EUR participants with metabolomic data from UKB phase 2 and non-EUR participants from all available phase 3 metabolomic data; (ii) FOS data derived from the baseline touchscreen questionnaire; (iii) RINT-based fatty acid concentrations; (iv) PGS constructed with SBayesRC. To ensure the robustness of our findings, we replicated these analyses by using OFI as a secondary dietary exposure and leveraging an additional set of EUR participants from phase 3 metabolomic data who were not included in the phase 2 release. Finally, we performed a series of sensitivity analyses that (i) replaced touchscreen-based ascertainment of FOS status with the 24-hour dietary recall questionnaire; (ii) analyzed raw (i.e., untransformed) fatty acid concentrations; (iii) used P+T-derived PGS (**Figure 1**).

To estimate the sample size required to detect moderate interaction effects, we performed power calculations using run.espresso.ExE function within ESPRESSO [39]. For simulations, both PGS and FOS were modeled as environmental exposures. Using RINT-based Omega-3 as a representative continuous phenotype (mean = 0 and SD = 1), we treated PGS as a continuous variable (effect = 0.3) and FOS as a binary variable (prevalence =0.3, effect = 0.36). At a nominal significance level of 0.05, we estimated the sample size required to detect a fixed interaction effect of 0.04. We also estimated the sample size required across a range of potential interaction effects.

## Results

### Participant characteristics

This study included a total of 437,803 UKB participants (**Figure 1** and **Supplementary Figure 1**). Among EUR participants (N = 237,380) in the discovery analysis (**Table 1)**, the mean age was 57 years, the mean BMI was 27 kg/m^2^, and 54% were female. Approximately 54% reported never smoking, and over 95% reported previous or current alcohol use. Approximately 16% of participants self-reported using statin medications. About 32% of participants reported FOS, and 56% consumed oily fish more than once a week (**Supplementary Tables 1 and 2**). Participants in the FOS and high-OFI groups generally had higher concentrations of Omega-3, Omega-3%, DHA, and DHA% than those in non-FOS and low-OFI groups (**Supplementary Tables 1 and 2**).

**Table 1.** Baseline characteristics of participants in the UKB. Values are reported as counts (%) for categorical variables and means (SD) for continuous variables. Abbreviations: BMI, body mass index; Omega-3, absolute concentration of total omega-3 fatty acids; Omega-3%, total omega-3 fatty acids to total fatty acids percentage; DHA, absolute concentration of docosahexaenoic acid; DHA%, docosahexaenoic acid to total fatty acids percentage; SD, standard deviation.

|  | European |  | Central/South Asian | African | East Asian |
| --- | --- | --- | --- | --- | --- |
|  | Discovery analysis<br>(N = 237,380) | Replication analysis<br>(N = 178,935) | (N = 8,237) | (N = 6,323) | (N = 2,571) |
| Age, years (SD) | 56.87 (8.01) | 57 (8.0) | 53.50 (8.48) | 52.38 (8.10) | 52.22 (7.86) |
| Sex, female (%) | 128,068 (53.95) | 97,459 (54.47) | 3,821 (46.39) | 3,787 (59.89) | 1,708 (66.43) |
| BMI, kg/m<sup>2</sup> (SD) | 27.43 (4.76) | 27.35 (4.76) | 27.18 (4.41) | 29.66 (5.45) | 24.58 (3.75) |
| Omega-3, mmol/L (SD) | 0.53 (0.22) | 0.55 (0.23) | 0.48 (0.23) | 0.59 (0.25) | 0.56 (0.27) |
| Omega-3% (SD) | 4.37 (1.53) | 4.36 (1.49) | 3.92 (1.54) | 5.60 (1.93) | 4.58 (1.87) |
| DHA, mmol/L (SD) | 0.24 (0.08) | 0.24 (0.09) | 0.21 (0.08) | 0.28 (0.10) | 0.26 (0.10) |
| DHA% (SD) | 1.99 (0.67) | 1.98 (0.67) | 1.76 (0.67) | 2.64 (0.79) | 2.14 (0.81) |
| Smoking status (%) |  |  |  |  |  |
| Never | 128,792 (54.26) | 97,198 (54.32) | 6,458 (78.40) | 4,461 (70.55) | 1,921 (74.72) |
| Previous | 84,206 (35.47) | 63,500 (35.49) | 1,043 (12.66) | 1,102 (17.43) | 434 (16.88) |
| Current | 24,382 (10.27) | 18,237 (10.19) | 736 (8.94) | 760 (12.02) | 216 (8.40) |
| Alcohol status (%) |  |  |  |  |  |
| Never | 7,373 (3.11) | 5,632 (3.15) | 3,301 (40.08) | 960 (15.18) | 532 (20.69) |
| Previous | 8,182 (3.45) | 6,195 (3.46) | 460 (5.58) | 331 (5.23) | 115 (4.47) |
| Current | 221,825 (93.45) | 167,108 (93.39) | 4,476 (54.34) | 5,032 (79.58) | 1,924 (74.83) |
| Physical activity (%) |  |  |  |  |  |
| Low | 43,714 (18.42) | 32,377 (18.09) | 1,976 (23.99) | 1,256 (19.86) | 487 (18.94) |
| Moderate | 96,503 (40.65) | 73,817 (41.25) | 3,249 (39.44) | 2,395 (37.88) | 1,054 (41.00) |
| High | 97,163 (40.93) | 72,741 (40.65) | 3,012 (36.57) | 2,672 (42.26) | 1,030 (40.06) |
| Statin use, yes (%) | 39,813 (16.77) | 28,336 (15.84) | 2,131 (25.87) | 939 (14.85) | 267 (10.39) |

### Association between FOS, OFI, and PGS with circulating omega-3 fatty acids

Both FOS and OFI were significantly associated with circulating concentrations of Omega-3, Omega-3%, DHA, and DHA% (**Figure 2** and **Supplementary Tables 3-4**). For example, FOS was associated with a 0.36 SD increase (95% CI: 0.35 - 0.37; P < 1X10^-300^) in RINT-based Omega-3, and high-OFI with a 0.45 SD increase (95% CI: 0.45 - 0.46; P < 1X10^-300^) in RINT-based Omega-3 (**Figure 2A**). For raw Omega-3, FOS was associated with a 0.068 mmol/L increase (95% CI: 0.067 - 0.070; P < 1X10^-300^) and high-OFI with a 0.083 mmol/L increase (95% CI: 0.083 - 0.086; P < 1X10^-300^). OFI and FOS accounted for 6.0 – 9.8% and 3.5 - 4.8% of phenotypic variance in four RINT-based omega-3 fatty acid concentrations (**Figure 2B and Supplementary Figure 3**).

**Figure 2.**
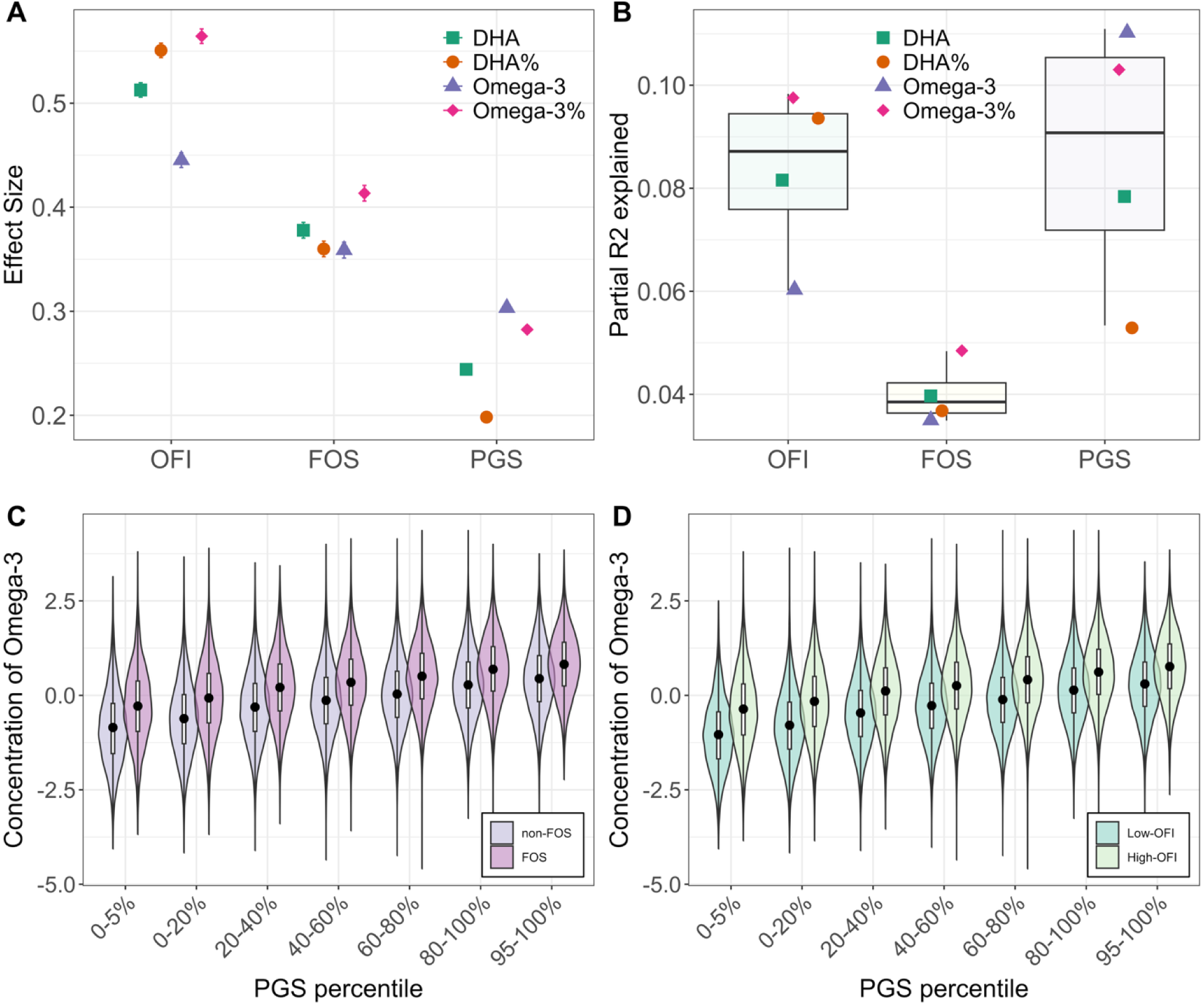
Associations of FOS, OFI, and PGS with circulating concentrations of omega-3 fatty acids in EUR participants. The associations between FOS, OFI, and PGS with four RINT-based omega-3 fatty acid traits (DHA, DHA%, Omega-3, and Omega-3%) were tested using models without interaction terms. **A)** estimated effect sizes of FOS, OFI, and PGS on circulating concentrations of omega-3 fatty acids. **B)** Proportion of phenotypic variance (partial R2) in the four RINT-based omega-3 fatty acids explained by FOS, OFI, and PGS. Increased PGS was consistently associated with higher absolute concentrations of total omega-3 fatty acids, independent of **C)** FOS and **D)** OFI status.

Four PGS models for circulating levels of Omega-3, Omega-3%, DHA, and DHA%, were developed using SBayesRC. Higher PGS for all four fatty acids were significantly associated with higher corresponding observed fatty acid concentrations regardless of FOS or OFI status (**Figures 2C and 2D**). PGS derived from SBayesRC showed slightly stronger correlation with the traits (Pearson r: 0.21 – 0.31; P < 2.2X 10^-16^) than PGS from the P+T approach (Pearson r: 0.18 – 0.27; P < 2.2X10^-16^) (**Supplementary Table 3**). Correlations were also consistently stronger for RINT-based phenotypes than for raw, untransformed measures (**Supplementary Figure 4**). Accordingly, we reported results primarily for SBayesRC-derived PGS and RINT-based phenotypes, while results for raw phenotypes were provided in the supplementary materials for easier clinical interpretation.

We estimated the association effects of PGS on the observed absolute concentrations and relative percentages of circulating omega-3 fatty acids (**Supplementary Table 4**). Per 1 SD increase in PGS was significantly associated with 0.24 SD (95% CI: 0.24 - 0.25; P < 1X10^-300^), 0.20 SD (95% CI: 0.20 - 0.21; P < 1X10^-300^), 0.30 SD (95% CI: 0.30 - 0.31; P < 1X10^-300^), and 0.28 SD (95% CI: 0.28 - 0.29; P < 1X10^-300^) increases in DHA, DHA%, Omega-3, and Omega-3%, respectively. While FOS and OFI were strongly associated with observed fatty acid levels, the estimated effects of the two dietary factors were notably greater than those of the PGS (**Figure 2A**). For example, RINT-based Omega-3 levels increased by 0.45 SD for high-OFI and 0.36 SD for FOS, representing 1.5-fold and 1.2-fold greater association effect than that observed for PGS (0.30 SD). In terms of phenotypic variance explained, PGS accounted for 5.3 - 11.1% of trait variance, similar to OFI and higher than FOS (Figure 2B).

### Evidence of PGS-by-FOS interactions on circulating omega-3 fatty acids in EUR participants

We observed evidence of interaction between PGS and FOS on circulating concentrations of DHA (P_Int_ = 1.52X 10^-11^), DHA% (P_Int_ = 5.91X 10^-07^), Omega-3 (P_Int_ = 4.38X 10^-25^), and Omega-3% (P_Int_ = 1.02 X 10^-30^; **Figure 3 and Supplementary Table 5**). While FOS was associated with increases in all four circulating omega-3 fatty acids at all levels of PGS (**Supplementary Figure 5**), the magnitude of the estimated association effect decreased as PGS increased, suggesting a gradient of response (**Figures 3A-3D and Table 2**). Among participants in the bottom 5% of the PGS distribution, FOS was significantly associated with a 0.40 SD (95% CI: 0.39 – 0.44; P = 5.15X10^-102^) increase in the RINT-based Omega-3 level. This association effect was 11.1% larger than the population average (β = 0.36; 95% CI: 0.35 – 0.37; P = 1X10^-300^; P_Int_ = 0.016) and 42.8% larger than the association effect observed in participants in the top 5% (>95^th^ percentile) of the PGS distribution (β = 0.28 SD; 95% CI: 0.25 – 0.32; P = 4.94X10^-63^; P_Int_ = 4.03X10^-10^). For raw Omega-3 levels (**Supplementary Table 6)**, the corresponding association effects were 0.067 mmol/L increase (95% CI: 0.061 – 0.074; P = 1.54X10^-99^) in the bottom 5% and 0.059 mmol/L increase (95% CI: 0.051 – 0.065; P = 7.33X10^-64^) in the top 5% of PGS distribution, a 14% increase of the FOS association effect (P_Int_ = 0.022).

**Figure 3.**
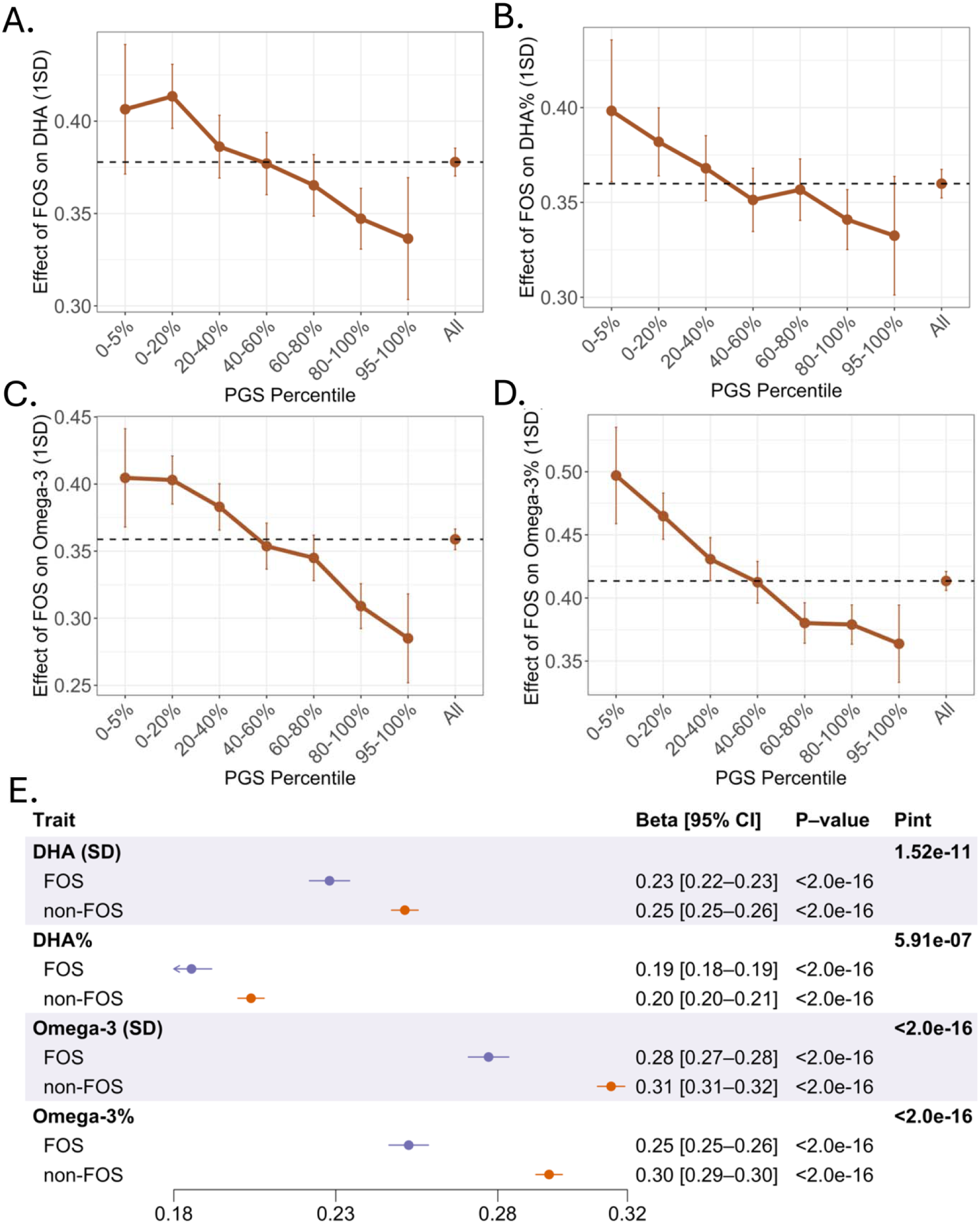
Evidence of PGS-by-FOS interactions on RINT-based circulating concentrations of omega-3 fatty acids in EUR participants. **A) – D)** PGS modified the positive associations of FOS with circulating levels of DHA, DHA%, Omega-3, Omega-3%. The X-axis categories PGS groups, and the Y-axis represents the association effect sizes of FOS on increasing circulating fatty acid levels. PGS was calculated using the SBayesRC algorithm, and RINT-based phenotypes were analyzed. **E)** PGS was associated with smaller increases in circulating omega-3 fatty acids among individuals in the FOS group compared to those in the non-FOS group. Estimated association effect sizes of PGS are presented as beta [95% CI] in the forest plot. P-value denotes the significance of the association between PGS and omega-3 concentrations within each stratum, while P_Int_ represents the interaction p-value between PGS and FOS.

**Table 2.** Association effects of FOS or high-OFI with circulating omega-3 concentrations stratified by PGS groups in EUR participants. Effect estimates and their 95% CI are reported. PGS was calculated using the SBayesRC algorithm, and RINT-based phenotypes were analyzed. P_Int_ represents the interaction p-value between PGS and FOS, derived from an analysis comprising participants in the top 5% and bottom 5% of the PGS distribution.

| Traits | Effects of dietary exposure ($\beta_{\text{diet}}$ [95% CI]) by PGS groups | | | | | | | $P_{\text{Int}}$<br>(0-5% vs 95-100% PGS) |
| --- | --- | --- | --- | --- | --- | --- | --- | --- |
|  | 0-5% | 0-20% | 20-40% | 40-60% | 60-80% | 80-100% | 95-100% |  |
| Dietary exposure: FOS |  |  |  |  |  |  |  |  |
| DHA | 0.406<br>(0.371, 0.441) | 0.413<br>(0.396, 0.431) | 0.386<br>(0.369, 0.403) | 0.377<br>(0.360, 0.394) | 0.365<br>(0.349, 0.382) | 0.347<br>(0.331, 0.364) | 0.336<br>(0.303, 0.369) | $3.20\times 10^{-04}$ |
| DHA% | 0.398<br>(0.361, 0.436) | 0.382<br>(0.364, 0.400) | 0.368<br>(0.351, 0.385) | 0.351<br>(0.335, 0.368) | 0.357<br>(0.341, 0.373) | 0.341<br>(0.325, 0.357) | 0.332<br>(0.301, 0.364) | $7.05\times 10^{-04}$ |
| Omega-3 | 0.405<br>(0.368, 0.441) | 0.403<br>(0.385, 0.421) | 0.383<br>(0.366, 0.400) | 0.354<br>(0.337, 0.371) | 0.345<br>(0.328, 0.362) | 0.309<br>(0.292, 0.326) | 0.285<br>(0.252, 0.318) | $4.03\times 10^{-10}$ |
| Omega-3% | 0.497<br>(0.459, 0.535) | 0.465<br>(0.447, 0.483) | 0.431<br>(0.414, 0.448) | 0.412<br>(0.396, 0.429) | 0.380<br>(0.364, 0.396) | 0.379<br>(0.363, 0.394) | 0.364<br>(0.333, 0.394) | $1.07\times 10^{-11}$ |
| Dietary exposure: OFI |  |  |  |  |  |  |  |  |
| DHA | 0.562<br>(0.530, 0.595) | 0.558<br>(0.542, 0.574) | 0.519<br>(0.504, 0.535) | 0.508<br>(0.492, 0.524) | 0.490<br>(0.474, 0.505) | 0.487<br>(0.472, 0.502) | 0.483<br>(0.452, 0.514) | $4.76\times 10^{-05}$ |
| DHA% | 0.620<br>(0.586, 0.654) | 0.590<br>(0.573, 0.607) | 0.561<br>(0.545, 0.577) | 0.543<br>(0.527, 0.558) | 0.529<br>(0.514, 0.545) | 0.531<br>(0.516, 0.546) | 0.531<br>(0.502, 0.561) | $4.84\times 10^{-06}$ |
| Omega-3 | 0.557<br>(0.523, 0.591) | 0.507<br>(0.490, 0.523) | 0.471<br>(0.455, 0.488) | 0.429<br>(0.413, 0.445) | 0.427<br>(0.411, 0.442) | 0.392<br>(0.376, 0.407) | 0.380<br>(0.349, 0.411) | $1.52\times 10^{-18}$ |
| Omega-3% | 0.719<br>(0.684, 0.754) | 0.655<br>(0.638, 0.672) | 0.589<br>(0.574, 0.605) | 0.544<br>(0.529, 0.559) | 0.522<br>(0.507, 0.537) | 0.513<br>(0.498, 0.527) | 0.514<br>(0.485, 0.542) | $1.54\times 10^{-24}$ |

Although PGS for all four fatty acids was significantly associated with increased concentrations of the corresponding fatty acids regardless of FOS or OFI status (**Figures 2C, 2D and Supplementary Figure 5**), the magnitude of this increase was smaller among individuals in the FOS group compared to those in the non-FOS group (**Figure 3E and Table 3**). For example, per 1 SD increase in PGS was associated with a 0.23 SD (95% CI: 0.22 – 0.23; P = 1X10^-300^) increase in DHA in the FOS group and a 0.25 SD (95% CI: 0.25 - 0.26; P = 1X10^-300^) increase in the non-FOS group. Per 1 SD increase in PGS was associated with a 0.28 SD (95% CI: 0.27 – 0.28; P = 1X10^-300^) increase in Omega-3 in the FOS group and a 0.31 SD (95% CI: 0.31 - 0.32; P = 1X10^-300^) increase in the non-FOS group. To enhance clinical interpretation, we also reported the results based on raw phenotypes (**Supplementary Figure 6 and Supplementary Table 7**). Per 1 SD increase in PGS, Omega-3 increases by 0.054 mmol/L (95% CI: 0.05 – 0.06; P = 1X10^-300^) in the FOS group and by 0.056 mmol/L (95% CI: 0.06 – 0.06; P = 1X10^-300^) in the non-FOS group (P_Int_ < 2.77X10^-04^).

**Table 3.** Associations of PGS with corresponding observed circulating omega-3 concentrations stratified by FOS or OFI in EUR participants. The number of participants used in the statistical analyses is reported as n. The association effect of PGS on circulating omega-3 concentrations is expressed as β_PGS_ (95%CI). FOS and OFI were the dietary exposures. PGS was calculated using the SBayesRC algorithm, and RINT-based phenotypes were analyzed.

| Trait | Exposed group |  | Control group |  | P<sub>Int</sub> |
| --- | --- | --- | --- | --- | --- |
|  | n | β<sub>PGS</sub> (95% CI) | n | β<sub>PGS</sub> (95% CI) |  |
| Dietary exposure: FOS |  |  |  |  |  |
| DHA | 72,024 | 0.228 (0.222, 0.234) | 158,667 | 0.251 (0.247, 0.255) | 1.52×10<sup>-11</sup> |
| DHA% | 72,400 | 0.185 (0.179, 0.192) | 158,652 | 0.204 (0.200, 0.208) | 5.91×10<sup>-07</sup> |
| Omega-3 | 72,313 | 0.277 (0.271, 0.283) | 158,930 | 0.315 (0.311, 0.319) | 4.38×10<sup>-25</sup> |
| Omega-3% | 71,971 | 0.253 (0.246, 0.259) | 158,686 | 0.296 (0.292, 0.300) | 1.02×10<sup>-30</sup> |
| Dietary exposure: OFI |  |  |  |  |  |
| DHA | 127,073 | 0.232 (0.227, 0.236) | 103,618 | 0.259 (0.255, 0.264) | 1.56×10<sup>-17</sup> |
| DHA% | 127,378 | 0.186 (0.181, 0.191) | 103,674 | 0.214 (0.209, 0.218) | 1.11×10<sup>-14</sup> |
| Omega-3 | 127,714 | 0.283 (0.278, 0.287) | 103,529 | 0.329 (0.323, 0.334) | 2.70×10<sup>-41</sup> |
| Omega-3% | 126,840 | 0.255 (0.250, 0.259) | 103,817 | 0.317 (0.312, 0.322) | 4.12×10<sup>-70</sup> |

### Replication of PGS-by-FOS interactions on circulating omega-3 fatty acids in EUR participants

To ensure the robustness of the findings, we 1) replicated the PGS-by-FOS analysis using an additional set of EUR participants from UKB phase 3 metabolomic data (N = 179,935) and 2) tested for PGS-by-OFI interaction using OFI as a secondary dietary exposure in UKB phase 2 EUR participants and the additional set of EUR participants from phase 3. The weak correlation between FOS and OFI among EUR participants (Pearson r: 0.11) indicates that these measures capture unique aspects of dietary omega-3 intake, providing an additional layer of validation (**Supplementary Figure 2 and Supplementary Table 3**).

Replication of PGS-by-FOS showed consistent results with the discovery analysis. We demonstrated that 1) PGS modified the association effect of FOS on increasing circulating concentrations of DHA, DHA%, Omega-3, and Omega-3%; and 2) PGS was associated with smaller increase in circulating omega-3 fatty acid concentrations among individuals in the FOS/high-OFI group compared to those in the non-FOS/low-OFI group (**Supplementary Tables 8-10 and Supplementary Figures 7-8**). The estimated association effects of FOS and PGS on increasing omega-3 fatty acid traits are very close to those from the discovery analysis (**Supplementary Tables 9-10**).

Consistent with findings in PGS-by-FOS interactions, PGS modifies the associations between OFI and circulating concentrations of DHA (P_Int_ = 1.56X10^-17^), DHA% (P_Int_ = 1.11X10^-14^), Omega-3 (P_Int_ = 2.70 X 10^-41^), and Omega-3% (P_Int_ = 4.12 X 10^-70^) (**Figure 4 and Supplementary Table 8-9**). The association effect of high-OFI on elevating the circulating omega-3 fatty acids decreased as the PGS increased. Among participants in the bottom 5% of the PGS distribution, high-OFI was associated with a 0.56 SD (95% CI: 0.52 – 0.59; P = 2.21X10^-220^) increase in RINT-based Omega-3, representing a 24.4% larger association effect (P_Int_ = 2.14X10^-10^) than the population average estimate (β = 0.45; 95% CI: 0.44 – 0.45; P < 1.0X 10^-300^). Furthermore, the association effect is 47% larger than that of participants in the top 5% of the PGS distribution (β = 0.38; 95% CI: 0.35 – 0.41; P = 1.07X10^-124^; P_Int_= 1.52X10^-18^; **Table 2**). For raw Omega-3, the corresponding association effects were 0.092 mmol/L increase (95% CI: 0.086 – 0.098; P = 1.10X10^-211^) in the bottom 5% and 0.078 mmol/L increase (95% CI: 0.072 – 0.084; P = 7.25X10^-126^) in the top 5% of the PGS distribution, an 15% decrease of the OFI association effect (P_Int_ = 5.10X10^-4^; **Supplementary Figure 9 and Supplementary Table 9**).

**Figure 4.**
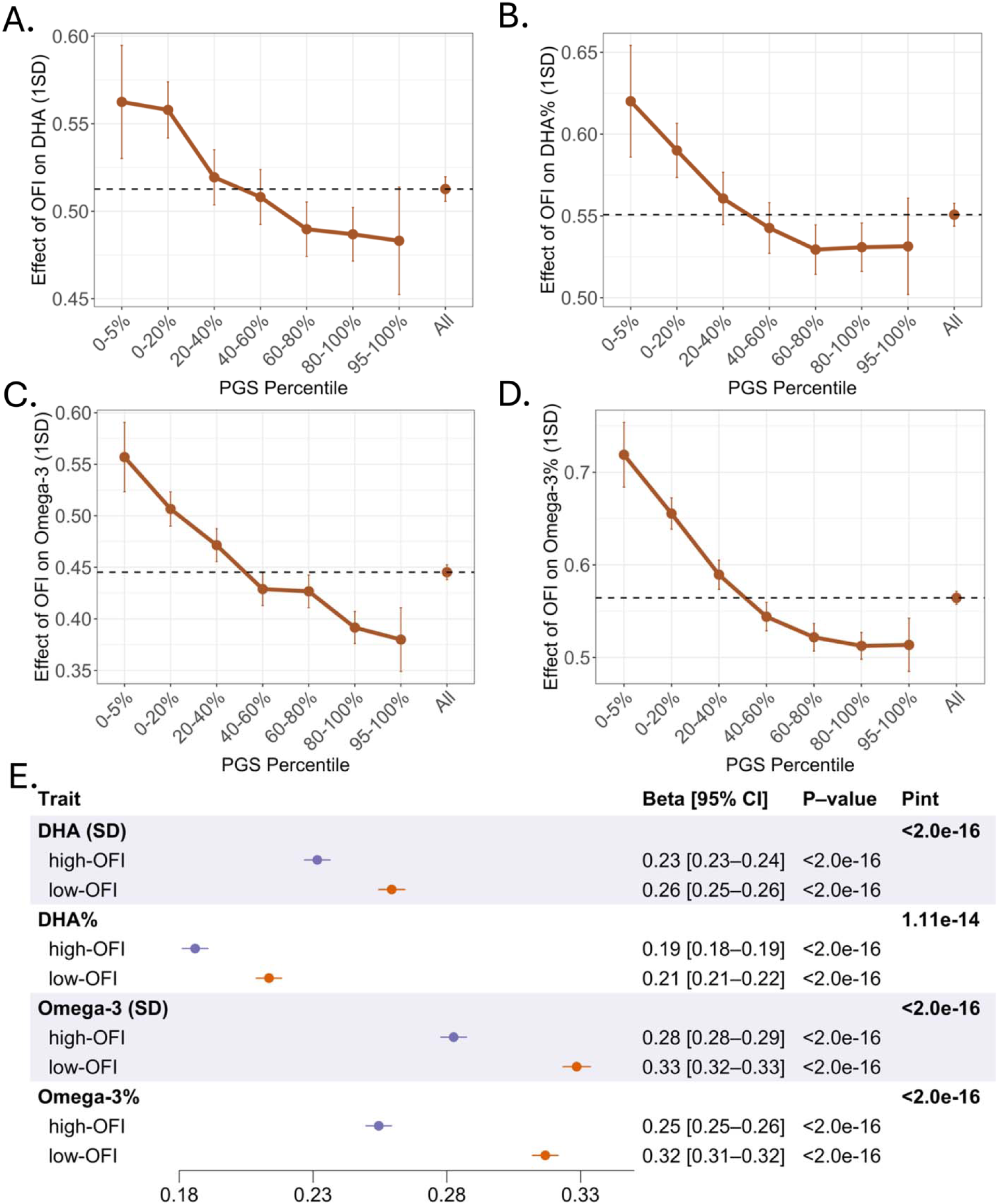
Evidence of PGS-by-OFI interactions on RINT-based circulating concentrations of omega-3 fatty acids in EUR participants. **A) – D)** PGS modified the positive associations of OFI with circulating concentrations of DHA, DHA%, Omega-3, Omega-3%. The X-axis categories PGS groups, and the Y-axis represents the association effect sizes of high-OFI on increasing circulating fatty acid levels. PGS was calculated using the SBayesRC algorithm, and RINT-based phenotypes were analyzed. **E)** PGS was associated with smaller increases in circulating omega-3 fatty acids among individuals in the high-OFI group compared to those in the low-OFI group. Estimated association effect sizes of PGS are presented as beta [95% CI] in the forest plot. P-value denotes the significance of the association between PGS and omega-3 concentrations within each stratum, while P_Int_ represents the interaction p-value between PGS and FOS.

PGS was associated with smaller increases in the four circulating fatty acids among individuals in the high-OFI group compared to those in the low-OFI group (**Figure 4E and Supplementary Table 10**). Per 1 SD increase in PGS was associated with a 0.28 SD increase (95% CI: 0.28 – 0.29; P = 1X10^-300^) in RINT-based Omega-3 in the high-OFI group and a 0.33 SD increase (95% CI: 0.32 - 0.33; P = 1 X 10^-300^) in the low-OFI group (P_Int_ = 2.70 X 10^-41^; **Supplementary Table 8**). For raw Omega-3, per 1 SD increase in PGS was associated with 0.054 mmol/L increase (95% CI: 0.053 – 0.055; P = 1X10^-300^) in the high-OFI group and 0.057 mmol/L increase (95% CI: 0.056 – 0.058; P = 1X10^-300^) in the low-OFI group (P_Int_ = 1.04X10^-06^; **Supplementary Tables 8and 10**).

### PGS-by-FOS and PGS-by-OFI interactions on circulating omega-3 fatty acids detected in non-European participants

FOS and OFI were each significantly associated (P < 0.05) with increasing levels of all investigated circulating fatty acid measures (DHA, DHA%, Omega-3, and Omega-3%) in participants of CSA, AFR, and EAS ancestries (**Supplementary Table 11**). PGS was significantly associated with all four fatty acid measures in CSA, but only with the corresponding DHA and Omega-3 in AFR and EAS ancestries. Averaging across the four fatty acid measures, OFI accounted for largest proportion of phenotypic variance (7% in CSA, 6% in EAS, and 5% in AFR), followed by FOS (2.7% in CSA, 1.3% in EAS, and 0.6% in AFR) and PGS (2.5% in CSA, 0.2% in EAS, and 0.2% in AFR).

We detected statistically significant interactions between PGS and FOS for DHA% (P_Int_ = 0.017; **Supplementary Figure 10**), and between PGS and OFI for Omega-3 (P_Int_ = 0.007) and Omega-3% (P_Int_ = 0.006) in CSA (**Supplementary Table 11 and Supplementary Figure 11**). Consistent with findings in the EUR population, the effect of OFI on increasing Omega-3 and Omega-3% decreased with increasing PGS (**Supplementary Table 12**). Among participants in the bottom 5% of the PGS distribution, high-OFI was associated with a 0.55 SD (95% CI: 0.35–0.76, P = 1.12X10^-07^) increase in RINT-based Omega-3. This association effect was 22.2% larger than the population average (β = 0.45; 95% CI: 0.40 – 0.49, P = 4.66X10^-94^) and 66.7% larger than that observed in the top 5% of the PGS distribution (β = 0.33; 95% CI: 0.14 – 0.53, P = 9.28 X 10^-04^). Furthermore, FOS and OFI also attenuated the association between PGS and observed levels of DHA%, Omega-3, and Omega-3% in CSA. Per 1 SD increase in PGS was associated with a 0.17 SD increase (95% CI: 0.14 – 0.20; P = 5.33X10^-25^) in Omega-3 in the high-OFI group and a 0.21 SD increase (95% CI: 0.18 - 0.23; P = 3.27X10^-55^) in low-OFI group (P_Int_ = 9.37X10^-03^; **Supplementary Table 13**). Overall, we observed the consistent PGS-by-FOS and PGS-by-OFI interaction patterns in the CSA population.

The interactions between PGS and either FOS or OFI for four circulating omega-3 fatty acids did not reach statistical significance in EAS and AFR participants (P_Int_ > 0.05), likely due to the combined issue of limited PGS transferability in non-EUR populations and small sample size (**Supplementary Figure 12 and Supplementary Table 11**).

### Extensive sensitivity analyses yielded consistent findings on PGS-by-FOS interactions

To further evaluate the robustness of our findings, we conducted sensitivity analyses by (i) replacing touchscreen-based ascertainment of FOS status with that from the 24-hour dietary recall questionnaire; (ii) analyzing raw (i.e., untransformed) fatty acid concentrations; and (iii) using P+T-derived PGS.

Using the FOS status from the 24-hour dietary recall questionnaire as the dietary exposure, we also detected significant PGS-by-FOS interactions for the RINT-based DHA (P_Int_ = 1.27X 10^-7^), DHA% (P_Int_ = 5.49X 10^-7^), Omega-3 (P_Int_ = 2.36X 10^-12^), and Omega-3% (P_Int_ = 2.65X10^-19^; **Supplementary Figure 13 and Supplementary Table 14**). Across raw fatty acid concentrations and P+T-derived PGS, we also observed consistent association and interaction patterns (**Supplementary Table 14**). Consistent with discovery analysis, the estimated effect of consuming fish oil on increasing circulating concentrations of omega-3 fatty acids decreased with increasing PGS (**Supplementary Table 15**), and the estimated effect of PGS on the observed omega-3 fatty acids in the non-FOS group was stronger than that in the FOS group (**Supplementary Table 16**). In short, our sensitivity analyses support the robustness of our findings of PGS-by-FOS interactions on circulating omega-3 fatty acids.

## Discussion

Our cross-sectional investigation of multi-ancestry UKB participants found that PGS accounted for 5.3 - 11.1% of trait variance in EUR participants. We found strong evidence of interactions between PGS and FOS on circulating DHA, DHA%, Omega-3, and Omega-3%. Specifically, PGS modify the associations between FOS and circulating fatty acids. Participants with polygenic predisposition to lower circulating omega-3 fatty acids (i.e., lower PGS) may derive greater benefit from taking FOS. In addition, PGS was associated with a smaller increase in all four circulating omega-3 fatty acid levels among individuals in the FOS group compared to those in the non-FOS group. Replication using OFI as a secondary dietary exposure and an additional set of EUR participants from the UKB phase 3 metabolomic data confirmed these findings. We also detected significant interactions between PGS and FOS for DHA%, and between PGS and OFI for Omega-3 and Omega-3% in CSA participants. Our findings support the interactions between habitual fish intake and polygenic predisposition captured by PGS on circulating omega-3 fatty acids, calling for precision nutrition tailored to an individual’s genetic background. We suggest that genome-informed dietary intervention, especially targeting individuals with polygenic predisposition to lower circulating omega-3 levels, can maximize the beneficial effect of FOS and improve long-term health outcomes.

Our PGS-by-FOS interaction analysis revealed that individuals with polygenic predisposition to lower circulating omega-3 fatty acids, as captured by lower PGS, achieve disproportionately greater metabolic gain from FOS. This metabolic gain likely results from their inherently lower baseline circulating omega-3 concentrations relative to the population average, leaving more room for dietary omega-3 intake to exert its effects. While individuals’ baseline circulating omega-3 concentrations increase with higher PGS, there is less room for dietary intake to further increase them. These observations suggest the presence of saturation in circulating omega-3 concentrations and thus diminishing returns of increasing PGS and dietary intake.

On the other hand, we found that FOS exhibits a broad attenuating effect on the polygenic predisposition to higher circulating omega-3 concentrations, suggesting a dynamic balance between external dietary intake and internal genetic contribution. In the absence of external FOS or OFI, internal biological mechanisms activate the fatty acid synthesis and long-chain PUFA metabolism pathways to maintain necessary levels. In contrast, external fish or fish oil enriched in PUFAs acts as transcriptional suppressors, inhibiting fatty acid-regulated transcription factors, such as *SREBP-1* [11, 40]. This process turns off internal synthesis pathways and attenuates genetic influence by inhibiting the expression of genes associated with lipogenesis (*FASN*), fatty acid synthesis, desaturation (*SCD1*, *FADS1/2*), and elongation (*ELOVL5/6*) [11]. Consequently, dietary omega-3 intake can maintain circulating concentrations sufficient to meet metabolic demands, effectively bypassing the requirement for internal synthesis. The observed interactions between PGS and FOS suggest a homeostatic mechanism where the body actively maintains metabolic stability once a specific physiological threshold is reached. Further longitudinal or functional studies are necessary to test this hypothesis and elucidate the underlying regulatory mechanisms.

In our study, the estimated effects of FOS on increasing circulating omega-3 fatty acid levels agree with findings from major randomized clinical trials. The STRENGTH trial reported that daily supplementation with 4 g of a carboxylic acid formulation of EPA and DHA over 12 months increased plasma DHA concentrations from 61.9 μg/ml to 90.7 μg/ml [12]. This equates to a median plasma increase of 7.2 μg/ml (0.0219 mmol/L, based on a DHA molar mass of 328.5 g/mol) per 1 g of omega-3 supplementation, which is consistent with our estimate of 0.027 mmol/L (**Supplementary Table 4**). Regarding relative percentages, the VITAL clinical trial found that daily intake of 1 g of fish oil capsule (containing 460 mg of EPA and 380 mg of DHA) for a year increased the mean plasma omega-3 index from 2.7% to 4.1%, corresponding to a 54.7% relative increase [13]. Similarly, the PISCES clinical trial observed that a 4 g/d of FOS (containing 1.6 g of EPA and 0.8 g of DHA) over three months increased median circulating levels from 3.44% to 4.24% for DHA, and from 6.17% to 8.57% for total omega-3 fatty acids [4]. This indicates that each 1 g of omega-3 supplementation daily is associated with a median increase of 20% for DHA and 60% for total omega-3 fatty acids. In our study, the association effect of FOS was 0.208 mmol/L (95% CI: 0.203 - 0.212) for DHA%, and 0.536 mmol/L (95% CI: 0.527 - 0.546) for Omega-3%, representing relative increases of 20.8% and 53.7%, respectively, compared to the non-FOS group **(Supplementary Table 4**). These estimates closely align with those reported in randomized clinical trials.

The efficacy of FOS in raising circulating omega-3 fatty acid concentrations exceeds the contribution of polygenic predisposition captured by PGS. In this study, FOS was associated with a 0.027 mmol/L increase in DHA levels, compared with a 0.017 mmol/L increase per 1 SD increase in PGS. In other words, taking FOS is equivalent to a 1.6-SD increment in PGS. Similarly, FOS was associated with a 0.068 mmol/L increase in Omega-3, whereas a 1 SD increase in PGS was associated with a 0.056 mmol/L increase. Taking FOS is equivalent to a 1.2-SD increment in PGS. These results suggest that external FOS or dietary fish intake may modulate omega-3 fatty acid profiles to a greater extent than genetic background alone.

We did not detect significant PGS-by-FOS interaction signals in AFR and EAS populations, likely due to the combined effects of the limited PGS transferability in non-EUR ancestries and small sample sizes. First, the limited cross-ancestry transferability of PGS, which were constructed primarily from European cohorts, may not accurately capture genetic variance in AFR and EAS populations. Consistent with this, we did not observe significant associations of PGS with observed concentrations of DHA% in AFR and EAS, nor with Omega-3% in AFR. Among the significant results, the association signals were markedly weaker in AFR and EAS populations compared with EUR and CSA, reflecting the well-documented reduced portability of PGS in non-EUR populations. Second, limited sample sizes (N = 6,323 in AFR, N = 2,571 in EAS) further constrained the statistical power to detect subtle interaction effects. Our power calculations indicate that a sample size of ∼25,000 is necessary to capture an interaction effect of 0.04 with 80% power at a significance threshold of 0.05 (**Supplementary Figure 14**). As expected, the required sample size increases substantially when aiming to detect smaller interaction effects. Further studies with larger sample sizes across these diverse ancestries are needed to investigate finer-scale differences.

Our study has several notable strengths. To the best of our knowledge, it is the first PGS-by-FOS study to explore how our polygenic predisposition interacts with the habitual FOS or OFI to influence the circulating omega-3 concentrations. Unlike single-gene or single-variant studies, our study offers a broader perspective by identifying specific genetic subgroups that achieve disproportionally greater metabolic gains from FOS and OFI, supporting the development of precision nutritional guidelines. Second, the sample size in our study is large (N = 416,315 in EUR), providing sufficient power to detect PGS-by-FOS and PGS-by-OFI interaction signals. Third, our cross-ancestry analysis reveals a common interaction pattern across both EUR and CSA populations, rather than an ancestry-specific pattern. Finally, we demonstrated the robustness of our findings through various replication and sensitivity analyses. Specifically, we replicated results using OFI as a dietary exposure and an additional European sample. We validated findings by applying two distinct PGS approaches, comparing raw and transformed phenotypes, and employing two different questionnaires to define FOS exposure.

Nonetheless, this study has some limitations. First, the limited sample sizes in non-EUR populations restricted the statistical power to detect interaction signals within those groups. Second, while we employed two PGS approaches and both captured portions of the phenotypic variance in observed circulating omega-3 concentrations, the utilization of cross-ancestry PGS methods or models incorporating interaction effects could further enhance the PGS predictive performance. Additionally, because the UKB dietary questionnaires use categorical survey questions and thus lack quantitative dosage information, both FOS and OFI were analyzed as binary exposures. Further research with detailed quantitative data may better dissect the interplay between dietary omega-3 intake and polygenic predisposition.

In conclusion, this study reveals that PGS and FOS interact to influence the circulating omega-3 fatty acid concentrations, including DHA, DHA%, Omega-3, and Omega-3%, in the EUR and CSA populations. PGS can identify specific genetic subgroups, among which individuals with lower PGS can experience disproportionately greater increases in circulating omega-3 concentrations in response to FOS. Our findings support the development of personalized dietary recommendations in order to maximize the health benefits of dietary omega-3 fatty acids. Specifically, those with lower genetic capacity (i.e., lower PGS) are expected to experience greater increases in circulating omega-3 concentrations following dietary intervention.

## Abbreviations

AFR: African
BMI: body mass index
CI: confidence interval
CSA: Central/South Asian
DHA: docosahexaenoic acid or its absolute circulating concentration
DHA%: docosahexaenoic acid to total fatty acids
EAS: East Asian
EPA: eicosapentaenoic acid
EUR: European
FOS: fish oil supplementation
GWAS: genome-wide association study
GEI: gene-by-environment interaction
HDL-C: high-density lipoprotein cholesterol
RINT: rank-based inverse normal transformation
LD: linkage disequilibrium
NMR: nuclear magnetic resonance
OFI: oily fish intake
Omega-3: absolute circulating concentration of total omega-3 fatty acids
Omega-3%: total omega-3 fatty acids to total fatty acids
P_Int_: p-value testing the interaction between polygenic score and dietary exposures
P+T: pruning and thresholding
PGS: polygenic score
PUFAs: polyunsaturated fatty acids
QC: quality control
SD: standard deviation
SNP: single nucleotide polymorphism
TSI: Townsend index
UKB: UK Biobank.

## Acknowledgements

We would like to thank the UK Biobank participants and administrative staff. This research has been conducted using the UK Biobank resource under Application Number 48818. We also want to thank the UGA GACRC staff for facilitating our data analysis.

## Author contributions

KY and HX took responsibility for the integrity of the data and the accuracy of the data analysis; KY conceived and designed the project; KY and HX conducted research; HX analyzed data with assistance from GY, YL, HF, SS, YS, CWKC, and BFD; HX and KY drafted the manuscript, and all other authors critically revised the manuscript. All authors read and approved the final manuscript.

## Data availability

The PGS weights constructed using SBayesRC and P+T approaches are publicly available in the PGS Catalog (https://www.pgscatalog.org/) upon publication. The GWAS summary statistics for four circulating omega-3 fatty acids were downloaded from GWAS Catalog (https://www.ebi.ac.uk/gwas/, GCST90301955, GCST90301956, GCST90301959, GCST90301960).

## References

1. Li, Z.-H., et al., Associations of habitual fish oil supplementation with cardiovascular outcomes and all cause mortality: evidence from a large population based cohort study. bmj, 2020. 368.

2. Del Gobbo, L.C., et al., omega-3 Polyunsaturated Fatty Acid Biomarkers and Coronary Heart Disease: Pooling Project of 19 Cohort Studies. JAMA Intern Med, 2016. 176(8): p. 1155–66.

3. Djousse, L., et al., Fish consumption, omega-3 fatty acids and risk of heart failure: a meta-analysis. Clin Nutr, 2012. 31(6): p. 846–53.

4. Lok, C.E., et al., Fish-Oil Supplementation and Cardiovascular Events in Patients Receiving Hemodialysis. N Engl J Med, 2026. 394(2): p. 128–137.

5. Zheng, J.S., et al., Intake of fish and marine n-3 polyunsaturated fatty acids and risk of breast cancer: meta-analysis of data from 21 independent prospective cohort studies. BMJ, 2013. 346: p. f3706.

6. Wu, S., et al., Fish consumption and colorectal cancer risk in humans: a systematic review and meta-analysis. Am J Med, 2012. 125(6): p. 551–9 e5.

7. Dangour, A.D., et al., B-vitamins and fatty acids in the prevention and treatment of Alzheimer’s disease and dementia: a systematic review. J Alzheimers Dis, 2010. 22(1): p. 205–24.

8. Shinto, L.H., et al., omega-3 PUFA for Secondary Prevention of White Matter Lesions and Neuronal Integrity Breakdown in Older Adults: A Randomized Clinical Trial. JAMA Netw Open, 2024. 7(8): p. e2426872.

9. Bouwens, M., et al., Fish-oil supplementation induces antiinflammatory gene expression profiles in human blood mononuclear cells. Am J Clin Nutr, 2009. 90(2): p. 415–24.

10. Ahmadi, A.R., et al., Impact of omega-3 fatty acids supplementation on the gene expression of peroxisome proliferator activated receptors-γ, α and fibroblast growth factor-21 serum levels in patients with various presentation of metabolic conditions: a GRADE assessed systematic review and dose–response meta-analysis of clinical trials. Frontiers in Nutrition, 2023. 10: p. 1202688.

11. Jump, D.B., S. Tripathy, and C.M. Depner, Fatty acid-regulated transcription factors in the liver. Annu Rev Nutr, 2013. 33: p. 249–69.

12. Nicholls, S.J., et al., Effect of high-dose omega-3 fatty acids vs corn oil on major adverse cardiovascular events in patients at high cardiovascular risk: the STRENGTH randomized clinical trial. Jama, 2020. 324(22): p. 2268–2280.

13. Manson, J.E., et al., Marine n-3 Fatty Acids and Prevention of Cardiovascular Disease and Cancer. N Engl J Med, 2019. 380(1): p. 23–32.

14. Francis, M., et al., Fifty-one novel and replicated GWAS loci for polyunsaturated and monounsaturated fatty acids in 124,024 Europeans. medRxiv, 2022: p. 2022.05. 27.22275343.

15. Kettunen, J., et al., Genome-wide association study identifies multiple loci influencing human serum metabolite levels. Nat Genet, 2012. 44(3): p. 269–76.

16. Yang, C., et al., Genome-wide association studies and fine-mapping identify genomic loci for n-3 and n-6 polyunsaturated fatty acids in Hispanic American and African American cohorts. Commun Biol, 2023. 6(1): p. 852.

17. Sun, Y., H. Xu, and K. Ye, Genome-wide association studies and multi-omics integrative analysis reveal novel loci and their molecular mechanisms for circulating polyunsaturated, monounsaturated, and saturated fatty acids. Human Genetics and Genomics Advances, 2025: p. 100470.

18. Hu, Y., et al., Genome-wide meta-analyses identify novel loci associated with n-3 and n-6 polyunsaturated fatty acid levels in Chinese and European-ancestry populations. Hum Mol Genet, 2016. 25(6): p. 1215–24.

19. Juan, J., et al., Joint effects of fatty acid desaturase 1 polymorphisms and dietary polyunsaturated fatty acid intake on circulating fatty acid proportions. Am J Clin Nutr, 2018. 107(5): p. 826–833.

20. Tomaszewski, N., et al., Effect of APOE Genotype on Plasma Docosahexaenoic Acid (DHA), Eicosapentaenoic Acid, Arachidonic Acid, and Hippocampal Volume in the Alzheimer’s Disease Cooperative Study-Sponsored DHA Clinical Trial. J Alzheimers Dis, 2020. 74(3): p. 975–990.

21. Ihejirika, S.A., et al., A multi-level gene-diet interaction analysis of fish oil and 14 polyunsaturated fatty acid traits identifies the FADS and GPR12 loci. HGG Adv, 2025. 6(3): p. 100459.

22. Thomas, D., Methods for investigating gene-environment interactions in candidate pathway and genome-wide association studies. Annu Rev Public Health, 2010. 31: p. 21–36.

23. Ye, Y., et al., Interactions between enhanced polygenic risk scores and lifestyle for cardiovascular disease, diabetes, and lipid levels. Circulation: Genomic and Precision Medicine, 2021. 14(1): p. e003128.

24. Sun, Y., et al., Fish oil supplementation modifies the associations between genetically predicted and observed concentrations of blood lipids: a cross-sectional gene-diet interaction study in UK Biobank. The American Journal of Clinical Nutrition, 2024. 120(3): p. 540–549.

25. Sudlow, C., et al., UK biobank: an open access resource for identifying the causes of a wide range of complex diseases of middle and old age. PLoS Med, 2015. 12(3): p. e1001779.

26. Karczewski, K.J., et al., Pan-UK Biobank genome-wide association analyses enhance discovery and resolution of ancestry-enriched effects. Nature genetics, 2025. 57(10): p. 2408–2417.

27. Bycroft, C., et al., The UK Biobank resource with deep phenotyping and genomic data. Nature, 2018. 562(7726): p. 203-209.

28. Bradbury, K.E., et al., Dietary assessment in UK Biobank: an evaluation of the performance of the touchscreen dietary questionnaire. J Nutr Sci, 2018. 7: p. e6.

29. Liu, B., et al., Development and evaluation of the Oxford WebQ, a low-cost, web-based method for assessment of previous 24 h dietary intakes in large-scale prospective studies. Public Health Nutr, 2011. 14(11): p. 1998–2005.

30. Karjalainen, M.K., et al., Genome-wide characterization of circulating metabolic biomarkers. Nature, 2024. 628(8006): p. 130-138.

31. Murphy, A.E., B.M. Schilder, and N.G. Skene, MungeSumstats: A Bioconductor package for the standardisation and quality control of many GWAS summary statistics. Bioinformatics, 2021. 37(23): p. 4593–6.

32. Zheng, Z., et al., Leveraging functional genomic annotations and genome coverage to improve polygenic prediction of complex traits within and between ancestries. Nature Genetics, 2024. 56(5): p. 767–777.

33. Goldstein, B.A., et al., Contemporary Considerations for Constructing a Genetic Risk Score: An Empirical Approach. Genet Epidemiol, 2015. 39(6): p. 439–45.

34. Marquez-Luna, C., et al., Incorporating functional priors improves polygenic prediction accuracy in UK Biobank and 23andMe data sets. Nat Commun, 2021. 12(1): p. 6052.

35. Finucane, H.K., et al., Partitioning heritability by functional annotation using genome-wide association summary statistics. Nat Genet, 2015. 47(11): p. 1228–35.

36. Dearden, E.K., C.D. Lloyd, and M. Green, Exploring the histories of health and deprivation in Britain, 1971-2011. Health Place, 2020. 61: p. 102255.

37. Van Buuren, S. and K. Groothuis-Oudshoorn, mice: Multivariate imputation by chained equations in R. Journal of statistical software, 2011. 45: p. 1–67.

38. Zhang, D., M.D. Zhang, and C. generalized R-squared, Package ‘rsq’. R Package Ver, 2024. 4: p. 2.

39. Gaye, A., T.W. Burton, and P.R. Burton, ESPRESSO: taking into account assessment errors on outcome and exposures in power analysis for association studies. Bioinformatics, 2015. 31(16): p. 2691–6.

40. Kim, H.J., M. Takahashi, and O. Ezaki, Fish oil feeding decreases mature sterol regulatory element-binding protein 1 (SREBP-1) by down-regulation of SREBP-1c mRNA in mouse liver. A possible mechanism for down-regulation of lipogenic enzyme mRNAs. J Biol Chem, 1999. 274(36): p. 25892–8.

