## Supplementary Figures for "Polygenic predisposition modifies the associations of fish oil supplementation with circulating omega-3 fatty acids: a cross-sectional gene-diet interaction study in UK Biobank"

**Abbreviations**

AFR, African; BMI, body mass index; CI, confidence interval; CSA, Central/South Asian; DHA, docosahexaenoic acid or its absolute circulating concentration; DHA%, docosahexaenoic acid to total fatty acids; EAS, East Asian; EUR, European; FOS, fish oil supplementation; RINT, rank-based inverse normal transformation; OFI, oily fish intake; Omega-3, absolute circulating concentration of total omega-3 fatty acids; Omega-3%, omega-3 fatty acids to total fatty acids; PhysicalAct, physical activity; P_Int_, p-value testing the interaction between polygenic score and dietary exposures; P+T, pruning and thresholding; PGS, polygenic score; SE, standard error; SD, standard deviation; TSI, Townsend index; UKB, UK Biobank.

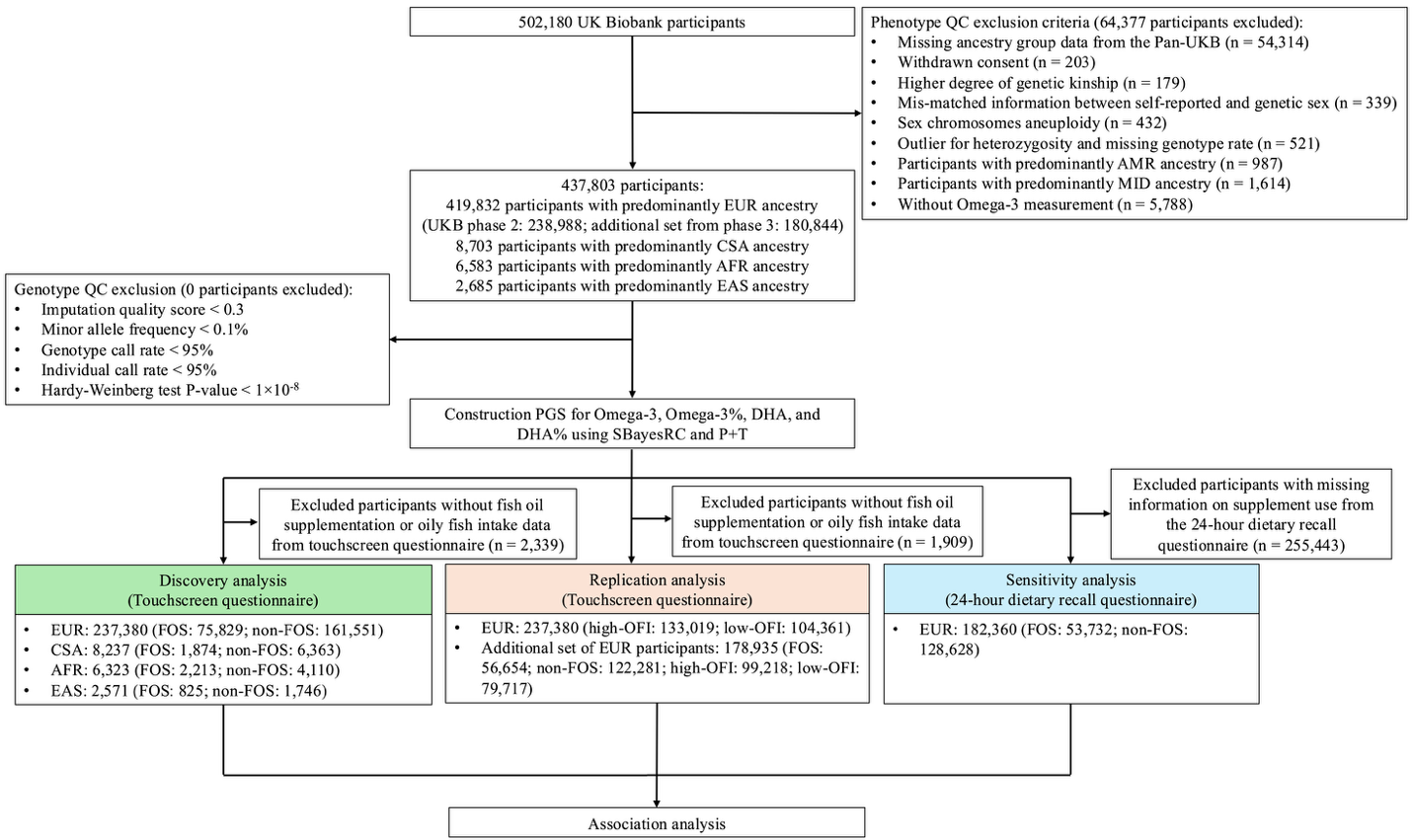

### **Supplementary Figure 1. Flowchart of study population selection in the UK Biobank**

Abbreviations: Omega-3, omega-3 fatty acids; Omega-3%, omega-3 fatty acids to total fatty acids percentage; DHA, docosahexaenoic acid; DHA%, docosahexaenoic acid to total fatty acids percentage; PGS, polygenic score; QC, quality control; EUR, European; EAS, East Asian; AFR, African; CSA, Central/South Asian; AMR, Admixed American; MID, Middle Eastern; UKB, UK Biobank.

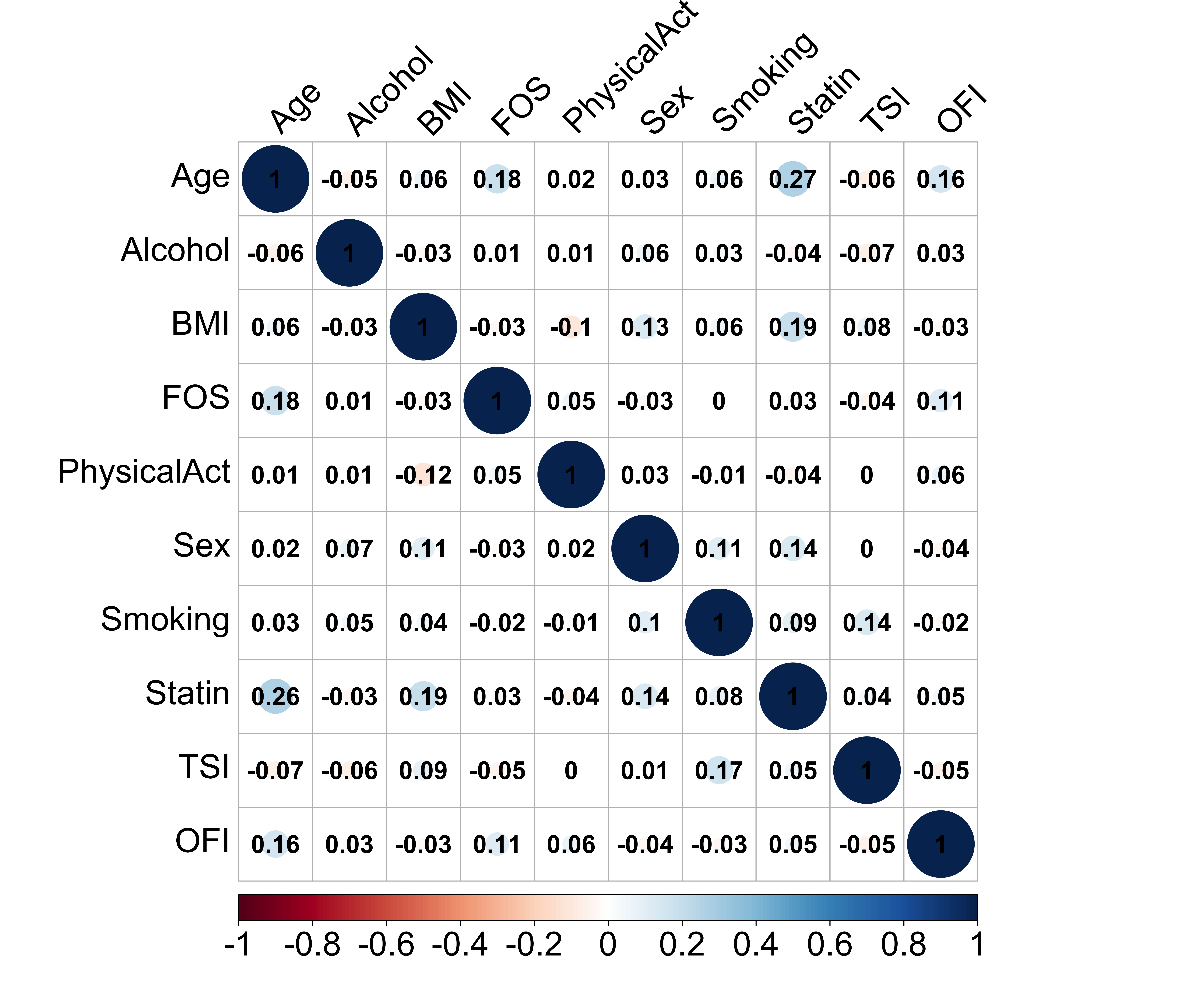

### **Supplementary Figure 2. Correlation between included study covariates**

The upper triangle shows the pairwise Spearman correlation coefficients, and the lower triangle shows the Pearson correlation coefficients. Color and number indicate the strength of the correlation.

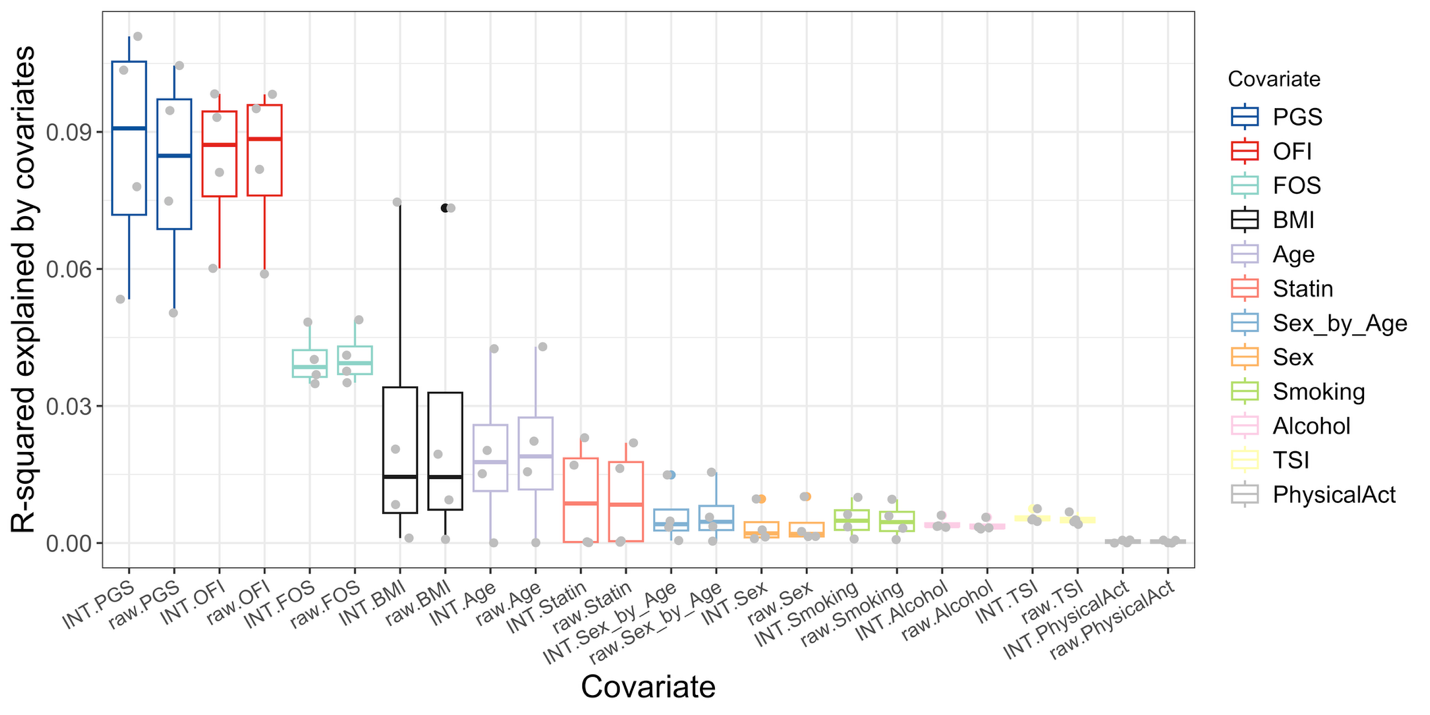

### **Supplementary Figure 3. Variance explained by covariates in circulating levels of four omega-3 fatty acid traits**

Association between twelve covariates and each of four omega-3 fatty acid traits (DHA, DHA%, Omega-3, and Oemga-3%) was tested, and the variance (partial R^2^) was estimated. Analyses were run on both the RINT-based and raw (i.e., untransformed) phenotypes. Results for both scales are displayed. Each dot represents a fatty acid trait. Abbreviations: BMI, body mass index; FOS, fish oil supplementation; OFI, oily fish intake; PhysicalAct, physical activity; PGS, polygenic score; TSI, Townsend index; RINT, rank-based inverse normal transformation.

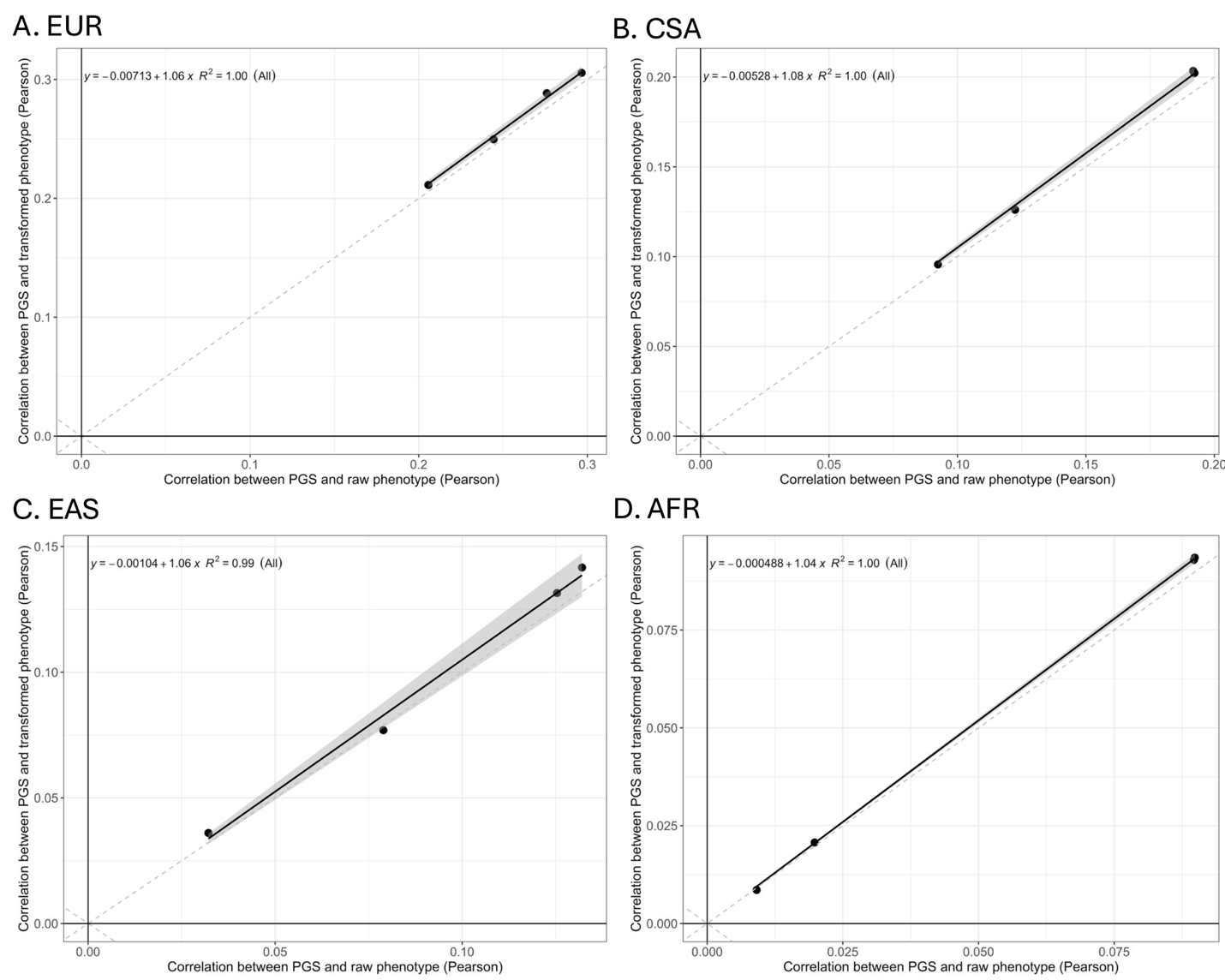

### **Supplementary Figure 4. Correlation of PGS with raw vs. RINT-based omega-3 fatty acid traits by ancestry**

(**A**) EUR, (**B**) CSA, (**C**) EAS and (**D**) AFR. Each data point represents one trait (DHA, DHA%, Omega-3, and Omega-3%). The x- and y-axis represent the Pearson correlation between SBayesRC-based PGS and the raw phenotype versus the rank-based inverse normal transformed phenotypes, respectively. The black line is the fitted line. R2 quantifies the correlation between x and y.

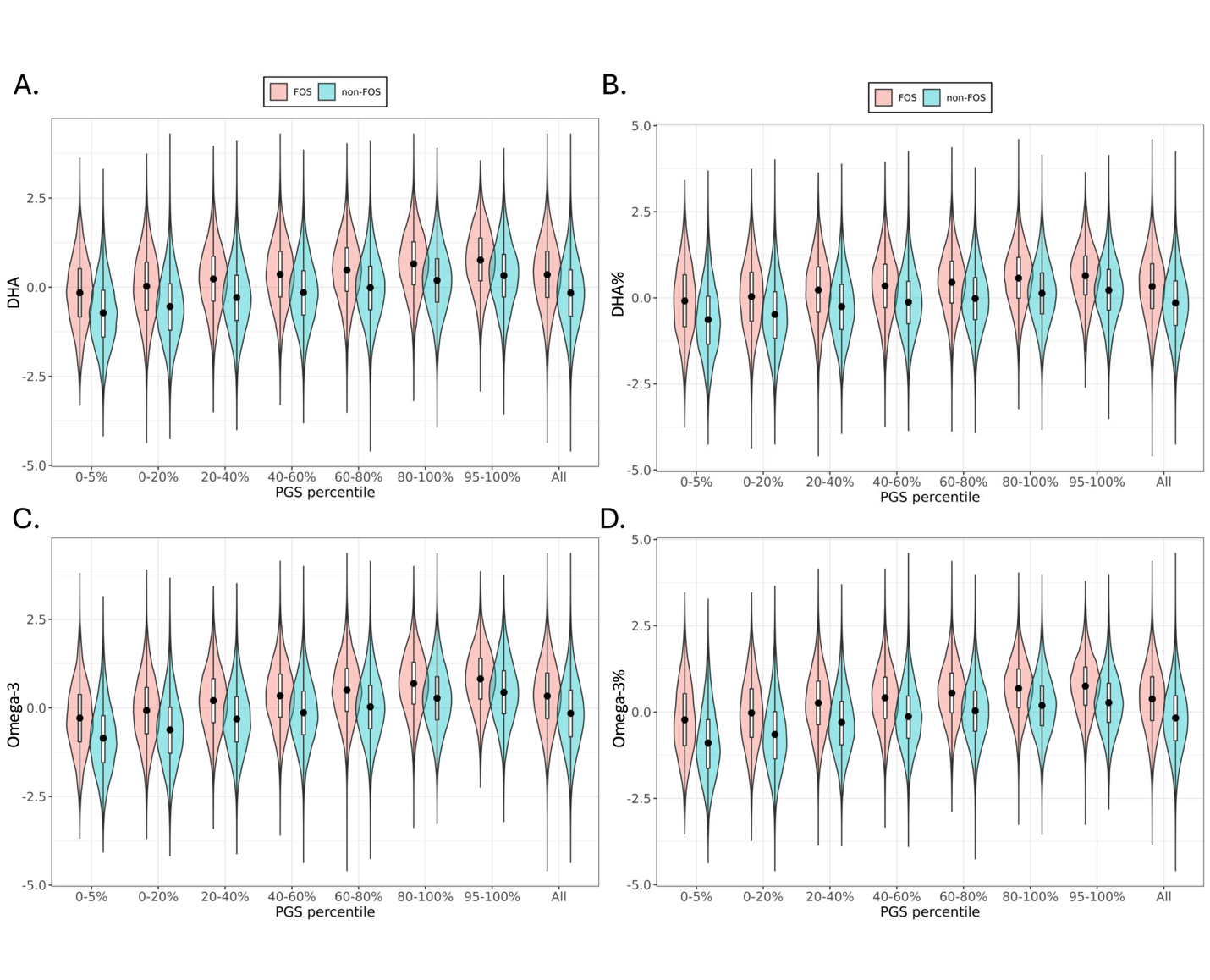

### **Supplementary Figure 5. Relationship between PGS with four observed circulating omega-3 fatty acid** **concentrations, stratified by FOS status and PGS subgroups**

Panels show results for **A)** DHA, **B)** DHA%, **C)** Omega-3, and **D)** Omega-3% based on participants of European ancestry (N = 237,380). The x-axis represents the PGS groups, while the y-axis displays the observed levels of four RINT-based phenotypes. PGS were categorized into seven groups based on their distribution: the bottom 5% (0–5%), five quintile-based bins (0–20%, 20–40%, 40–60%, 60–80%, and 80–100%), and the top 5% (95–100%). FOS users and non-FOS users are denoted as red and blue, respectively. Black dots indicate mean fatty acid levels within each subgroup. Box corresponds to 25–75% interval.

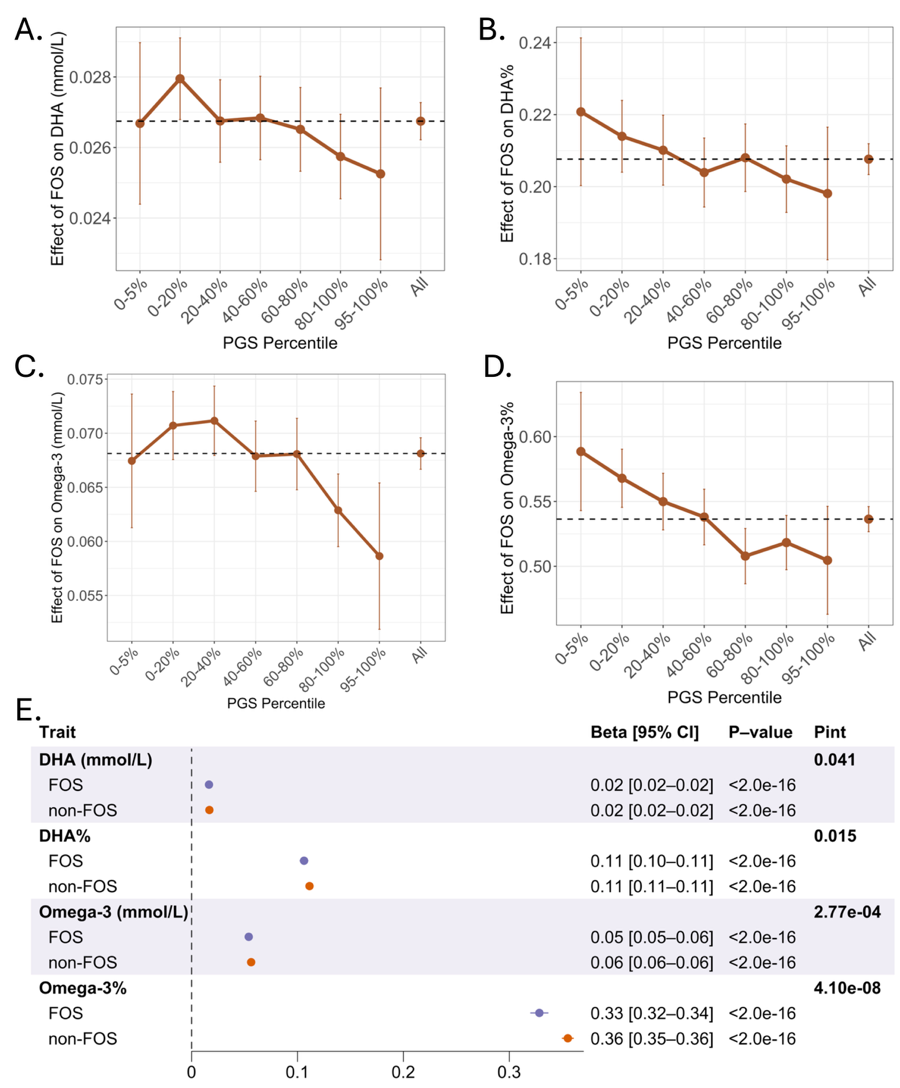

### **Supplementary Figure 6. Evidence of PGS-by-FOS interactions on raw circulating concentrations of omega-3 fatty acids in EUR participants**

**A) – D)** PGS modified the positive associations of FOS with raw circulating concentrations of DHA, DHA%, Omega-3, Omega-3%. We calculated PGS using the SBayesRC algorithm, and performed analyses on participants of EUR ancestry (N = 237,380). **E)** PGS was associated with a smaller increase in circulating omega-3 fatty acid concentrations among individuals in the FOS group compared to those in the non-FOS group. Estimated effect sizes of PGS are presented as beta [95% CI] in the forest plot. P-value denotes the significance of the association between PGS and omega-3 within each stratum, while P_Int_ represents the interaction p-value between PGS and FOS.

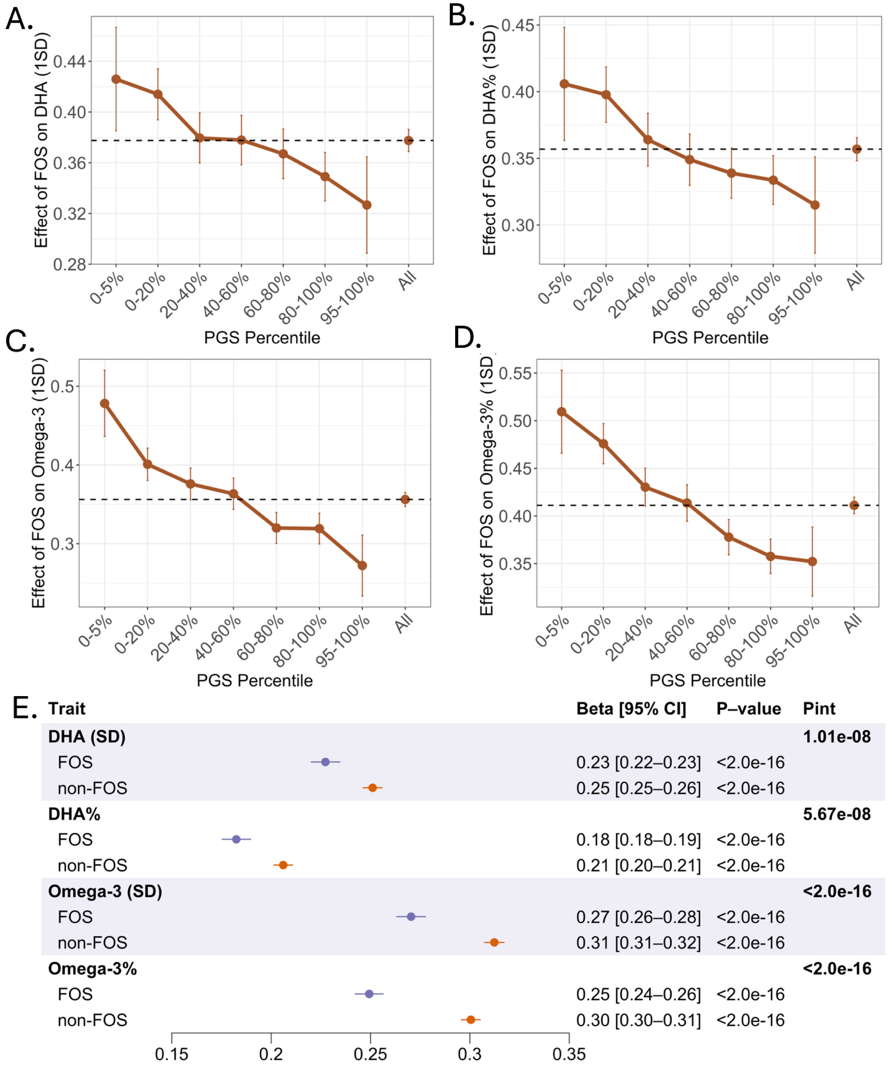

### **Supplementary Figure 7. Evidence of PGS-by-FOS interactions on RINT-based circulating concentrations of omega-3 fatty acids in EUR participants (Replication analysis)**

**A) – D)** PGS modified the positive associations of FOS with RINT-based circulating concentrations of DHA, DHA%, Omega-3, Omega-3%. We calculated PGS using the SBayesRC algorithm, and performed analyses on participants of EUR ancestry (N = 178,935). The touchscreen questionnaire was analyzed. **E)** PGS was associated with a smaller increase in circulating omega-3 fatty acid concentrations among individuals in the FOS group compared to those in the non-FOS group. Estimated effect sizes of PGS are presented as beta [95% CI] in the forest plot. P-value denotes the significance of the association between PGS and omega-3 fatty acids within each stratum, while P_Int_ represents the interaction p-value between PGS and FOS.

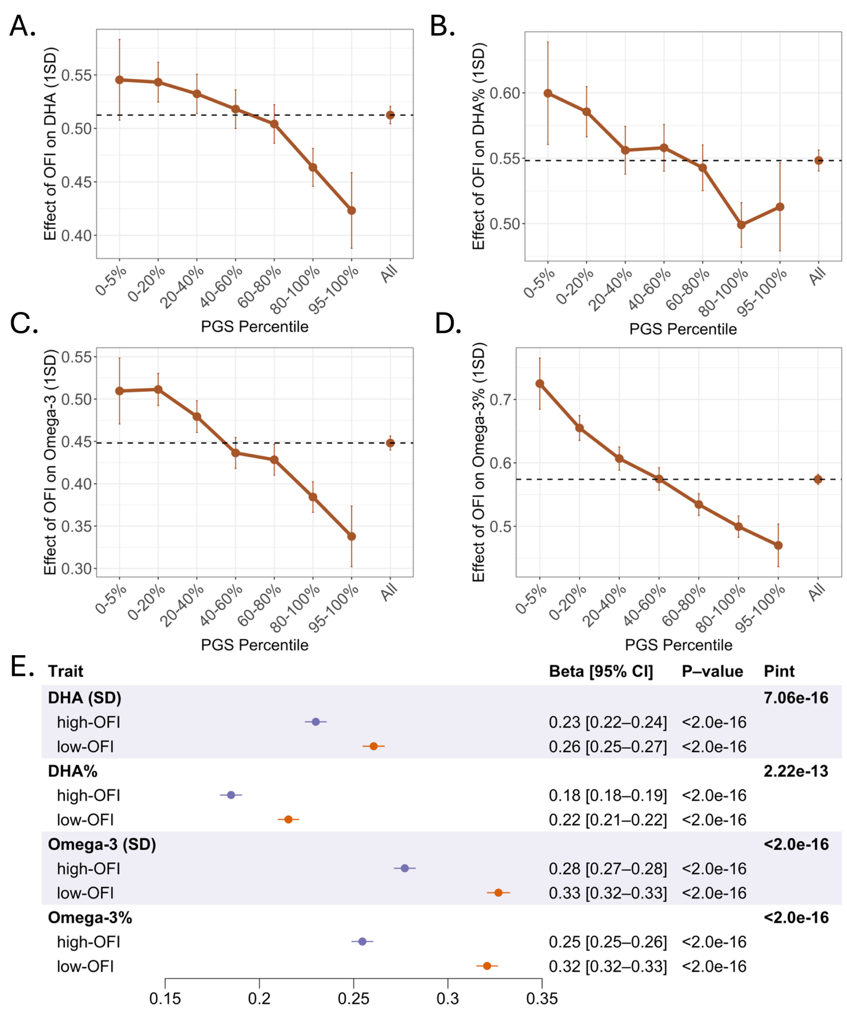

### **Supplementary Figure 8. Evidence of PGS-by-OFI interactions on RINT-based circulating concentrations of omega-3 fatty acids in EUR participants (Replication analysis)**

**A) – D)** PGS modified the positive associations of OFI with RINT-based circulating concentrations of DHA, DHA%, Omega-3, Omega-3%. We calculated PGS using the SBayesRC algorithm, and performed analyses on participants of EUR ancestry (N = 178,935). The touchscreen questionnaire was analyzed. **E)** PGS was associated with a smaller increase in circulating omega-3 fatty acid concentrations among individuals in the high-OFI group compared to those in the low-OFI group. Estimated effect sizes of PGS are presented as beta [95% CI] in the forest plot. P-value denotes the significance of the association between PGS and omega-3 fatty acids within each stratum, while P_Int_ represents the interaction p-value between PGS and OFI.

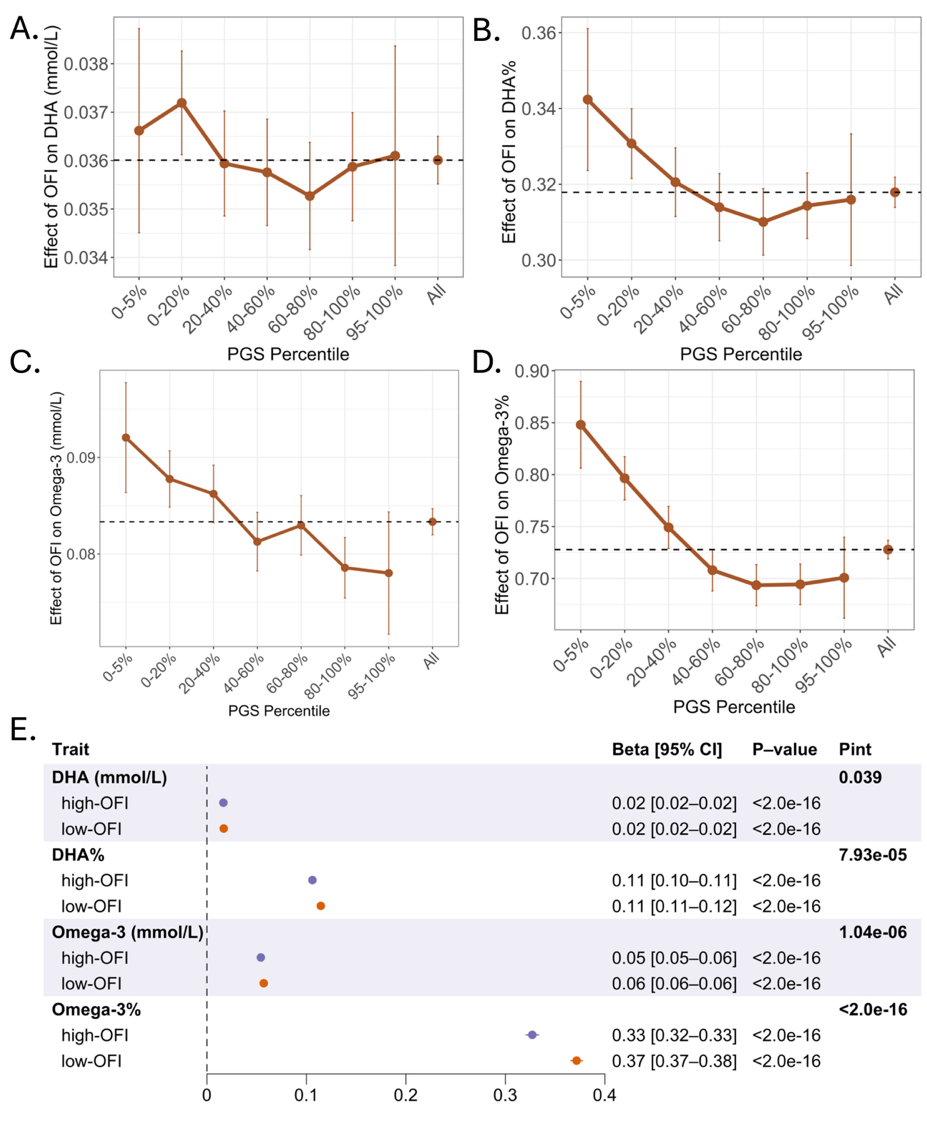

### **Supplementary Figure 9. Evidence of PGS-by-OFI interactions on raw circulating concentrations of omega-3 fatty acids in EUR participants**

**A) – D)** PGS modified the positive associations of OFI with raw circulating concentrations of DHA, DHA%, Omega-3, Omega-3%. We calculated PGS using the SBayesRC algorithm, and performed analyses on participants of EUR ancestry (N = 237,380). **E)** PGS was associated with a smaller increase in circulating omega-3 fatty acid levels among individuals in the high-OFI group compared to those in the low-OFI group. Estimated effect sizes of PGS are presented as beta [95% CI] in the forest plot. P-value denotes the significance of the association between PGS and omega-3 fatty acids within each stratum, while P_Int_ represents the interaction p-value between PGS and OFI.

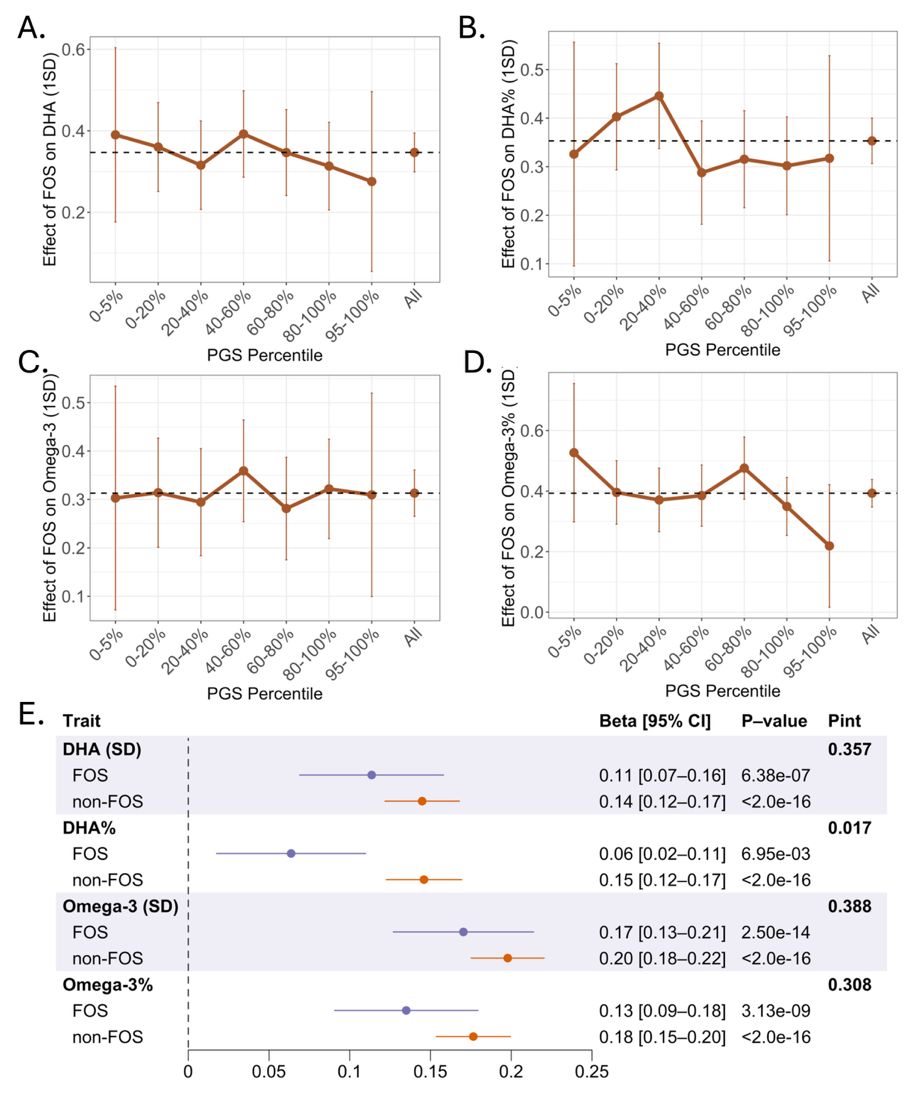

### **Supplementary Figure 10. Evidence of PGS-by-FOS interactions on RINT-based circulating** **concentration of DHA% in CSA participants**

**A) – D)** Changes in the association effects of FOS on RINT-based circulating concentrations of DHA, DHA%, omega-3, and omega-3% across PGS groups. We calculated PGS using the SBayesRC algorithm, and performed analyses on participants of CSA ancestry (N = 8,237). **E)** PGS was associated with a smaller increase in circulating DHA% among individuals in the FOS group compared to those in the non-FOS group. Estimated effect sizes of PGS are presented as beta [95% CI] in the forest plot. P-value denotes the significance of the association between PGS and omega-3 within each stratum, while P_Int_ represents the interaction p-value between PGS and FOS.

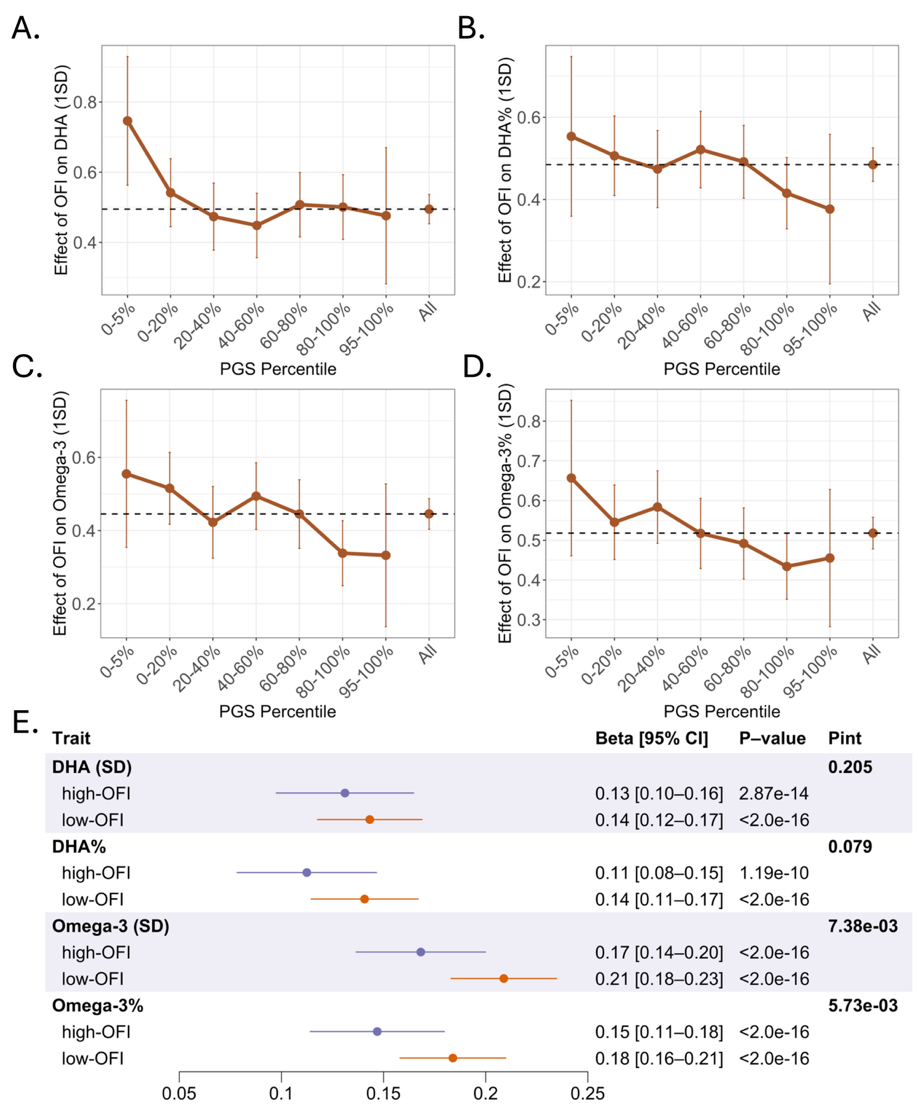

### **Supplementary Figure 11. Evidence of PGS-by-OFI interactions on RINT-based circulating concentrations of Omega-3 and Omega-3% in CSA participants**

**A) – D)** Changes in the association effects of high-OFI on RINT-based circulating concentrations of DHA, DHA%, omega-3, and omega-3% across PGS groups. We calculated PGS using the SBayesRC algorithm, and performed analyses on participants of CSA ancestry (N = 8,237). **E)** PGS was associated with a smaller increase in circulating omega-3 fatty acid concentrations among individuals in the high-OFI group compared to those in the lw-OFI group. Estimated effect sizes of PGS are presented as beta [95% CI] in the forest plot. P-value denotes the significance of the association between PGS and omega-3 fatty acids within each stratum, while P_Int_ represents the interaction p-value between PGS and OFI.

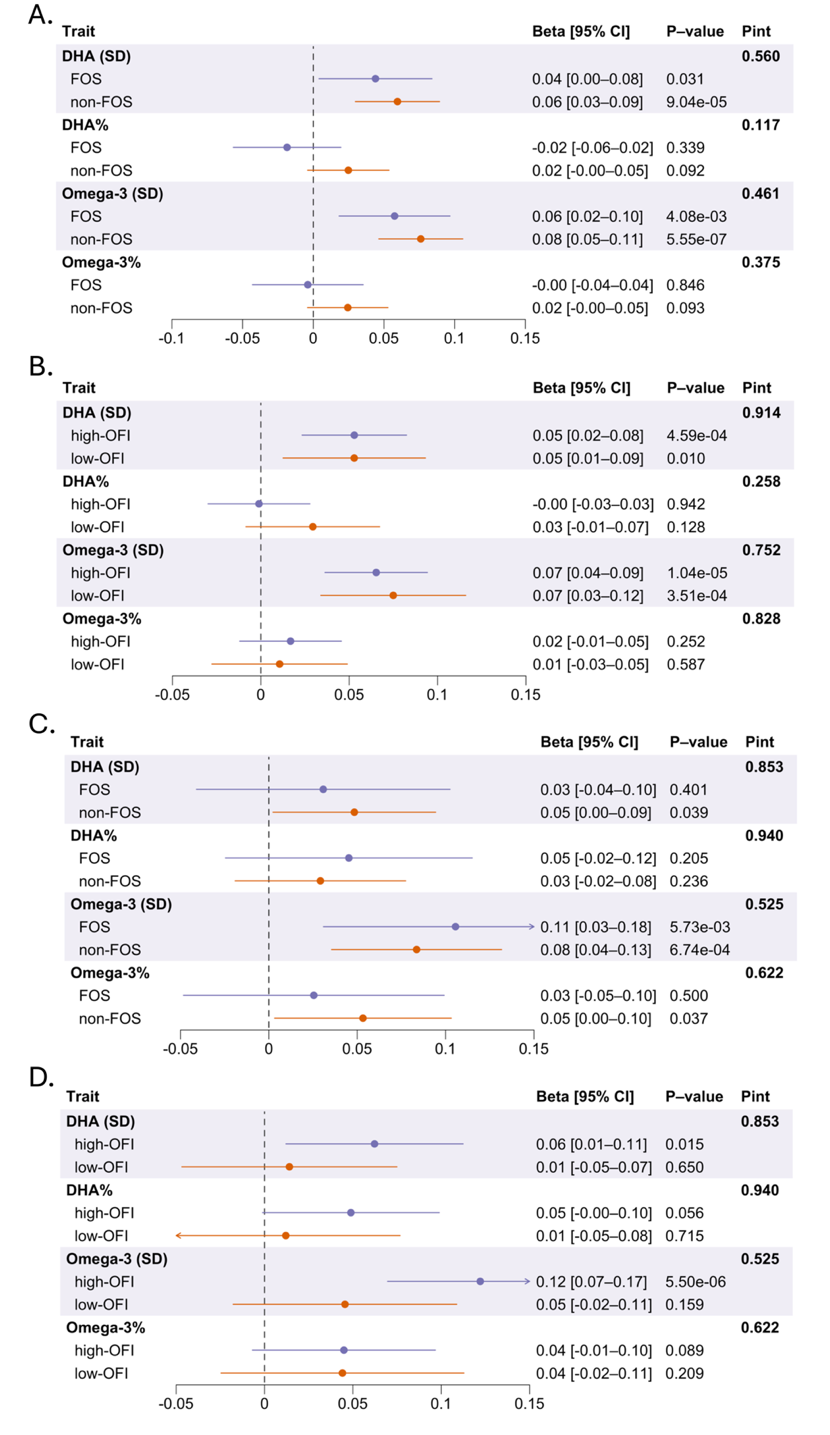

### **Supplementary Figure 12. Absence of PGS-by-FOS and PGS-by-OFI interactions on RINT-based circulating concentrations of omega-3 fatty acids in AFR and EAS participants**

The association between PGS and RINT-based circulating omega-3 concentrations did not significantly differ by dietary exposures across ancestry groups: **A)** FOS vs. non-FOS groups in AFR ancestry, **B)** high-OFI vs. low-OFI groups in AFR ancestry, **C)** FOS vs. non-FOS groups in EAS ancestry, and **D)** high-OFI vs. low-OFI groups in EAS ancestry. We calculated PGS using the SBayesRC algorithm, and performed analyses on participants of AFR (N = 6,323) and EAS (N = 2,571) ancestries, respectively. Estimated effect sizes of PGS are presented as beta [95% CI] in the forest plot. P-value denotes the significance of the association between PGS and omega-3 fatty acids within each stratum, while P_Int_ represents the interaction p-value between PGS and dietary exposures.

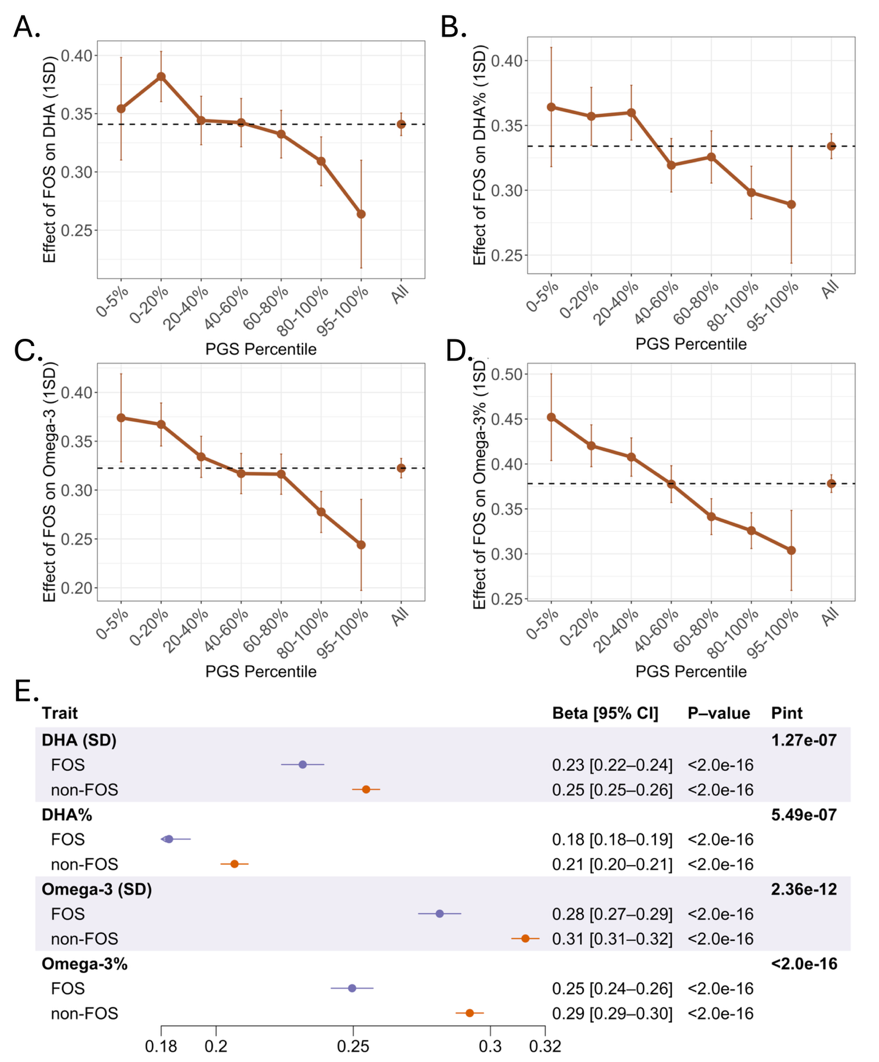

### **Supplementary Figure 13. Evidence of PGS-by-FOS interactions on RINT-based circulating concentrations of omega-3 fatty acids in EUR participants using a 24-hour dietary recall questionnaire**

**A) – D)** PGS modified the positive associations of FOS with RINT-based circulating concentrations of DHA, DHA%, Omega-3, Omega-3%. We calculated PGS using the SBayesRC algorithm, and performed analyses on participants of EUR ancestry from all UKB phases (N = 182,360). The 24-hour dietary recall questionnaire was analyzed. **E)** PGS was associated with a smaller increase in circulating omega-3 fatty acid concentrations among individuals in the FOS group compared to those in the non-FOS group. Estimated effect sizes of PGS are presented as beta [95% CI] in the forest plot. P-value denotes the significance of the association between PGS and omega-3 fatty acids within each stratum, while P_Int_ represents the interaction p-value between PGS and FOS.

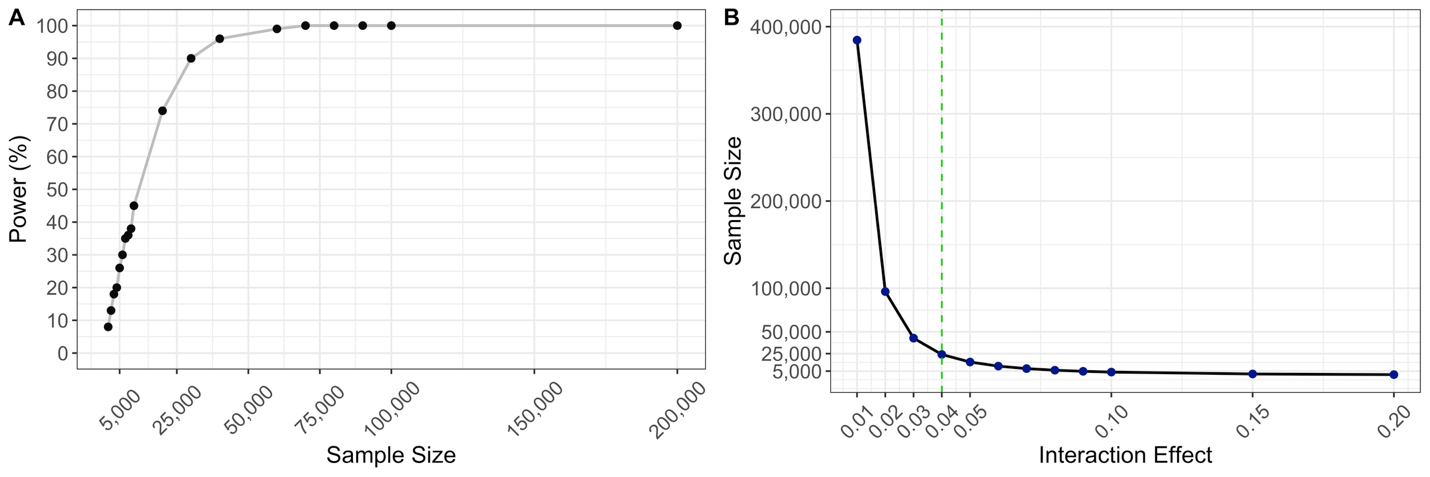

### **Supplementary Figure 14. Power calculation for the detection of PGS-by-FOS interactions**

**A)** Estimated sample sizes required to detect a fixed interaction effect of 0.04 across a gradient of statistical power (P < 0.05). **B)** Minimum sample size required to achieve 80% power across a range of interaction effect magnitudes.
