## Supplementary Tables for "Polygenic predisposition modifies the associations of fish oil supplementation with circulating omega-3 fatty acids: a cross-sectional gene-diet interaction study in UK Biobank"

### **Supplementary Table 1.** Participant characteristics stratified by FOS status

|  | **EUR (Discovery analysis)** | | **EUR (Replication analysis)** | | **CSA** | | **AFR** | | **EAS** | |
| --- | --- | --- | --- | --- | --- | --- | --- | --- | --- | --- |
|  | FOS  (N = 75,829) | non-FOS  (N = 161,551) | FOS  (N = 56,654) | non-FOS  (N = 122,281) | FOS  (N = 1,874) | non-FOS  (N = 6,363) | FOS  (N = 2,213) | non-FOS  (N = 4,110) | FOS  (N = 825) | non-FOS  (N = 1,746) |
| Age, y (SD) | 59 (7.3) | 56 (8.1) | 58.95 (7.32) | 55.83 (8.15) | 55.46 (8.36) | 52.92 (8.43) | 54.11 (8.2) | 51.45 (7.88) | 54.22 (7.89) | 51.28 (7.67) |
| Sex, female (%) | 42,386 (55.9) | 85,682 (53.0) | 24,503 (43.25) | 56,973 (46.59) | 932 (49.73) | 3,484 (54.75) | 766 (34.61) | 1,770 (43.07) | 231 (28.00) | 632 (36.20) |
| BMI, kg/m2 (SD) | 27 (4.5) | 28 (4.9) | 27.13 (4.54) | 27.45 (4.86) | 26.92 (4.24) | 27.26 (4.45) | 29.55 (5.37) | 29.72 (5.49) | 24.20 (3.48) | 24.76 (3.85) |
| Omega-3, mmol/L (SD) | 0.61 (0.24) | 0.50 (0.20) | 0.62 (0.24) | 0.51 (0.21) | 0.56 (0.25) | 0.46 (0.21) | 0.63 (0.25) | 0.57 (0.24) | 0.62 (0.29) | 0.53 (0.25) |
| Omega-3% (SD) | 4.95 (1.62) | 4.09 (1.40) | 4.91 (1.57) | 4.10 (1.38) | 4.55 (1.66) | 3.73 (1.46) | 5.90 (1.95) | 5.44 (1.91) | 5.01 (1.98) | 4.38 (1.78) |
| DHA, mmol/L (SD) | 0.27 (0.09) | 0.22 (0.08) | 0.27 (0.09) | 0.23 (0.08) | 0.24 (0.09) | 0.20 (0.08) | 0.29 (0.10) | 0.27 (0.09) | 0.28 (0.11) | 0.24 (0.10) |
| DHA% (SD) | 2.21 (0.70) | 1.88 (0.62) | 2.20 (0.70) | 1.88 (0.63) | 2.02 (0.72) | 1.68 (0.63) | 2.76 (0.79) | 2.58 (0.78) | 2.32 (0.86) | 2.06 (0.78) |
| Smoking status (%) | | | | | | | | | | |
| Never | 40,461 (53.4) | 88,331 (54.7) | 30,402 (53.66) | 66,796 (54.63) | 1,445 (77.11) | 5,013 (78.78) | 1,603 (72.44) | 2,858 (69.54) | 633 (76.73) | 1,288 (73.77) |
| Previous | 29,392 (38.8) | 54,814 (33.9) | 21,854 (38.57) | 41,646 (34.06) | 277 (14.78) | 766 (12.04) | 365 (16.49) | 737 (17.93) | 132 (16.00) | 302 (17.30) |
| Current | 5,976 (7.9) | 18,406 (11.4) | 4,398 (7.76) | 13,839 (11.32) | 152 (8.11) | 584 (9.18) | 245 (11.07) | 515 (12.53) | 60 (7.27) | 156 (8.93) |
| Alcohol status (%) | | | | | | | | | | |
| Never | 2,324 (3.1) | 5,049 (3.1) | 1,686 (2.98) | 3,946 (3.23) | 538 (28.71) | 2,763 (43.42) | 297 (13.42) | 663 (16.13) | 167 (20.24) | 365 (20.90) |
| Previous | 2,360 (3.1) | 5,822 (3.6) | 1,860 (3.28) | 4,335 (3.55) | 103 (5.50) | 357 (5.61) | 97 (4.38) | 234 (5.69) | 34 (4.12) | 81 (4.64) |
| Current | 71,145 (93.8) | 150,680 (93.3) | 53,108 (93.74) | 114,000 (93.23) | 1,233 (65.80) | 3,243 (50.97) | 1,819 (82.20) | 3,213 (78.18) | 624 (75.64) | 1,300 (74.46) |
| Physical activity (%) | | | | | | | | | | |
| Low | 12,000 (15.8) | 31,714 (19.6) | 8,878 (15.67) | 23,499 (19.22) | 371 (19.80) | 1,605 (25.22) | 376 (16.99) | 880 (21.41) | 140 (16.97) | 347 (19.87) |
| Moderate | 30,321 (40.0) | 66,182 (41.0) | 22,946 (40.50) | 50,871 (41.60) | 719 (38.37) | 2,530 (39.76) | 801 (36.20) | 1,594 (38.78) | 325 (39.39) | 729 (41.75) |
| High | 33,508 (44.2) | 63,655 (39.4) | 24,830 (43.83) | 47,911 (39.18) | 784 (41.84) | 2,228 (35.01) | 1,036 (46.81) | 1,636 (39.81) | 360 (43.64) | 670 (38.37) |
| Statin use, yes (%) | 13,923 (18.4) | 25,890 (16.0) | 9,754 (17.22) | 18,582 (15.20) | 560 (29.88) | 1,571 (24.69) | 337 (15.23) | 602 (14.65) | 92 (11.15) | 175 (10.02) |

Values are reported as numbers (%) for categorical variables and mean (SD) for continuous variables. Abbreviations: BMI, body mass index; Omega-3, omega-3 fatty acids; Omega-3%, omega-3 fatty acids to total fatty acids percentage; DHA, docosahexaenoic acid; DHA%, docosahexaenoic acid to total fatty acids percentage; SD, standard deviation; EUR, European; CSA, Central/South Asian; AFR, African; EAS, East Asian; FOS, fish oil supplementation user; non-FOS, non-fish oil supplementation user.

### **Supplementary Table 2.** Participant characteristics stratified by OFI status

|  | **EUR (Discovery analysis)** | | **EUR (Replication analysis)** | | **CSA** | | **AFR** | | **EAS** | |
| --- | --- | --- | --- | --- | --- | --- | --- | --- | --- | --- |
|  | high-OFI  (N = 133,019) | low-OFI  (N = 104,361) | high-OFI  (N = 99,218) | low-OFI  (N = 79,717) | high-OFI  (N = 3,382) | low-OFI  (N = 4,855) | high-OFI  (N = 4,267) | low-OFI  (N = 2,056) | high-OFI  (N = 1,593) | low-OFI  (N = 978) |
| Age, y (SD) | 57.99 (7.77) | 55.44 (8.07) | 57.93 (7.79) | 55.44 (8.10) | 53.61 (8.66) | 53.42 (8.35) | 53.26 (8.23) | 50.55 (7.50) | 52.75 (7.91) | 51.37 (7.72) |
| Sex, female (%) | 73,904 (55.6) | 54,164 (51.9) | 43,715 (44.06) | 37,761 (47.37) | 1,925 (56.92) | 2,491 (51.31) | 1,662 (38.95) | 874 (42.51) | 470 (29.50) | 393 (40.18) |
| BMI, kg/m2 (SD) | 27.30 (4.64) | 27.60 (4.91) | 27.21 (4.65) | 27.52 (4.89) | 27.12 (4.23) | 27.23 (4.53) | 29.74 (5.36) | 29.50 (5.62) | 24.52 (3.69) | 24.69 (3.84) |
| Omega-3, mmol/L (SD) | 0.59 (0.23) | 0.46 (0.18) | 0.60 (0.24) | 0.48 (0.19) | 0.57 (0.25) | 0.42 (0.18) | 0.63 (0.26) | 0.51 (0.21) | 0.61 (0.28) | 0.47 (0.22) |
| Omega-3% (SD) | 4.84 (1.61) | 3.77 (1.18) | 4.82 (1.57) | 3.78 (1.15) | 4.61 (1.71) | 3.44 (1.20) | 5.98 (2.00) | 4.82 (1.50) | 4.99 (1.93) | 3.92 (1.54) |
| DHA, mmol/L (SD) | 0.26 (0.09) | 0.21 (0.07) | 0.27 (0.09) | 0.21 (0.07) | 0.24 (0.10) | 0.19 (0.06) | 0.29 (0.10) | 0.24 (0.07) | 0.28 (0.11) | 0.22 (0.08) |
| DHA% (SD) | 2.18 (0.70) | 1.73 (0.52) | 2.18 (0.70) | 1.73 (0.53) | 2.04 (0.76) | 1.56 (0.52) | 2.80 (0.82) | 2.31 (0.61) | 2.32 (0.83) | 1.85 (0.69) |
| Smoking status (%) | | | | | | | | | | |
| Never | 72,575 (54.6) | 56,217 (53.9) | 54,310 (54.74) | 42,888 (53.80) | 2,532 (74.87) | 3,926 (80.87) | 3,075 (72.06) | 1,386 (67.41) | 1,201 (75.39) | 720 (73.62) |
| Previous | 48,779 (36.7) | 35,427 (33.9) | 36,287 (36.57) | 27,213 (34.14) | 506 (14.96) | 537 (11.06) | 713 (16.71) | 389 (18.92) | 266 (16.70) | 168 (17.18) |
| Current | 11,665 (8.8) | 12,717 (12.2) | 8,621 (8.69) | 9,616 (12.06) | 344 (10.17) | 392 (8.07) | 479 (11.23) | 281 (13.67) | 126 (7.91) | 90 (9.20) |
| Alcohol status (%) | | | | | | | | | | |
| Never | 3,744 (2.8) | 3,629 (3.5) | 2,786 (2.81) | 2,846 (3.57) | 1,086 (32.11) | 2,215 (45.62) | 638 (14.95) | 322 (15.66) | 330 (20.72) | 202 (20.65) |
| Previous | 4,030 (3.0) | 4,152 (4.0) | 3,032 (3.06) | 3,163 (3.97) | 158 (4.67) | 302 (6.22) | 219 (5.13) | 112 (5.45) | 59 (3.70) | 56 (5.73) |
| Current | 125,245 (94.2) | 96,580 (92.5) | 93,400 (94.14) | 73,708 (92.46) | 2,138 (63.22) | 2,338 (48.16) | 3,410 (79.92) | 1,622 (78.89) | 1,204 (75.58) | 720 (73.62) |
| Physical activity (%) | | | | | | | | | | |
| Low | 21,825 (16.4) | 21,889 (21.0) | 15,976 (16.10) | 16,401 (20.57) | 694 (20.52) | 1,282 (26.41) | 781 (18.30) | 475 (23.10) | 268 (16.82) | 219 (22.39) |
| Moderate | 53,891 (40.5) | 42,612 (40.8) | 40,744 (41.07) | 33,073 (41.49) | 1,286 (38.02) | 1,963 (40.43) | 1,600 (37.50) | 795 (38.67) | 632 (39.67) | 422 (43.15) |
| High | 57,303 (43.1) | 39,860 (38.2) | 42,498 (42.83) | 30,243 (37.94) | 1,402 (41.45) | 1,610 (33.16) | 1,886 (44.20) | 786 (38.23) | 693 (43.50) | 337 (34.46) |
| Statin use, yes (%) | 24,441 (18.4) | 15,372 (14.7) | 17,091 (17.23) | 11,245 (14.11) | 923 (27.29) | 1,208 (24.88) | 708 (16.59) | 231 (11.24) | 181 (11.36) | 86 (8.79) |

Values are reported as numbers (%) for categorical variables and mean (SD) for continuous variables. Abbreviations: BMI, body mass index; Omega-3, omega-3 fatty acids; Omega-3%, omega-3 fatty acids to total fatty acids percentage; DHA, docosahexaenoic acid; DHA%, docosahexaenoic acid to total fatty acids percentage; SD, standard deviation; EUR, European; CSA, Central/South Asian; AFR, African; EAS, East Asian; OFI, oily fish intake.

### **Supplementary Table 3.** Correlations between variables in EUR participants (discovery analysis)

| **Variable1** | **Variable2** | **Coef (Pearson)** | **P (Pearson)** | **Coef (Spearman)** | **P (Spearman)** |
| --- | --- | --- | --- | --- | --- |
| PGS_DHA% | BMI | -0.018 | < 2.220446e-16 | -0.01625 | 4.94E-15 |
| PGS_Omega-3 | BMI | -0.016 | 2E-14 | -0.01511 | 3.27E-13 |
| PGS_DHA | BMI | -0.014 | 8E-12 | -0.01318 | 2.23E-10 |
| PGS_Omega-3% | BMI | -0.014 | 5E-11 | -0.01316 | 2.38E-10 |
| PGS_DHA% | TSI | -0.009 | 1E-05 | -0.00882 | 2.14E-05 |
| PGS_Omega-3% | TSI | -0.006 | 2E-03 | -0.00593 | 4.30E-03 |
| PGS_DHA | TSI | -0.004 | 6E-02 | -0.00363 | 8.09E-02 |
| PGS_DHA% | Smoking | -0.003 | 1E-01 | -0.00150 | 4.70E-01 |
| PGS_Omega-3 | TSI | -0.003 | 1E-01 | -0.00269 | 1.95E-01 |
| PGS_Omega-3 | Age | -0.001 | 7E-01 | -0.00135 | 5.16E-01 |
| PGS_Omega-3% | Sex | -0.001 | 7E-01 | -0.00049 | 8.13E-01 |
| PGS_Omega-3% | sex-by-age | -0.001 | 7E-01 | -0.00041 | 8.44E-01 |
| PGS_Omega-3% | Smoking | -0.001 | 8E-01 | -0.00024 | 9.07E-01 |
| PGS_DHA | Age | 0.000 | 9E-01 | -0.00042 | 8.39E-01 |
| PGS_DHA% | Sex | 0.001 | 8E-01 | 0.00073 | 7.24E-01 |
| PGS_Omega-3% | Statin use | 0.001 | 7E-01 | 0.00106 | 6.10E-01 |
| PGS_DHA% | sex-by-age | 0.001 | 6E-01 | 0.00151 | 4.67E-01 |
| PGS_Omega-3% | Age | 0.001 | 6E-01 | 0.00134 | 5.19E-01 |
| PGS_DHA | Physical activity | 0.001 | 6E-01 | 0.00046 | 8.23E-01 |
| PGS_DHA% | Statin use | 0.002 | 4E-01 | 0.00181 | 3.82E-01 |
| PGS_Omega-3 | Sex | 0.002 | 3E-01 | 0.00187 | 3.67E-01 |
| PGS_Omega-3 | Smoking | 0.002 | 3E-01 | 0.00317 | 1.26E-01 |
| PGS_DHA | Smoking | 0.002 | 3E-01 | 0.00351 | 9.10E-02 |
| PGS_Omega-3 | sex-by-age | 0.003 | 2E-01 | 0.00237 | 2.54E-01 |
| PGS_Omega-3% | Alcohol use | 0.003 | 2E-01 | 0.00221 | 2.88E-01 |

Continued

| PGS_DHA% | Physical activity | 0.003 | 2E-01 | 0.00228 | 2.73E-01 |
| --- | --- | --- | --- | --- | --- |
| PGS_DHA | Sex | 0.003 | 2E-01 | 0.00242 | 2.43E-01 |
| PGS_Omega-3 | Alcohol use | 0.003 | 1E-01 | 0.00256 | 2.17E-01 |
| PGS_DHA | Alcohol use | 0.003 | 1E-01 | 0.00287 | 1.67E-01 |
| PGS_DHA | FOS | 0.003 | 1E-01 | 0.00299 | 1.50E-01 |
| PGS_Omega-3% | Physical activity | 0.003 | 1E-01 | 0.00330 | 1.12E-01 |
| PGS_DHA | sex-by-age | 0.003 | 1E-01 | 0.00316 | 1.28E-01 |
| PGS_DHA% | Alcohol use | 0.004 | 6E-02 | 0.00381 | 6.67E-02 |
| PGS_Omega-3 | Physical activity | 0.004 | 6E-02 | 0.00337 | 1.05E-01 |
| PGS_DHA% | Age | 0.004 | 4E-02 | 0.00442 | 3.32E-02 |
| PGS_Omega-3 | FOS | 0.005 | 2E-02 | 0.00489 | 1.83E-02 |
| PGS_Omega-3% | FOS | 0.008 | 1E-04 | 0.00745 | 3.35E-04 |
| PGS_Omega-3% | OFI | 0.009 | 3E-05 | 0.00808 | 1.04E-04 |
| PGS_DHA% | FOS | 0.010 | 2E-06 | 0.00916 | 1.02E-05 |
| PGS_Omega-3 | OFI | 0.014 | 4E-12 | 0.01378 | 3.39E-11 |
| PGS_DHA | OFI | 0.017 | < 2.22e-16 | 0.01637 | 3.70E-15 |
| PGS_DHA% | OFI | 0.021 | < 2.22e-16 | 0.02008 | < 2.22e-16 |
| PGS_DHA | Statin use | 0.063 | < 2.22e-16 | 0.06169 | < 2.22e-16 |
| PGS_Omega-3 | Statin use | 0.074 | < 2.22e-16 | 0.07226 | < 2.22e-16 |
| Sex | Age | 0.025 | < 2.22e-16 | 0.02912 | < 2.22e-16 |
| Sex | sex-by-age | 0.982 | < 2.22e-16 | 0.94021 | < 2.22e-16 |
| Sex | BMI | 0.107 | < 2.22e-16 | 0.12900 | < 2.22e-16 |
| Sex | TSI | 0.012 | 2E-09 | 0.00439 | 3.20E-02 |
| Sex | Smoking | 0.102 | < 2.22e-16 | 0.10570 | < 2.22e-16 |
| Sex | Alcohol use | 0.073 | < 2.22e-16 | 0.06110 | < 2.22e-16 |
| Sex | Physical activity | 0.022 | < 2.22e-16 | 0.02632 | < 2.22e-16 |
| Sex | Statin use | 0.137 | < 2.22e-16 | 0.13660 | < 2.22e-16 |

Continued

| Sex | FOS | -0.027 | < 2.22e-16 | -0.02671 | < 2.22e-16 |
| --- | --- | --- | --- | --- | --- |
| Sex | OFI | -0.036 | < 2.22e-16 | -0.03643 | < 2.22e-16 |
| Age | sex-by-age | 0.155 | < 2.22e-16 | 0.26162 | < 2.22e-16 |
| Age | BMI | 0.057 | < 2.22e-16 | 0.06353 | < 2.22e-16 |
| Age | TSI | -0.066 | < 2.22e-16 | -0.06015 | < 2.22e-16 |
| Age | Smoking | 0.034 | < 2.22e-16 | 0.06140 | < 2.22e-16 |
| Age | Alcohol use | -0.058 | < 2.22e-16 | -0.05343 | < 2.22e-16 |
| Age | Physical activity | 0.013 | 7E-11 | 0.01609 | 3.96E-15 |
| Age | Statin use | 0.264 | < 2.22e-16 | 0.26895 | < 2.22e-16 |
| Age | FOS | 0.182 | < 2.22e-16 | 0.18024 | < 2.22e-16 |
| Age | OFI | 0.158 | < 2.22e-16 | 0.15906 | < 2.22e-16 |
| sex-by-age | BMI | 0.108 | < 2.22e-16 | 0.12671 | < 2.22e-16 |
| sex-by-age | TSI | 0.002 | 3E-01 | -0.01258 | 8.02E-10 |
| sex-by-age | Smoking | 0.109 | < 2.22e-16 | 0.12484 | < 2.22e-16 |
| sex-by-age | Alcohol use | 0.069 | < 2.22e-16 | 0.05285 | < 2.22e-16 |
| sex-by-age | Physical activity | 0.021 | < 2.22e-16 | 0.02439 | < 2.22e-16 |
| sex-by-age | Statin use | 0.176 | < 2.22e-16 | 0.20398 | < 2.22e-16 |
| sex-by-age | FOS | -0.005 | 1E-02 | 0.01235 | 1.63E-09 |
| sex-by-age | OFI | -0.016 | 2E-15 | 0.00123 | 5.49E-01 |
| BMI | TSI | 0.087 | < 2.22e-16 | 0.07596 | < 2.22e-16 |
| BMI | Smoking | 0.040 | < 2.22e-16 | 0.05798 | < 2.22e-16 |
| BMI | Alcohol use | -0.033 | < 2.22e-16 | -0.03036 | < 2.22e-16 |
| BMI | Physical activity | -0.117 | < 2.22e-16 | -0.10493 | < 2.22e-16 |
| BMI | Statin use | 0.193 | < 2.22e-16 | 0.19294 | < 2.22e-16 |
| BMI | FOS | -0.034 | < 2.22e-16 | -0.03081 | < 2.22e-16 |
| BMI | OFI | -0.029 | < 2.22e-16 | -0.02689 | < 2.22e-16 |
| TSI | Smoking | 0.166 | < 2.22e-16 | 0.13784 | < 2.22e-16 |

Continued

| TSI | Alcohol use | -0.061 | < 2.22e-16 | -0.06758 | < 2.22e-16 |
| --- | --- | --- | --- | --- | --- |
| TSI | Physical activity | -0.003 | 1E-01 | 0.00261 | 2.02E-01 |
| TSI | Statin use | 0.051 | < 2.22e-16 | 0.04483 | < 2.22e-16 |
| TSI | FOS | -0.048 | < 2.22e-16 | -0.04318 | < 2.22e-16 |
| TSI | OFI | -0.053 | < 2.22e-16 | -0.05127 | < 2.22e-16 |
| Smoking | Alcohol use | 0.049 | < 2.22e-16 | 0.03079 | < 2.22e-16 |
| Smoking | Physical activity | -0.014 | 3E-12 | -0.00802 | 8.95E-05 |
| Smoking | Statin use | 0.081 | < 2.22e-16 | 0.09160 | < 2.22e-16 |
| Smoking | FOS | -0.016 | 3E-14 | -0.00412 | 4.43E-02 |
| Smoking | OFI | -0.030 | < 2.22e-16 | -0.02106 | < 2.22e-16 |
| Alcohol use | Physical activity | 0.013 | 7E-11 | 0.01312 | 1.47E-10 |
| Alcohol use | Statin use | -0.035 | < 2.22e-16 | -0.03848 | < 2.22e-16 |
| Alcohol use | FOS | 0.008 | 1E-04 | 0.01052 | 2.76E-07 |
| Alcohol use | OFI | 0.029 | < 2.22e-16 | 0.03217 | < 2.22e-16 |
| Physical activity | Statin use | -0.042 | < 2.22e-16 | -0.04149 | < 2.22e-16 |
| Physical activity | FOS | 0.054 | < 2.22e-16 | 0.05359 | < 2.22e-16 |
| Physical activity | OFI | 0.064 | < 2.22e-16 | 0.06202 | < 2.22e-16 |
| Statin use | FOS | 0.029 | < 2.22e-16 | 0.02900 | < 2.22e-16 |
| Statin use | OFI | 0.048 | < 2.22e-16 | 0.04842 | < 2.22e-16 |
| FOS | OFI | 0.115 | < 2.22e-16 | 0.11463 | < 2.22e-16 |
| DHA (Raw) | PGS-by-FOS | 0.130 | < 2.22e-16 | 0.11162 | < 2.22e-16 |
| DHA (Raw) | PGS-by-OFI | 0.176 | < 2.22e-16 | 0.16217 | < 2.22e-16 |
| DHA (Raw) | DHA (RINT) | 0.994 | < 2.22e-16 | 1.00000 | < 2.22e-16 |
| DHA (Raw) | PGS_DHA | 0.244 | < 2.22e-16 | 0.24391 | < 2.22e-16 |
| DHA% (Raw) | PGS-by-FOS | 0.110 | < 2.22e-16 | 0.10755 | < 2.22e-16 |
| DHA% (Raw) | PGS-by-OFI | 0.146 | < 2.22e-16 | 0.14873 | < 2.22e-16 |
| DHA% (Raw) | DHA% (RINT) | 0.997 | < 2.22e-16 | 1.00000 | < 2.22e-16 |

Continued

| DHA% (Raw) | PGS_DHA% | 0.206 | < 2.22e-16 | 0.20655 | < 2.22e-16 |
| --- | --- | --- | --- | --- | --- |
| Omega-3 (Raw) | PGS-by-FOS | 0.158 | < 2.22e-16 | 0.13856 | < 2.22e-16 |
| Omega-3 (Raw) | PGS-by-OFI | 0.212 | < 2.22e-16 | 0.19696 | < 2.22e-16 |
| Omega-3 (Raw) | Omega-3 (RINT) | 0.991 | < 2.22e-16 | 1.00000 | < 2.22e-16 |
| Omega-3 (Raw) | PGS_Omega-3 | 0.297 | < 2.22e-16 | 0.29605 | < 2.22e-16 |
| Omega-3% (Raw) | PGS-by-FOS | 0.145 | < 2.22e-16 | 0.14351 | < 2.22e-16 |
| Omega-3% (Raw) | PGS-by-OFI | 0.192 | < 2.22e-16 | 0.19813 | < 2.22e-16 |
| Omega-3% (Raw) | Omega-3% (RINT) | 0.994 | < 2.22e-16 | 1.00000 | < 2.22e-16 |
| Omega-3% (Raw) | PGS_Omega-3% | 0.276 | < 2.22e-16 | 0.27963 | < 2.22e-16 |
| DHA (RINT) | PGS-by-FOS | 0.126 | < 2.22e-16 | 0.11162 | < 2.22e-16 |
| DHA (RINT) | PGS-by-OFI | 0.172 | < 2.22e-16 | 0.16217 | < 2.22e-16 |
| DHA (RINT) | PGS_DHA | 0.250 | < 2.22e-16 | 0.24391 | < 2.22e-16 |
| DHA% (RINT) | PGS-by-FOS | 0.109 | < 2.22e-16 | 0.10755 | < 2.22e-16 |
| DHA% (RINT) | PGS-by-OFI | 0.146 | < 2.22e-16 | 0.14873 | < 2.22e-16 |
| DHA% (RINT) | PGS_DHA% | 0.211 | < 2.22e-16 | 0.20655 | < 2.22e-16 |
| Omega-3 (RINT) | PGS-by-FOS | 0.153 | < 2.22e-16 | 0.13856 | < 2.22e-16 |
| Omega-3 (RINT) | PGS-by-OFI | 0.208 | < 2.22e-16 | 0.19696 | < 2.22e-16 |
| Omega-3 (RINT) | PGS_Omega-3 | 0.306 | < 2.22e-16 | 0.29605 | < 2.22e-16 |
| Omega-3% (RINT) | PGS-by-FOS | 0.143 | < 2.22e-16 | 0.14351 | < 2.22e-16 |
| Omega-3% (RINT) | PGS-by-OFI | 0.192 | < 2.22e-16 | 0.19813 | < 2.22e-16 |
| Omega-3% (RINT) | PGS_Omega-3% | 0.288 | < 2.22e-16 | 0.27963 | < 2.22e-16 |
| PGS-by-FOS | Sex | 0.006 | 3E-03 | 0.00530 | 1.07E-02 |
| PGS-by-OFI | Sex | 0.006 | 8E-03 | 0.00416 | 4.59E-02 |
| PGS-by-FOS | Age | -0.001 | 6E-01 | 0.00183 | 3.77E-01 |
| PGS-by-OFI | Age | -0.003 | 1E-01 | -0.00043 | 8.35E-01 |
| PGS-by-FOS | sex-by-age | 0.006 | 3E-03 | 0.00540 | 9.32E-03 |
| PGS-by-OFI | sex-by-age | 0.005 | 9E-03 | 0.00401 | 5.44E-02 |

Continued

| PGS-by-FOS | BMI | -0.007 | 1E-03 | -0.00571 | 5.95E-03 |
| --- | --- | --- | --- | --- | --- |
| PGS-by-OFI | BMI | -0.008 | 6E-05 | -0.00785 | 1.62E-04 |
| PGS-by-FOS | TSI | 0.002 | 3E-01 | 0.00005 | 9.79E-01 |
| PGS-by-OFI | TSI | 0.001 | 6E-01 | 0.00163 | 4.33E-01 |
| PGS-by-FOS | Smoking | 0.005 | 2E-02 | 0.00421 | 4.25E-02 |
| PGS-by-OFI | Smoking | 0.003 | 2E-01 | 0.00319 | 1.26E-01 |
| PGS-by-FOS | Alcohol use | 0.000 | 1E+00 | -0.00059 | 7.75E-01 |
| PGS-by-OFI | Alcohol use | 0.004 | 4E-02 | 0.00438 | 3.53E-02 |
| PGS-by-FOS | Physical activity | 0.002 | 2E-01 | 0.00077 | 7.09E-01 |
| PGS-by-OFI | Physical activity | 0.001 | 7E-01 | 0.00059 | 7.78E-01 |
| PGS-by-FOS | Statin use | 0.044 | < 2.22e-16 | 0.03803 | < 2.22e-16 |
| PGS-by-OFI | Statin use | 0.052 | < 2.22e-16 | 0.04761 | < 2.22e-16 |
| PGS-by-FOS | FOS | 0.004 | 6E-02 | 0.01969 | < 2.22e-16 |
| PGS-by-OFI | FOS | -0.001 | 6E-01 | 0.00191 | 3.60E-01 |
| PGS-by-FOS | OFI | 0.006 | 7E-03 | 0.00724 | 5.02E-04 |
| PGS-by-OFI | OFI | 0.011 | 3E-07 | 0.02394 | < 2.22e-16 |
| DHA (Raw) | Sex | -0.247 | < 2.22e-16 | -0.25120 | < 2.22e-16 |
| DHA (Raw) | Age | 0.170 | < 2.22e-16 | 0.16199 | < 2.22e-16 |
| DHA (Raw) | sex-by-age | -0.227 | < 2.22e-16 | -0.20957 | < 2.22e-16 |
| DHA (Raw) | BMI | -0.164 | < 2.22e-16 | -0.15784 | < 2.22e-16 |
| DHA (Raw) | TSI | -0.116 | < 2.22e-16 | -0.11209 | < 2.22e-16 |
| DHA (Raw) | Smoking | -0.093 | < 2.22e-16 | -0.07720 | < 2.22e-16 |
| DHA (Raw) | Alcohol use | 0.054 | < 2.22e-16 | 0.06471 | < 2.22e-16 |
| DHA (Raw) | Physical activity | 0.034 | < 2.22e-16 | 0.02950 | < 2.22e-16 |
| DHA (Raw) | Statin use | -0.020 | < 2.22e-16 | -0.01721 | < 2.22e-16 |
| DHA (Raw) | FOS | 0.240 | < 2.22e-16 | 0.23934 | < 2.22e-16 |
| DHA (Raw) | OFI | 0.314 | < 2.22e-16 | 0.31540 | < 2.22e-16 |

Continued

| DHA% (Raw) | Sex | -0.208 | < 2.220446e-16 | -0.21238 | < 2.220446e-16 |
| --- | --- | --- | --- | --- | --- |
| DHA% (Raw) | Age | 0.110 | < 2.220446e-16 | 0.10918 | < 2.220446e-16 |
| DHA% (Raw) | sex-by-age | -0.186 | < 2.220446e-16 | -0.16523 | < 2.220446e-16 |
| DHA% (Raw) | BMI | -0.264 | < 2.220446e-16 | -0.26541 | < 2.220446e-16 |
| DHA% (Raw) | TSI | -0.123 | < 2.220446e-16 | -0.11828 | < 2.220446e-16 |
| DHA% (Raw) | Smoking | -0.120 | < 2.220446e-16 | -0.10431 | < 2.220446e-16 |
| DHA% (Raw) | Alcohol use | 0.050 | < 2.220446e-16 | 0.05859 | < 2.220446e-16 |
| DHA% (Raw) | Physical activity | 0.047 | < 2.220446e-16 | 0.04321 | < 2.220446e-16 |
| DHA% (Raw) | Statin use | 0.058 | < 2.220446e-16 | 0.06117 | < 2.220446e-16 |
| DHA% (Raw) | FOS | 0.224 | < 2.220446e-16 | 0.22508 | < 2.220446e-16 |
| DHA% (Raw) | OFI | 0.328 | < 2.220446e-16 | 0.32770 | < 2.220446e-16 |
| Omega-3 (Raw) | Sex | -0.147 | < 2.220446e-16 | -0.14671 | < 2.220446e-16 |
| Omega-3 (Raw) | Age | 0.214 | < 2.220446e-16 | 0.20859 | < 2.220446e-16 |
| Omega-3 (Raw) | sex-by-age | -0.131 | < 2.220446e-16 | -0.11390 | < 2.220446e-16 |
| Omega-3 (Raw) | BMI | -0.002 | 5E-01 | 0.01280 | 6.89E-10 |
| Omega-3 (Raw) | TSI | -0.095 | < 2.220446e-16 | -0.09189 | < 2.220446e-16 |
| Omega-3 (Raw) | Smoking | -0.053 | < 2.220446e-16 | -0.03767 | < 2.220446e-16 |
| Omega-3 (Raw) | Alcohol use | 0.037 | < 2.220446e-16 | 0.04538 | < 2.220446e-16 |
| Omega-3 (Raw) | Physical activity | 0.002 | 3E-01 | -0.00166 | 4.24E-01 |
| Omega-3 (Raw) | Statin use | 0.040 | < 2.220446e-16 | 0.04493 | < 2.220446e-16 |
| Omega-3 (Raw) | FOS | 0.228 | < 2.220446e-16 | 0.22754 | < 2.220446e-16 |
| Omega-3 (Raw) | OFI | 0.275 | < 2.220446e-16 | 0.27676 | < 2.220446e-16 |
| Omega-3% (Raw) | Sex | -0.137 | < 2.220446e-16 | -0.13814 | < 2.220446e-16 |
| Omega-3% (Raw) | Age | 0.205 | < 2.220446e-16 | 0.20444 | < 2.220446e-16 |
| Omega-3% (Raw) | sex-by-age | -0.114 | < 2.220446e-16 | -0.09075 | < 2.220446e-16 |
| Omega-3% (Raw) | BMI | -0.089 | < 2.220446e-16 | -0.07808 | < 2.220446e-16 |
| Omega-3% (Raw) | TSI | -0.120 | < 2.220446e-16 | -0.11609 | < 2.220446e-16 |

Continued

| Omega-3% (Raw) | Smoking | -0.091 | < 2.220446e-16 | -0.07150 | < 2.220446e-16 |
| --- | --- | --- | --- | --- | --- |
| Omega-3% (Raw) | Alcohol use | 0.039 | < 2.220446e-16 | 0.04570 | < 2.220446e-16 |
| Omega-3% (Raw) | Physical activity | 0.013 | 1E-10 | 0.00812 | 9.30E-05 |
| Omega-3% (Raw) | Statin use | 0.132 | < 2.220446e-16 | 0.13942 | < 2.220446e-16 |
| Omega-3% (Raw) | FOS | 0.258 | < 2.220446e-16 | 0.25858 | < 2.220446e-16 |
| Omega-3% (Raw) | OFI | 0.339 | < 2.220446e-16 | 0.33999 | < 2.220446e-16 |

Pearson and Spearman correlation coefficients and p-values are reported. PGS for DHA, DHA%, Omega-3, and Omega-3% were denoted as PGS_DHA, PGS_DHA%, PGS_Omega-3, and PGS_Omega-3%, respectively. PGS for the four traits were calculated using SBayesRC in a sample of 237,380 participants of European ancestry. Abbreviations: BMI, body mass index; Omega-3, omega-3 fatty acids; Omega-3%, omega-3 fatty acids to total fatty acids percentage; DHA, docosahexaenoic acid; DHA%, docosahexaenoic acid to total fatty acids percentage; EUR, European; FOS, fish oil supplementation; PGS, polygenic score; TSI, Townsend index; OFI, oily fish intake; RINT, rank-based inverse normal transformation; PhysicalAct, physical activity.

### **Supplementary Table 4.** Association of covariates with the observed circulating concentrations of omega-3 fatty acids in EUR participants (discovery analysis)

| **Trait** | **DHA** | **DHA%** | **Omega-3** | **Omega-3%** | **DHA** | **DHA%** | **Omega-3** | **Omega-3%** |
| --- | --- | --- | --- | --- | --- | --- | --- | --- |
| Data type | RINT | RINT | RINT | RINT | Raw | Raw | Raw | Raw |
| Sample size | 230,691 | 231,052 | 231,243 | 230,657 | 230,691 | 231,052 | 231,243 | 230,657 |
| Beta (Sex) | 0.3748 | -0.6452 | 1.2142 | 0.4358 | 0.0318 | -0.3440 | 0.2359 | 0.5875 |
| SE (Sex) | 0.0250 | 0.0250 | 0.0256 | 0.0249 | 0.0018 | 0.0143 | 0.0048 | 0.0321 |
| P (Sex) | 1.29E-50 | 9.41E-147 | 1.00E-300 | 1.26E-68 | 1.26E-73 | 2.72E-128 | 1.00E-300 | 1.03E-74 |
| Beta (Age) | 0.0214 | 0.0010 | 0.0321 | 0.0183 | 0.0016 | 0.0008 | 0.0061 | 0.0240 |
| SE (Age) | 0.0003 | 0.0003 | 0.0003 | 0.0003 | 0.0000 | 0.0002 | 0.0001 | 0.0004 |
| P (Age) | 1.00E-300 | 0.0009 | 1.00E-300 | 1.00E-300 | 1.00E-300 | 1.73E-06 | 1.00E-300 | 1.00E-300 |
| Beta (Sex-by-age) | -0.0146 | 0.0049 | -0.0264 | -0.0123 | -0.0011 | 0.0024 | -0.0051 | -0.0163 |
| SE (Sex-by-age) | 0.0004 | 0.0004 | 0.0004 | 0.0004 | 0.0000 | 0.0002 | 0.0001 | 0.0006 |
| P (Sex-by-age) | 1.77E-243 | 4.16E-29 | 1.00E-300 | 1.08E-175 | 1.63E-286 | 1.51E-22 | 1.00E-300 | 4.99E-185 |
| Beta (BMI) | -1.34E-05 | -2.62E-05 | 3.13E-06 | -8.47E-06 | -9.12E-07 | -1.48E-05 | 5.09E-07 | -1.16E-05 |
| SE (BMI) | 1.93E-07 | 1.92E-07 | 1.97E-07 | 1.91E-07 | 1.35E-08 | 1.10E-07 | 3.73E-08 | 2.47E-07 |
| P (BMI) | 1.00E-300 | 1.00E-300 | 7.40E-57 | 1.00E-300 | 1.00E-300 | 1.00E-300 | 1.60E-42 | 1.00E-300 |
| Beta (TSI) | -4.26E-06 | -4.34E-06 | -4.21E-06 | -5.15E-06 | -2.86E-07 | -2.43E-06 | -7.40E-07 | -6.34E-06 |
| SE (TSI) | 1.24E-07 | 1.24E-07 | 1.27E-07 | 1.23E-07 | 8.69E-09 | 7.07E-08 | 2.40E-08 | 1.59E-07 |
| P (TSI) | 4.48E-257 | 4.03E-269 | 2.99E-240 | 1.00E-300 | 1.98E-236 | 7.92E-258 | 5.98E-208 | 1.00E-300 |
| Beta (Smoking) | -0.0758 | -0.1276 | -0.0389 | -0.1002 | -0.0051 | -0.0713 | -0.0068 | -0.1258 |
| SE (Smoking) | 0.0027 | 0.0026 | 0.0027 | 0.0026 | 0.0002 | 0.0015 | 0.0005 | 0.0034 |
| P (Smoking) | 1.40E-179 | 1.00E-300 | 1.03E-46 | 3.42e-316 | 8.81E-165 | 1.00E-300 | 5.64E-40 | 9.08E-300 |
| Beta (Alcohol) | 0.1710 | 0.1321 | 0.1372 | 0.1274 | 0.0115 | 0.0737 | 0.0244 | 0.1557 |
| SE (Alcohol) | 0.0046 | 0.0045 | 0.0046 | 0.0045 | 0.0003 | 0.0026 | 0.0009 | 0.0058 |
| P (Alcohol) | 5.05E-308 | 7.15E-186 | 4.62E-191 | 1.94E-174 | 3.38E-285 | 8.66E-178 | 6.24E-169 | 1.09E-156 |
| Beta (PhysicalAcivity) | -0.0080 | -0.0005 | -0.0298 | -0.0280 | -0.0006 | -0.0009 | -0.0055 | -0.0354 |
| SE (PhysicalAcivity) | 0.0024 | 0.0024 | 0.0024 | 0.0024 | 0.0002 | 0.0014 | 0.0005 | 0.0031 |

Continued

| P (PhysicalAcivity) | 0.0008 | 0.8185 | 2.97E-34 | 5.07E-32 | 0.0001 | 0.4881 | 5.79E-33 | 5.69E-31 |
| --- | --- | --- | --- | --- | --- | --- | --- | --- |
| Beta (Statin) | -0.0394 | 0.3149 | -0.0237 | 0.3663 | -0.0037 | 0.1759 | -0.0067 | 0.4608 |
| SE (Statin) | 0.0050 | 0.0050 | 0.0051 | 0.0050 | 0.0003 | 0.0028 | 0.0010 | 0.0064 |
| P (Statin) | 2.81E-15 | 1.00E-300 | 3.46E-06 | 1.00E-300 | 1.07E-25 | 1.00E-300 | 5.88E-12 | 1.00E-300 |
| Beta (FOS) | 0.3778 | 0.3599 | 0.3588 | 0.4135 | 0.0267 | 0.2076 | 0.0681 | 0.5364 |
| SE (FOS) | 0.0038 | 0.0038 | 0.0039 | 0.0038 | 0.0003 | 0.0022 | 0.0007 | 0.0049 |
| P (FOS) | 1.00E-300 | 1.00E-300 | 1.00E-300 | 1.00E-300 | 1.00E-300 | 1.00E-300 | 1.00E-300 | 1.00E-300 |
| Beta (OFI) | 0.5127 | 0.5507 | 0.4453 | 0.5644 | 0.0360 | 0.3179 | 0.0833 | 0.7278 |
| SE (OFI) | 0.0036 | 0.0036 | 0.0037 | 0.0035 | 0.0003 | 0.0020 | 0.0007 | 0.0046 |
| P (OFI) | 1.00E-300 | 1.00E-300 | 1.00E-300 | 1.00E-300 | 1.00E-300 | 1.00E-300 | 1.00E-300 | 1.00E-300 |
| Beta (PGS) | 0.2442 | 0.1982 | 0.3033 | 0.2824 | 0.0167 | 0.1098 | 0.0555 | 0.3469 |
| SE (PGS) | 0.0017 | 0.0017 | 0.0018 | 0.0017 | 0.0001 | 0.0010 | 0.0003 | 0.0022 |
| P (PGS) | 1.00E-300 | 1.00E-300 | 1.00E-300 | 1.00E-300 | 1.00E-300 | 1.00E-300 | 1.00E-300 | 1.00E-300 |
| R2 (Sex) | 0.0010 | 0.0029 | 0.0096 | 0.0013 | 0.0014 | 0.0025 | 0.0102 | 0.0014 |
| R2 (Age) | 0.0203 | 4.37E-05 | 0.0425 | 0.0152 | 0.0223 | 9.47E-05 | 0.0430 | 0.0156 |
| R2 (Sex-by-age) | 0.0048 | 0.0005 | 0.0149 | 0.0035 | 0.0057 | 0.0004 | 0.0155 | 0.0036 |
| R2 (BMI) | 0.0206 | 0.0746 | 0.0011 | 0.0084 | 0.0194 | 0.0733 | 0.0008 | 0.0094 |
| R2 (TSI) | 0.0051 | 0.0053 | 0.0047 | 0.0075 | 0.0047 | 0.0051 | 0.0041 | 0.0068 |
| R2 (Smoking) | 0.0035 | 0.0100 | 0.0009 | 0.0062 | 0.0032 | 0.0096 | 0.0008 | 0.0059 |
| R2 (Alcohol) | 0.0061 | 0.0036 | 0.0037 | 0.0034 | 0.0056 | 0.0035 | 0.0033 | 0.0031 |
| R2 (PhysicalAcivity) | 4.43E-05 | -4.10E-06 | 0.0006 | 0.0006 | 6.02E-05 | -2.25E-06 | 0.0006 | 0.0006 |
| R2 (Statin) | 0.0003 | 0.0170 | 0.0001 | 0.0230 | 0.0005 | 0.0163 | 0.0002 | 0.0219 |
| R2 (FOS) | 0.0402 | 0.0369 | 0.0349 | 0.0484 | 0.0411 | 0.0376 | 0.0351 | 0.0489 |
| R2 (OFI) | 0.0811 | 0.0932 | 0.0601 | 0.0983 | 0.0818 | 0.0951 | 0.0589 | 0.0982 |
| R2 (PGS) | 0.0780 | 0.0534 | 0.1109 | 0.1035 | 0.0749 | 0.0504 | 0.1045 | 0.0947 |

A linear regression model excluding the PGS-by-diet interaction term was applied. Effect sizes (Beta and SE) and p-values testing the association between each covariate and circulating concentrations of omega-3 fatty acids are reported. R2 represents the unique contribution of each variable to the variance in each of the fatty acid concentrations when controlling for others. Abbreviations: BMI, body mass index; Omega-3, omega-3 fatty acids; Omega-3%, omega-3 fatty acids to total fatty acids percentage; DHA, docosahexaenoic acid; DHA%, docosahexaenoic acid to total fatty acids percentage; FOS, fish oil supplementation; EUR, European; PGS, polygenic score; TSI, Townsend index; OFI, oily fish intake; SE, standard error; RINT, rank-based inverse normal transformation; PhysicalAct, physical activity; BMI, body mass index.

### **Supplementary Table 5.** Association of PGS-by-FOS with the observed circulating concentrations of omega-3 fatty acids in EUR participants (discovery analysis)

| **Trait** | **Sample Size** | **Data Type** | **Beta (PGS-by-FOS)** | **SE (PGS-by-FOS)** | **P (PGS-by-FOS)** |
| --- | --- | --- | --- | --- | --- |
| DHA | 230,691 | RINT | -0.02543 | 0.00377 | 1.52E-11 |
| DHA% | 231,052 | RINT | -0.01875 | 0.00375 | 5.91E-07 |
| Omega-3 | 231,243 | RINT | -0.03981 | 0.00385 | 4.38E-25 |
| Omega-3% | 230,657 | RINT | -0.04313 | 0.00374 | 1.02E-30 |
| DHA | 230,691 | Raw | -0.00054 | 0.00026 | 0.04092 |
| DHA% | 231,052 | Raw | -0.00522 | 0.00214 | 0.01487 |
| Omega-3 | 231,243 | Raw | -0.00265 | 0.00073 | 0.00028 |
| Omega-3% | 230,657 | Raw | -0.02651 | 0.00483 | 4.10E-08 |

A linear regression model including the PGS-by-FOS interaction term was applied. Effect sizes (Beta and SE) and p-values testing the association between PGS-by-FOS and circulating concentrations of omega-3 fatty acids are reported. Abbreviations: BMI, body mass index; Omega-3, omega-3 fatty acids; Omega-3%, omega-3 fatty acids to total fatty acids percentage; DHA, docosahexaenoic acid; DHA%, docosahexaenoic acid to total fatty acids percentage; EUR, European; FOS, fish oil supplementation; PGS, polygenic score; TSI, Townsend index; OFI, oily fish intake; SE, standard error; RINT, rank-based inverse normal transformation.

### **Supplementary Table 6.** Association of FOS with the observed circulating concentrations of omega-3 fatty acids stratified by PGS groups in EUR participants (discovery analysis)

| **Trait** | **Group** | **Data Type** | **Beta (FOS)** | **SE (FOS)** | **P (FOS)** | **R2 (FOS)** |
| --- | --- | --- | --- | --- | --- | --- |
| DHA | all PGS | RINT | 0.3778 | 0.0038 | 1.00E-300 | 0.0402 |
| DHA | 0-5% PGS | RINT | 0.4064 | 0.0179 | 8.62E-112 | 0.0428 |
| DHA | 95-100% PGS | RINT | 0.3364 | 0.0168 | 2.27E-87 | 0.0334 |
| DHA | 0-20% PGS | RINT | 0.4134 | 0.0088 | 1.00E-300 | 0.0453 |
| DHA | 20-40% PGS | RINT | 0.3862 | 0.0087 | 1.00E-300 | 0.0414 |
| DHA | 40-60% PGS | RINT | 0.3770 | 0.0086 | 1.00E-300 | 0.0401 |
| DHA | 60-80% PGS | RINT | 0.3653 | 0.0085 | 1.00E-300 | 0.0385 |
| DHA | 80-100% PGS | RINT | 0.3472 | 0.0084 | 1.00E-300 | 0.0358 |
| DHA | all PGS | Raw | 0.0267 | 0.0003 | 1.00E-300 | 0.0411 |
| DHA | 0-5% PGS | Raw | 0.0267 | 0.0012 | 5.49E-113 | 0.0433 |
| DHA | 95-100% PGS | Raw | 0.0253 | 0.0012 | 2.90E-90 | 0.0345 |
| DHA | 0-20% PGS | Raw | 0.0279 | 0.0006 | 1.00E-300 | 0.0463 |
| DHA | 20-40% PGS | Raw | 0.0268 | 0.0006 | 1.00E-300 | 0.0418 |
| DHA | 40-60% PGS | Raw | 0.0268 | 0.0006 | 1.00E-300 | 0.0412 |
| DHA | 60-80% PGS | Raw | 0.0265 | 0.0006 | 1.00E-300 | 0.0399 |
| DHA | 80-100% PGS | Raw | 0.0257 | 0.0006 | 1.00E-300 | 0.0370 |
| DHA% | all PGS | RINT | 0.3599 | 0.0038 | 1.00E-300 | 0.0369 |
| DHA% | 0-5% PGS | RINT | 0.3983 | 0.0191 | 5.02E-95 | 0.0363 |
| DHA% | 95-100% PGS | RINT | 0.3325 | 0.0159 | 8.43E-95 | 0.0362 |
| DHA% | 0-20% PGS | RINT | 0.3819 | 0.0092 | 1.00E-300 | 0.0363 |
| DHA% | 20-40% PGS | RINT | 0.3680 | 0.0087 | 1.00E-300 | 0.0369 |
| DHA% | 40-60% PGS | RINT | 0.3513 | 0.0085 | 1.00E-300 | 0.0356 |
| DHA% | 60-80% PGS | RINT | 0.3567 | 0.0083 | 1.00E-300 | 0.0388 |
| DHA% | 80-100% PGS | RINT | 0.3409 | 0.0081 | 1.00E-300 | 0.0373 |
| DHA% | all PGS | Raw | 0.2076 | 0.0022 | 1.00E-300 | 0.0376 |

Continued

| DHA% | 0-5% PGS | Raw | 0.2208 | 0.0105 | 4.48E-97 | 0.0371 |
| --- | --- | --- | --- | --- | --- | --- |
| DHA% | 95-100% PGS | Raw | 0.1981 | 0.0094 | 6.65E-97 | 0.0370 |
| DHA% | 0-20% PGS | Raw | 0.2140 | 0.0051 | 1.00E-300 | 0.0369 |
| DHA% | 20-40% PGS | Raw | 0.2101 | 0.0050 | 1.00E-300 | 0.0374 |
| DHA% | 40-60% PGS | Raw | 0.2039 | 0.0049 | 1.00E-300 | 0.0365 |
| DHA% | 60-80% PGS | Raw | 0.2080 | 0.0048 | 1.00E-300 | 0.0393 |
| DHA% | 80-100% PGS | Raw | 0.2021 | 0.0047 | 1.00E-300 | 0.0382 |
| Omega-3 | all PGS | RINT | 0.3588 | 0.0039 | 1.00E-300 | 0.0349 |
| Omega-3 | 0-5% PGS | RINT | 0.4046 | 0.0187 | 5.15E-102 | 0.0390 |
| Omega-3 | 95-100% PGS | RINT | 0.2850 | 0.0169 | 4.94E-63 | 0.0240 |
| Omega-3 | 0-20% PGS | RINT | 0.4030 | 0.0091 | 1.00E-300 | 0.0404 |
| Omega-3 | 20-40% PGS | RINT | 0.3830 | 0.0088 | 1.00E-300 | 0.0392 |
| Omega-3 | 40-60% PGS | RINT | 0.3537 | 0.0087 | 1.00E-300 | 0.0343 |
| Omega-3 | 60-80% PGS | RINT | 0.3450 | 0.0086 | 1.00E-300 | 0.0334 |
| Omega-3 | 80-100% PGS | RINT | 0.3090 | 0.0085 | 3.37E-283 | 0.0276 |
| Omega-3 | all PGS | Raw | 0.0681 | 0.0007 | 1.00E-300 | 0.0351 |
| Omega-3 | 0-5% PGS | Raw | 0.0674 | 0.0032 | 1.54E-99 | 0.0381 |
| Omega-3 | 95-100% PGS | Raw | 0.0586 | 0.0035 | 7.33E-64 | 0.0243 |
| Omega-3 | 0-20% PGS | Raw | 0.0707 | 0.0016 | 1.00E-300 | 0.0402 |
| Omega-3 | 20-40% PGS | Raw | 0.0712 | 0.0016 | 1.00E-300 | 0.0393 |
| Omega-3 | 40-60% PGS | Raw | 0.0679 | 0.0017 | 1.00E-300 | 0.0350 |
| Omega-3 | 60-80% PGS | Raw | 0.0681 | 0.0017 | 1.00E-300 | 0.0341 |
| Omega-3 | 80-100% PGS | Raw | 0.0629 | 0.0017 | 2.25E-290 | 0.0283 |
| Omega-3% | all PGS | RINT | 0.4135 | 0.0038 | 1.00E-300 | 0.0484 |
| Omega-3% | 0-5% PGS | RINT | 0.4970 | 0.0195 | 1.69E-139 | 0.0534 |
| Omega-3% | 95-100% PGS | RINT | 0.3637 | 0.0156 | 3.43E-117 | 0.0449 |
| Omega-3% | 0-20% PGS | RINT | 0.4648 | 0.0093 | 1.00E-300 | 0.0510 |

Continued

| Omega-3% | 20-40% PGS | RINT | 0.4307 | 0.0087 | 1.00E-300 | 0.0505 |
| --- | --- | --- | --- | --- | --- | --- |
| Omega-3% | 40-60% PGS | RINT | 0.4124 | 0.0084 | 1.00E-300 | 0.0494 |
| Omega-3% | 60-80% PGS | RINT | 0.3801 | 0.0082 | 1.00E-300 | 0.0445 |
| Omega-3% | 80-100% PGS | RINT | 0.3789 | 0.0079 | 1.00E-300 | 0.0473 |
| Omega-3% | all PGS | Raw | 0.5364 | 0.0049 | 1.00E-300 | 0.0489 |
| Omega-3% | 0-5% PGS | Raw | 0.5885 | 0.0232 | 1.27E-137 | 0.0526 |
| Omega-3% | 95-100% PGS | Raw | 0.5046 | 0.0212 | 6.27E-122 | 0.0467 |
| Omega-3% | 0-20% PGS | Raw | 0.5679 | 0.0115 | 1.00E-300 | 0.0506 |
| Omega-3% | 20-40% PGS | Raw | 0.5499 | 0.0111 | 1.00E-300 | 0.0504 |
| Omega-3% | 40-60% PGS | Raw | 0.5380 | 0.0110 | 1.00E-300 | 0.0497 |
| Omega-3% | 60-80% PGS | Raw | 0.5078 | 0.0109 | 1.00E-300 | 0.0452 |
| Omega-3% | 80-100% PGS | Raw | 0.5183 | 0.0107 | 1.00E-300 | 0.0486 |

The PGS were stratified into seven groups based on their distribution: the bottom 5% (0–5%), five quintile-based bins (0–20%, 20–40%, 40–60%, 60–80%, and 80–100%), and the top 5% (95–100%). A linear regression model excluding the PGS-by-FOS interaction term was applied in each PGS group. Effect sizes (Beta and SE) and p-values testing the association between FOS and circulating concentrations of omega-3 fatty acids are reported. Abbreviations: Omega-3, omega-3 fatty acids; Omega-3%, omega-3 fatty acids to total fatty acids percentage; DHA, docosahexaenoic acid; DHA%, docosahexaenoic acid to total fatty acids percentage; FOS, fish oil supplementation; PGS, polygenic score; SE, standard error; RINT, rank-based inverse normal transformation.

### **Supplementary Table 7.** Association of PGS with the observed circulating concentrations of omega-3 fatty acids stratified by FOS status in EUR participants (discovery analysis)

| **Trait** | **Sample Size** | **Data Type** | **Dietary group** | **Beta (PGS)** | **SE (PGS)** | **P (PGS)** | **R2 (PGS)** |
| --- | --- | --- | --- | --- | --- | --- | --- |
| DHA | 72,024 | RINT | FOS | 0.2281 | 0.0032 | 1.00E-300 | 0.0672 |
| DHA | 158,667 | RINT | non-FOS | 0.2513 | 0.0021 | 1.00E-300 | 0.0833 |
| DHA% | 72,400 | RINT | FOS | 0.1855 | 0.0032 | 1.00E-300 | 0.0450 |
| DHA% | 158,652 | RINT | non-FOS | 0.2038 | 0.0021 | 1.00E-300 | 0.0575 |
| Omega-3 | 72,313 | RINT | FOS | 0.2771 | 0.0032 | 1.00E-300 | 0.0950 |
| Omega-3 | 158,930 | RINT | non-FOS | 0.3150 | 0.0022 | 1.00E-300 | 0.1186 |
| Omega-3% | 71,971 | RINT | FOS | 0.2525 | 0.0031 | 1.00E-300 | 0.0844 |
| Omega-3% | 158,686 | RINT | non-FOS | 0.2958 | 0.0021 | 1.00E-300 | 0.1130 |
| DHA | 72,024 | Raw | FOS | 0.0164 | 0.0002 | 1.00E-300 | 0.0659 |
| DHA | 158,667 | Raw | non-FOS | 0.0168 | 0.0001 | 1.00E-300 | 0.0797 |
| DHA% | 72,400 | Raw | FOS | 0.1062 | 0.0019 | 1.00E-300 | 0.0432 |
| DHA% | 158,652 | Raw | non-FOS | 0.1114 | 0.0012 | 1.00E-300 | 0.0542 |
| Omega-3 | 72,313 | Raw | FOS | 0.0540 | 0.0006 | 1.00E-300 | 0.0917 |
| Omega-3 | 158,930 | Raw | non-FOS | 0.0562 | 0.0004 | 1.00E-300 | 0.1113 |
| Omega-3% | 71,971 | Raw | FOS | 0.3283 | 0.0042 | 1.00E-300 | 0.0792 |
| Omega-3% | 158,686 | Raw | non-FOS | 0.3552 | 0.0026 | 1.00E-300 | 0.1029 |

A linear regression model excluding the PGS-by-FOS interaction term was applied stratified by FOS status (yes vs no). Effect sizes (Beta and SE) and p-values testing the association between PGS and circulating concentrations of omega-3 fatty acids are reported. Abbreviations: Omega-3, omega-3 fatty acids; Omega-3%, omega-3 fatty acids to total fatty acids percentage; DHA, docosahexaenoic acid; DHA%, docosahexaenoic acid to total fatty acids percentage; FOS, fish oil supplementation; PGS, polygenic score; SE, standard error; RINT, rank-based inverse normal transformation.

### **Supplementary Table 8.** Association of PGS-by-diet with the observed circulating concentrations of omega-3 fatty acids in EUR participants (replication analysis)

| **Trait** | **Sample Size** | **Population** | **Data Type** | **Dietary exposure** | **Beta (PGS-by-diet)** | **SE (PGS-by-diet)** | **P (PGS-by-diet)** |
| --- | --- | --- | --- | --- | --- | --- | --- |
| DHA | 230,691 | European (Phase 2) | Raw | OFI | -0.0005 | 0.0002 | 0.0390 |
| DHA | 230,691 | European (Phase 2) | RINT | OFI | -0.0298 | 0.0035 | 1.56E-17 |
| DHA% | 231,052 | European (Phase 2) | Raw | OFI | -0.0079 | 0.0020 | 7.93E-05 |
| DHA% | 231,052 | European (Phase 2) | RINT | OFI | -0.0270 | 0.0035 | 1.11E-14 |
| Omega-3 | 231,243 | European (Phase 2) | Raw | OFI | -0.0033 | 0.0007 | 1.04E-06 |
| Omega-3 | 231,243 | European (Phase 2) | RINT | OFI | -0.0481 | 0.0036 | 2.70E-41 |
| Omega-3% | 230,657 | European (Phase 2) | Raw | OFI | -0.0433 | 0.0045 | 4.84E-22 |
| Omega-3% | 230,657 | European (Phase 2) | RINT | OFI | -0.0616 | 0.0035 | 4.12E-70 |
| DHA | 173,827 | European (additional set from Phase 3) | Raw | FOS | -0.0007 | 0.0003 | 0.0385 |
| DHA | 173,827 | European (additional set from Phase 3) | RINT | FOS | -0.0251 | 0.0044 | 1.01E-08 |
| DHA% | 174,187 | European (additional set from Phase 3) | Raw | FOS | -0.0084 | 0.0025 | 0.0007 |
| DHA% | 174,187 | European (additional set from Phase 3) | RINT | FOS | -0.0235 | 0.0043 | 5.67E-08 |
| Omega-3 | 174,285 | European (additional set from Phase 3) | Raw | FOS | -0.0035 | 0.0009 | 4.75E-05 |
| Omega-3 | 174,285 | European (additional set from Phase 3) | RINT | FOS | -0.0426 | 0.0045 | 1.25E-21 |
| Omega-3% | 173,649 | European (additional set from Phase 3) | Raw | FOS | -0.0353 | 0.0054 | 6.80E-11 |
| Omega-3% | 173,649 | European (additional set from Phase 3) | RINT | FOS | -0.0505 | 0.0043 | 1.95E-31 |
| DHA | 173,827 | European (additional set from Phase 3) | Raw | OFI | -0.0010 | 0.0003 | 0.0009 |
| DHA | 173,827 | European (additional set from Phase 3) | RINT | OFI | -0.0328 | 0.0041 | 7.06E-16 |
| DHA% | 174,187 | European (additional set from Phase 3) | Raw | OFI | -0.0101 | 0.0023 | 1.37E-05 |
| DHA% | 174,187 | European (additional set from Phase 3) | RINT | OFI | -0.0296 | 0.0040 | 2.22E-13 |

Continued

| Omega-3 | 174,285 | European (additional set from Phase 3) | Raw | OFI | -0.0045 | 0.0008 | 1.17E-08 |
| --- | --- | --- | --- | --- | --- | --- | --- |
| Omega-3 | 174,285 | European (additional set from Phase 3) | RINT | OFI | -0.0515 | 0.0041 | 1.62E-35 |
| Omega-3% | 173,649 | European (additional set from Phase 3) | Raw | OFI | -0.0478 | 0.0050 | 1.92E-21 |
| Omega-3% | 173,649 | European (additional set from Phase 3) | RINT | OFI | -0.0650 | 0.0040 | 6.63E-59 |

Linear regression model including the PGS-by-FOS interaction term was applied. When FOS is defined as the dietary exposure variable, all reported statistics, including the coefficient (Beta), standard error (SE), p-value, variance explained (R2), and interaction terms, pertain specifically to the effects of FOS. Conversely, when OFI is used as dietary exposure, these statistical parameters apply to OFI. Abbreviations: Omega-3, omega-3 fatty acids; Omega-3%, omega-3 fatty acids to total fatty acids percentage; DHA, docosahexaenoic acid; DHA%, docosahexaenoic acid to total fatty acids percentage; FOS, fish oil supplementation; OFI, oily fish intake; PGS, polygenic score; SE, standard error; RINT, rank-based inverse normal transformation.

### **Supplementary Table 9.** Association of dietary exposures and the observed circulating concentrations of omega-3 fatty acids stratified by PGS groups in EUR participants (replication analysis)

| **Trait** | **Population** | **Group** | **Data Type** | **Dietary exposure** | **Beta (Dietary exposure)** | **SE (Dietary exposure)** | **P (Dietary exposure)** | **R2 (Dietary exposure)** |
| --- | --- | --- | --- | --- | --- | --- | --- | --- |
| DHA | European (Phase 2) | all PGS | RINT | OFI | 0.513 | 0.004 | 1.00E-300 | 0.081 |
| DHA | European (Phase 2) | 0-5% PGS | RINT | OFI | 0.562 | 0.016 | 2.27E-243 | 0.091 |
| DHA | European (Phase 2) | 95-100% PGS | RINT | OFI | 0.483 | 0.016 | 1.39E-200 | 0.076 |
| DHA | European (Phase 2) | 0-20% PGS | RINT | OFI | 0.558 | 0.008 | 1.00E-300 | 0.091 |
| DHA | European (Phase 2) | 20-40% PGS | RINT | OFI | 0.519 | 0.008 | 1.00E-300 | 0.083 |
| DHA | European (Phase 2) | 40-60% PGS | RINT | OFI | 0.508 | 0.008 | 1.00E-300 | 0.080 |
| DHA | European (Phase 2) | 60-80% PGS | RINT | OFI | 0.490 | 0.008 | 1.00E-300 | 0.076 |
| DHA | European (Phase 2) | 80-100% PGS | RINT | OFI | 0.487 | 0.008 | 1.00E-300 | 0.076 |
| DHA | European (Phase 2) | all PGS | Raw | OFI | 0.036 | 0.000 | 1.00E-300 | 0.082 |
| DHA | European (Phase 2) | 0-5% PGS | Raw | OFI | 0.037 | 0.001 | 4.98E-242 | 0.091 |
| DHA | European (Phase 2) | 95-100% PGS | Raw | OFI | 0.036 | 0.001 | 1.99E-205 | 0.078 |
| DHA | European (Phase 2) | 0-20% PGS | Raw | OFI | 0.037 | 0.001 | 1.00E-300 | 0.090 |
| DHA | European (Phase 2) | 20-40% PGS | Raw | OFI | 0.036 | 0.001 | 1.00E-300 | 0.083 |
| DHA | European (Phase 2) | 40-60% PGS | Raw | OFI | 0.036 | 0.001 | 1.00E-300 | 0.080 |
| DHA | European (Phase 2) | 60-80% PGS | Raw | OFI | 0.035 | 0.001 | 1.00E-300 | 0.077 |
| DHA | European (Phase 2) | 80-100% PGS | Raw | OFI | 0.036 | 0.001 | 1.00E-300 | 0.078 |
| DHA% | European (Phase 2) | all PGS | RINT | OFI | 0.551 | 0.004 | 1.00E-300 | 0.093 |
| DHA% | European (Phase 2) | 0-5% PGS | RINT | OFI | 0.620 | 0.017 | 2.20E-263 | 0.098 |
| DHA% | European (Phase 2) | 95-100% PGS | RINT | OFI | 0.531 | 0.015 | 8.84E-260 | 0.098 |
| DHA% | European (Phase 2) | 0-20% PGS | RINT | OFI | 0.590 | 0.008 | 1.00E-300 | 0.095 |
| DHA% | European (Phase 2) | 20-40% PGS | RINT | OFI | 0.561 | 0.008 | 1.00E-300 | 0.092 |
| DHA% | European (Phase 2) | 40-60% PGS | RINT | OFI | 0.543 | 0.008 | 1.00E-300 | 0.092 |
| DHA% | European (Phase 2) | 60-80% PGS | RINT | OFI | 0.529 | 0.008 | 1.00E-300 | 0.092 |
| DHA% | European (Phase 2) | 80-100% PGS | RINT | OFI | 0.531 | 0.008 | 1.00E-300 | 0.096 |

Continued

| DHA% | European (Phase 2) | all PGS | Raw | OFI | 0.318 | 0.002 | 1.00E-300 | 0.095 |
| --- | --- | --- | --- | --- | --- | --- | --- | --- |
| DHA% | European (Phase 2) | 0-5% PGS | Raw | OFI | 0.342 | 0.010 | 6.77E-267 | 0.099 |
| DHA% | European (Phase 2) | 95-100% PGS | Raw | OFI | 0.316 | 0.009 | 2.32E-264 | 0.099 |
| DHA% | European (Phase 2) | 0-20% PGS | Raw | OFI | 0.331 | 0.005 | 1.00E-300 | 0.097 |
| DHA% | European (Phase 2) | 20-40% PGS | Raw | OFI | 0.321 | 0.005 | 1.00E-300 | 0.094 |
| DHA% | European (Phase 2) | 40-60% PGS | Raw | OFI | 0.314 | 0.005 | 1.00E-300 | 0.094 |
| DHA% | European (Phase 2) | 60-80% PGS | Raw | OFI | 0.310 | 0.004 | 1.00E-300 | 0.094 |
| DHA% | European (Phase 2) | 80-100% PGS | Raw | OFI | 0.314 | 0.004 | 1.00E-300 | 0.098 |
| Omega-3 | European (Phase 2) | all PGS | RINT | OFI | 0.445 | 0.004 | 1.00E-300 | 0.060 |
| Omega-3 | European (Phase 2) | 0-5% PGS | RINT | OFI | 0.557 | 0.017 | 2.21E-220 | 0.083 |
| Omega-3 | European (Phase 2) | 95-100% PGS | RINT | OFI | 0.380 | 0.016 | 1.07E-124 | 0.048 |
| Omega-3 | European (Phase 2) | 0-20% PGS | RINT | OFI | 0.507 | 0.008 | 1.00E-300 | 0.072 |
| Omega-3 | European (Phase 2) | 20-40% PGS | RINT | OFI | 0.471 | 0.008 | 1.00E-300 | 0.067 |
| Omega-3 | European (Phase 2) | 40-60% PGS | RINT | OFI | 0.429 | 0.008 | 1.00E-300 | 0.056 |
| Omega-3 | European (Phase 2) | 60-80% PGS | RINT | OFI | 0.427 | 0.008 | 1.00E-300 | 0.057 |
| Omega-3 | European (Phase 2) | 80-100% PGS | RINT | OFI | 0.392 | 0.008 | 1.00E-300 | 0.049 |
| Omega-3 | European (Phase 2) | all PGS | Raw | OFI | 0.083 | 0.001 | 1.00E-300 | 0.059 |
| Omega-3 | European (Phase 2) | 0-5% PGS | Raw | OFI | 0.092 | 0.003 | 1.10E-211 | 0.080 |
| Omega-3 | European (Phase 2) | 95-100% PGS | Raw | OFI | 0.078 | 0.003 | 7.25E-126 | 0.048 |
| Omega-3 | European (Phase 2) | 0-20% PGS | Raw | OFI | 0.088 | 0.001 | 1.00E-300 | 0.070 |
| Omega-3 | European (Phase 2) | 20-40% PGS | Raw | OFI | 0.086 | 0.002 | 1.00E-300 | 0.065 |
| Omega-3 | European (Phase 2) | 40-60% PGS | Raw | OFI | 0.081 | 0.002 | 1.00E-300 | 0.056 |
| Omega-3 | European (Phase 2) | 60-80% PGS | Raw | OFI | 0.083 | 0.002 | 1.00E-300 | 0.057 |
| Omega-3 | European (Phase 2) | 80-100% PGS | Raw | OFI | 0.079 | 0.002 | 1.00E-300 | 0.049 |
| Omega-3% | European (Phase 2) | all PGS | RINT | OFI | 0.564 | 0.004 | 1.00E-300 | 0.098 |
| Omega-3% | European (Phase 2) | 0-5% PGS | RINT | OFI | 0.719 | 0.018 | 1.00E-300 | 0.123 |
| Omega-3% | European (Phase 2) | 95-100% PGS | RINT | OFI | 0.514 | 0.015 | 7.51E-256 | 0.097 |
| Omega-3% | European (Phase 2) | 0-20% PGS | RINT | OFI | 0.655 | 0.009 | 1.00E-300 | 0.110 |

Continued

| Omega-3% | European (Phase 2) | 20-40% PGS | RINT | OFI | 0.589 | 0.008 | 1.00E-300 | 0.103 |
| --- | --- | --- | --- | --- | --- | --- | --- | --- |
| Omega-3% | European (Phase 2) | 40-60% PGS | RINT | OFI | 0.544 | 0.008 | 1.00E-300 | 0.094 |
| Omega-3% | European (Phase 2) | 60-80% PGS | RINT | OFI | 0.522 | 0.008 | 1.00E-300 | 0.092 |
| Omega-3% | European (Phase 2) | 80-100% PGS | RINT | OFI | 0.513 | 0.007 | 1.00E-300 | 0.094 |
| Omega-3% | European (Phase 2) | all PGS | Raw | OFI | 0.728 | 0.005 | 1.00E-300 | 0.098 |
| Omega-3% | European (Phase 2) | 0-5% PGS | Raw | OFI | 0.848 | 0.021 | 1.00E-300 | 0.120 |
| Omega-3% | European (Phase 2) | 95-100% PGS | Raw | OFI | 0.701 | 0.020 | 8.50E-258 | 0.097 |
| Omega-3% | European (Phase 2) | 0-20% PGS | Raw | OFI | 0.796 | 0.011 | 1.00E-300 | 0.109 |
| Omega-3% | European (Phase 2) | 20-40% PGS | Raw | OFI | 0.749 | 0.010 | 1.00E-300 | 0.102 |
| Omega-3% | European (Phase 2) | 40-60% PGS | Raw | OFI | 0.708 | 0.010 | 1.00E-300 | 0.094 |
| Omega-3% | European (Phase 2) | 60-80% PGS | Raw | OFI | 0.694 | 0.010 | 1.00E-300 | 0.093 |
| Omega-3% | European (Phase 2) | 80-100% PGS | Raw | OFI | 0.694 | 0.010 | 1.00E-300 | 0.095 |
| DHA | European (additional set from Phase 3) | all PGS | Raw | FOS | 0.027 | 0.000 | 1.00E-300 | 0.040 |
| DHA | European (additional set from Phase 3) | 0-5% PGS | Raw | FOS | 0.029 | 0.001 | 1.21E-91 | 0.046 |
| DHA | European (additional set from Phase 3) | 95-100% PGS | Raw | FOS | 0.025 | 0.001 | 9.20E-65 | 0.033 |
| DHA | European (additional set from Phase 3) | 0-20% PGS | Raw | FOS | 0.029 | 0.001 | 1.00E-300 | 0.045 |
| DHA | European (additional set from Phase 3) | 20-40% PGS | Raw | FOS | 0.027 | 0.001 | 1.44e-310 | 0.040 |
| DHA | European (additional set from Phase 3) | 40-60% PGS | Raw | FOS | 0.028 | 0.001 | 1.37e-316 | 0.041 |
| DHA | European (additional set from Phase 3) | 60-80% PGS | Raw | FOS | 0.027 | 0.001 | 1.01E-298 | 0.039 |
| DHA | European (additional set from Phase 3) | 80-100% PGS | Raw | FOS | 0.026 | 0.001 | 6.93E-282 | 0.036 |
| DHA% | European (additional set from Phase 3) | all PGS | Raw | FOS | 0.207 | 0.003 | 1.00E-300 | 0.037 |
| DHA% | European (additional set from Phase 3) | 0-5% PGS | Raw | FOS | 0.227 | 0.012 | 2.05E-78 | 0.040 |
| DHA% | European (additional set from Phase 3) | 95-100% PGS | Raw | FOS | 0.190 | 0.011 | 1.03E-66 | 0.034 |
| DHA% | European (additional set from Phase 3) | 0-20% PGS | Raw | FOS | 0.224 | 0.006 | 1.15E-306 | 0.039 |
| DHA% | European (additional set from Phase 3) | 20-40% PGS | Raw | FOS | 0.209 | 0.006 | 2.54E-284 | 0.037 |
| DHA% | European (additional set from Phase 3) | 40-60% PGS | Raw | FOS | 0.202 | 0.006 | 2.21E-274 | 0.035 |
| DHA% | European (additional set from Phase 3) | 60-80% PGS | Raw | FOS | 0.199 | 0.006 | 1.17E-272 | 0.035 |
| DHA% | European (additional set from Phase 3) | 80-100% PGS | Raw | FOS | 0.199 | 0.005 | 3.74E-282 | 0.036 |

Continued

| Omega-3 | European (additional set from Phase 3) | all PGS | Raw | FOS | 0.069 | 0.001 | 1.00E-300 | 0.034 |
| --- | --- | --- | --- | --- | --- | --- | --- | --- |
| Omega-3 | European (additional set from Phase 3) | 0-5% PGS | Raw | FOS | 0.082 | 0.004 | 1.77E-107 | 0.054 |
| Omega-3 | European (additional set from Phase 3) | 95-100% PGS | Raw | FOS | 0.057 | 0.004 | 4.24E-44 | 0.022 |
| Omega-3 | European (additional set from Phase 3) | 0-20% PGS | Raw | FOS | 0.071 | 0.002 | 1.84e-310 | 0.040 |
| Omega-3 | European (additional set from Phase 3) | 20-40% PGS | Raw | FOS | 0.071 | 0.002 | 2.25E-287 | 0.037 |
| Omega-3 | European (additional set from Phase 3) | 40-60% PGS | Raw | FOS | 0.071 | 0.002 | 5.43E-285 | 0.037 |
| Omega-3 | European (additional set from Phase 3) | 60-80% PGS | Raw | FOS | 0.064 | 0.002 | 3.54E-221 | 0.028 |
| Omega-3 | European (additional set from Phase 3) | 80-100% PGS | Raw | FOS | 0.066 | 0.002 | 8.28E-227 | 0.029 |
| Omega-3% | European (additional set from Phase 3) | all PGS | Raw | FOS | 0.515 | 0.006 | 1.00E-300 | 0.048 |
| Omega-3% | European (additional set from Phase 3) | 0-5% PGS | Raw | FOS | 0.584 | 0.026 | 2.35E-111 | 0.056 |
| Omega-3% | European (additional set from Phase 3) | 95-100% PGS | Raw | FOS | 0.472 | 0.024 | 2.58E-82 | 0.042 |
| Omega-3% | European (additional set from Phase 3) | 0-20% PGS | Raw | FOS | 0.560 | 0.013 | 1.00E-300 | 0.052 |
| Omega-3% | European (additional set from Phase 3) | 20-40% PGS | Raw | FOS | 0.531 | 0.012 | 1.00E-300 | 0.049 |
| Omega-3% | European (additional set from Phase 3) | 40-60% PGS | Raw | FOS | 0.523 | 0.012 | 1.00E-300 | 0.049 |
| Omega-3% | European (additional set from Phase 3) | 60-80% PGS | Raw | FOS | 0.490 | 0.012 | 1.00E-300 | 0.045 |
| Omega-3% | European (additional set from Phase 3) | 80-100% PGS | Raw | FOS | 0.471 | 0.012 | 1.00E-300 | 0.042 |
| DHA | European (additional set from Phase 3) | all PGS | RINT | FOS | 0.378 | 0.004 | 1.00E-300 | 0.039 |
| DHA | European (additional set from Phase 3) | 0-5% PGS | RINT | FOS | 0.426 | 0.021 | 6.81E-91 | 0.046 |
| DHA | European (additional set from Phase 3) | 95-100% PGS | RINT | FOS | 0.327 | 0.019 | 9.12E-63 | 0.032 |
| DHA | European (additional set from Phase 3) | 0-20% PGS | RINT | FOS | 0.414 | 0.010 | 1.00E-300 | 0.045 |
| DHA | European (additional set from Phase 3) | 20-40% PGS | RINT | FOS | 0.380 | 0.010 | 2.26E-304 | 0.039 |
| DHA | European (additional set from Phase 3) | 40-60% PGS | RINT | FOS | 0.378 | 0.010 | 0.00E+00 | 0.040 |
| DHA | European (additional set from Phase 3) | 60-80% PGS | RINT | FOS | 0.367 | 0.010 | 2.10E-292 | 0.038 |
| DHA | European (additional set from Phase 3) | 80-100% PGS | RINT | FOS | 0.349 | 0.010 | 4.87E-277 | 0.036 |
| DHA% | European (additional set from Phase 3) | all PGS | RINT | FOS | 0.357 | 0.004 | 1.00E-300 | 0.036 |
| DHA% | European (additional set from Phase 3) | 0-5% PGS | RINT | FOS | 0.406 | 0.022 | 3.28E-77 | 0.039 |
| DHA% | European (additional set from Phase 3) | 95-100% PGS | RINT | FOS | 0.315 | 0.018 | 1.75E-64 | 0.032 |
| DHA% | European (additional set from Phase 3) | 0-20% PGS | RINT | FOS | 0.398 | 0.011 | 4.30E-302 | 0.039 |

Continued

| DHA% | European (additional set from Phase 3) | 20-40% PGS | RINT | FOS | 0.364 | 0.010 | 2.32E-279 | 0.036 |
| --- | --- | --- | --- | --- | --- | --- | --- | --- |
| DHA% | European (additional set from Phase 3) | 40-60% PGS | RINT | FOS | 0.349 | 0.010 | 1.95E-271 | 0.035 |
| DHA% | European (additional set from Phase 3) | 60-80% PGS | RINT | FOS | 0.339 | 0.010 | 8.86E-268 | 0.034 |
| DHA% | European (additional set from Phase 3) | 80-100% PGS | RINT | FOS | 0.334 | 0.009 | 1.41E-275 | 0.035 |
| Omega-3 | European (additional set from Phase 3) | all PGS | RINT | FOS | 0.356 | 0.005 | 1.00E-300 | 0.034 |
| Omega-3 | European (additional set from Phase 3) | 0-5% PGS | RINT | FOS | 0.478 | 0.021 | 7.27E-107 | 0.054 |
| Omega-3 | European (additional set from Phase 3) | 95-100% PGS | RINT | FOS | 0.272 | 0.020 | 8.16E-43 | 0.021 |
| Omega-3 | European (additional set from Phase 3) | 0-20% PGS | RINT | FOS | 0.401 | 0.010 | 6.34e-313 | 0.040 |
| Omega-3 | European (additional set from Phase 3) | 20-40% PGS | RINT | FOS | 0.376 | 0.010 | 3.53E-286 | 0.037 |
| Omega-3 | European (additional set from Phase 3) | 40-60% PGS | RINT | FOS | 0.363 | 0.010 | 2.03E-278 | 0.036 |
| Omega-3 | European (additional set from Phase 3) | 60-80% PGS | RINT | FOS | 0.320 | 0.010 | 3.81E-219 | 0.028 |
| Omega-3 | European (additional set from Phase 3) | 80-100% PGS | RINT | FOS | 0.319 | 0.010 | 4.03E-222 | 0.029 |
| Omega-3% | European (additional set from Phase 3) | all PGS | RINT | FOS | 0.411 | 0.004 | 1.00E-300 | 0.047 |
| Omega-3% | European (additional set from Phase 3) | 0-5% PGS | RINT | FOS | 0.509 | 0.022 | 2.82E-113 | 0.057 |
| Omega-3% | European (additional set from Phase 3) | 95-100% PGS | RINT | FOS | 0.352 | 0.018 | 1.89E-79 | 0.040 |
| Omega-3% | European (additional set from Phase 3) | 0-20% PGS | RINT | FOS | 0.476 | 0.011 | 1.00E-300 | 0.053 |
| Omega-3% | European (additional set from Phase 3) | 20-40% PGS | RINT | FOS | 0.430 | 0.010 | 1.00E-300 | 0.050 |
| Omega-3% | European (additional set from Phase 3) | 40-60% PGS | RINT | FOS | 0.414 | 0.010 | 1.00E-300 | 0.049 |
| Omega-3% | European (additional set from Phase 3) | 60-80% PGS | RINT | FOS | 0.378 | 0.009 | 1.00E-300 | 0.044 |
| Omega-3% | European (additional set from Phase 3) | 80-100% PGS | RINT | FOS | 0.358 | 0.009 | 2.57e-322 | 0.042 |
| DHA | European (additional set from Phase 3) | all PGS | Raw | OFI | 0.037 | 0.000 | 1.00E-300 | 0.081 |
| DHA | European (additional set from Phase 3) | 0-5% PGS | Raw | OFI | 0.037 | 0.001 | 4.65E-169 | 0.084 |
| DHA | European (additional set from Phase 3) | 95-100% PGS | Raw | OFI | 0.032 | 0.001 | 4.49E-120 | 0.061 |
| DHA | European (additional set from Phase 3) | 0-20% PGS | Raw | OFI | 0.038 | 0.001 | 1.00E-300 | 0.087 |
| DHA | European (additional set from Phase 3) | 20-40% PGS | Raw | OFI | 0.038 | 0.001 | 1.00E-300 | 0.086 |
| DHA | European (additional set from Phase 3) | 40-60% PGS | Raw | OFI | 0.038 | 0.001 | 1.00E-300 | 0.083 |
| DHA | European (additional set from Phase 3) | 60-80% PGS | Raw | OFI | 0.037 | 0.001 | 1.00E-300 | 0.081 |
| DHA | European (additional set from Phase 3) | 80-100% PGS | Raw | OFI | 0.035 | 0.001 | 1.00E-300 | 0.071 |

Continued

| DHA% | European (additional set from Phase 3) | all PGS | Raw | OFI | 0.319 | 0.002 | 1.00E-300 | 0.094 |
| --- | --- | --- | --- | --- | --- | --- | --- | --- |
| DHA% | European (additional set from Phase 3) | 0-5% PGS | Raw | OFI | 0.336 | 0.011 | 3.32E-192 | 0.095 |
| DHA% | European (additional set from Phase 3) | 95-100% PGS | Raw | OFI | 0.308 | 0.010 | 2.47E-191 | 0.095 |
| DHA% | European (additional set from Phase 3) | 0-20% PGS | Raw | OFI | 0.331 | 0.005 | 1.00E-300 | 0.094 |
| DHA% | European (additional set from Phase 3) | 20-40% PGS | Raw | OFI | 0.320 | 0.005 | 1.00E-300 | 0.094 |
| DHA% | European (additional set from Phase 3) | 40-60% PGS | Raw | OFI | 0.325 | 0.005 | 1.00E-300 | 0.099 |
| DHA% | European (additional set from Phase 3) | 60-80% PGS | Raw | OFI | 0.319 | 0.005 | 1.00E-300 | 0.098 |
| DHA% | European (additional set from Phase 3) | 80-100% PGS | Raw | OFI | 0.298 | 0.005 | 1.00E-300 | 0.088 |
| Omega-3 | European (additional set from Phase 3) | all PGS | Raw | OFI | 0.085 | 0.001 | 1.00E-300 | 0.059 |
| Omega-3 | European (additional set from Phase 3) | 0-5% PGS | Raw | OFI | 0.086 | 0.003 | 9.72E-137 | 0.068 |
| Omega-3 | European (additional set from Phase 3) | 95-100% PGS | Raw | OFI | 0.069 | 0.004 | 2.18E-73 | 0.037 |
| Omega-3 | European (additional set from Phase 3) | 0-20% PGS | Raw | OFI | 0.090 | 0.002 | 1.00E-300 | 0.072 |
| Omega-3 | European (additional set from Phase 3) | 20-40% PGS | Raw | OFI | 0.089 | 0.002 | 1.00E-300 | 0.066 |
| Omega-3 | European (additional set from Phase 3) | 40-60% PGS | Raw | OFI | 0.083 | 0.002 | 1.00E-300 | 0.057 |
| Omega-3 | European (additional set from Phase 3) | 60-80% PGS | Raw | OFI | 0.084 | 0.002 | 1.00E-300 | 0.056 |
| Omega-3 | European (additional set from Phase 3) | 80-100% PGS | Raw | OFI | 0.078 | 0.002 | 1.00E-300 | 0.047 |
| Omega-3% | European (additional set from Phase 3) | all PGS | Raw | OFI | 0.717 | 0.005 | 1.00E-300 | 0.101 |
| Omega-3% | European (additional set from Phase 3) | 0-5% PGS | Raw | OFI | 0.829 | 0.024 | 3.60E-248 | 0.122 |
| Omega-3% | European (additional set from Phase 3) | 95-100% PGS | Raw | OFI | 0.620 | 0.023 | 1.12E-158 | 0.079 |
| Omega-3% | European (additional set from Phase 3) | 0-20% PGS | Raw | OFI | 0.773 | 0.012 | 1.00E-300 | 0.109 |
| Omega-3% | European (additional set from Phase 3) | 20-40% PGS | Raw | OFI | 0.746 | 0.012 | 1.00E-300 | 0.107 |
| Omega-3% | European (additional set from Phase 3) | 40-60% PGS | Raw | OFI | 0.724 | 0.011 | 1.00E-300 | 0.104 |
| Omega-3% | European (additional set from Phase 3) | 60-80% PGS | Raw | OFI | 0.689 | 0.011 | 1.00E-300 | 0.098 |
| Omega-3% | European (additional set from Phase 3) | 80-100% PGS | Raw | OFI | 0.655 | 0.011 | 1.00E-300 | 0.090 |
| DHA | European (additional set from Phase 3) | all PGS | RINT | OFI | 0.512 | 0.004 | 1.00E-300 | 0.081 |
| DHA | European (additional set from Phase 3) | 0-5% PGS | RINT | OFI | 0.545 | 0.019 | 5.89E-169 | 0.084 |
| DHA | European (additional set from Phase 3) | 95-100% PGS | RINT | OFI | 0.423 | 0.018 | 5.62E-118 | 0.060 |
| DHA | European (additional set from Phase 3) | 0-20% PGS | RINT | OFI | 0.543 | 0.009 | 1.00E-300 | 0.086 |

Continued

| DHA | European (additional set from Phase 3) | 20-40% PGS | RINT | OFI | 0.532 | 0.009 | 1.00E-300 | 0.086 |
| --- | --- | --- | --- | --- | --- | --- | --- | --- |
| DHA | European (additional set from Phase 3) | 40-60% PGS | RINT | OFI | 0.518 | 0.009 | 1.00E-300 | 0.083 |
| DHA | European (additional set from Phase 3) | 60-80% PGS | RINT | OFI | 0.504 | 0.009 | 1.00E-300 | 0.080 |
| DHA | European (additional set from Phase 3) | 80-100% PGS | RINT | OFI | 0.464 | 0.009 | 1.00E-300 | 0.070 |
| DHA% | European (additional set from Phase 3) | all PGS | RINT | OFI | 0.548 | 0.004 | 1.00E-300 | 0.092 |
| DHA% | European (additional set from Phase 3) | 0-5% PGS | RINT | OFI | 0.600 | 0.020 | 3.53E-188 | 0.093 |
| DHA% | European (additional set from Phase 3) | 95-100% PGS | RINT | OFI | 0.513 | 0.017 | 1.99E-186 | 0.093 |
| DHA% | European (additional set from Phase 3) | 0-20% PGS | RINT | OFI | 0.586 | 0.010 | 1.00E-300 | 0.092 |
| DHA% | European (additional set from Phase 3) | 20-40% PGS | RINT | OFI | 0.556 | 0.009 | 1.00E-300 | 0.092 |
| DHA% | European (additional set from Phase 3) | 40-60% PGS | RINT | OFI | 0.558 | 0.009 | 1.00E-300 | 0.097 |
| DHA% | European (additional set from Phase 3) | 60-80% PGS | RINT | OFI | 0.543 | 0.009 | 1.00E-300 | 0.096 |
| DHA% | European (additional set from Phase 3) | 80-100% PGS | RINT | OFI | 0.499 | 0.009 | 1.00E-300 | 0.086 |
| Omega-3 | European (additional set from Phase 3) | all PGS | RINT | OFI | 0.448 | 0.004 | 1.00E-300 | 0.060 |
| Omega-3 | European (additional set from Phase 3) | 0-5% PGS | RINT | OFI | 0.510 | 0.020 | 1.58E-139 | 0.070 |
| Omega-3 | European (additional set from Phase 3) | 95-100% PGS | RINT | OFI | 0.338 | 0.018 | 1.04E-74 | 0.038 |
| Omega-3 | European (additional set from Phase 3) | 0-20% PGS | RINT | OFI | 0.511 | 0.010 | 1.00E-300 | 0.074 |
| Omega-3 | European (additional set from Phase 3) | 20-40% PGS | RINT | OFI | 0.479 | 0.010 | 1.00E-300 | 0.067 |
| Omega-3 | European (additional set from Phase 3) | 40-60% PGS | RINT | OFI | 0.436 | 0.009 | 1.00E-300 | 0.059 |
| Omega-3 | European (additional set from Phase 3) | 60-80% PGS | RINT | OFI | 0.428 | 0.009 | 1.00E-300 | 0.057 |
| Omega-3 | European (additional set from Phase 3) | 80-100% PGS | RINT | OFI | 0.384 | 0.009 | 1.00E-300 | 0.047 |
| Omega-3% | European (additional set from Phase 3) | all PGS | RINT | OFI | 0.574 | 0.004 | 1.00E-300 | 0.101 |
| Omega-3% | European (additional set from Phase 3) | 0-5% PGS | RINT | OFI | 0.725 | 0.021 | 3.01E-253 | 0.124 |
| Omega-3% | European (additional set from Phase 3) | 95-100% PGS | RINT | OFI | 0.470 | 0.017 | 6.36E-158 | 0.079 |
| Omega-3% | European (additional set from Phase 3) | 0-20% PGS | RINT | OFI | 0.655 | 0.010 | 1.00E-300 | 0.110 |
| Omega-3% | European (additional set from Phase 3) | 20-40% PGS | RINT | OFI | 0.607 | 0.009 | 1.00E-300 | 0.108 |
| Omega-3% | European (additional set from Phase 3) | 40-60% PGS | RINT | OFI | 0.575 | 0.009 | 1.00E-300 | 0.103 |
| Omega-3% | European (additional set from Phase 3) | 60-80% PGS | RINT | OFI | 0.534 | 0.009 | 1.00E-300 | 0.097 |
| Omega-3% | European (additional set from Phase 3) | 80-100% PGS | RINT | OFI | 0.500 | 0.009 | 1.00E-300 | 0.089 |

The PGS were stratified into seven groups based on their distribution: the bottom 5% (0–5%), five quintile-based bins (0–20%, 20–40%, 40–60%, 60–80%, and 80–100%), and the top 5% (95–100%). FOS or OFI were defined as dietary exposure, respectively. Linear regression model excluding the PGS-by-diet interaction term was applied in each PGS group. Effect sizes (Beta and SE) and p-values testing the association between dietary exposure and circulating concentrations of omega-3 fatty acids are reported. R2 represents the unique contribution of each dietary exposure to the variance in each of the fatty acid concentrations when controlling for others. Abbreviations: Omega-3, omega-3 fatty acids; Omega-3%, omega-3 fatty acids to total fatty acids percentage; DHA, docosahexaenoic acid; DHA%, docosahexaenoic acid to total fatty acids percentage; FOS, fish oil supplementation; OFI, oily fish intake; PGS, polygenic score; SE, standard error; RINT, rank-based inverse normal transformation.

### **Supplementary Table 10.** Association of PGS with the observed circulating concentrations of omega-3 fatty acids stratified by dietary status in EUR participants (replication analysis)

| **Trait** | **Sample Size** | **Population** | **Data Type** | **Dietary group** | **Beta (PGS)** | **SE (PGS)** | **P (PGS)** | **R2 (PGS)** |
| --- | --- | --- | --- | --- | --- | --- | --- | --- |
| DHA | 127,073 | European (Phase 2) | RINT | high-OFI | 0.232 | 0.002 | 1.00E-300 | 0.066 |
| DHA | 103,618 | European (Phase 2) | RINT | low-OFI | 0.259 | 0.002 | 1.00E-300 | 0.095 |
| DHA% | 127,378 | European (Phase 2) | RINT | high-OFI | 0.186 | 0.002 | 1.00E-300 | 0.043 |
| DHA% | 103,674 | European (Phase 2) | RINT | low-OFI | 0.214 | 0.002 | 1.00E-300 | 0.070 |
| Omega-3 | 127,714 | European (Phase 2) | RINT | high-OFI | 0.283 | 0.002 | 1.00E-300 | 0.095 |
| Omega-3 | 103,529 | European (Phase 2) | RINT | low-OFI | 0.329 | 0.003 | 1.00E-300 | 0.133 |
| Omega-3% | 126,840 | European (Phase 2) | RINT | high-OFI | 0.255 | 0.002 | 1.00E-300 | 0.080 |
| Omega-3% | 103,817 | European (Phase 2) | RINT | low-OFI | 0.317 | 0.002 | 1.00E-300 | 0.140 |
| DHA | 127,073 | European (Phase 2) | Raw | high-OFI | 0.017 | 0.000 | 1.00E-300 | 0.065 |
| DHA | 103,618 | European (Phase 2) | Raw | low-OFI | 0.017 | 0.000 | 1.00E-300 | 0.092 |
| DHA% | 127,378 | European (Phase 2) | Raw | high-OFI | 0.106 | 0.001 | 1.00E-300 | 0.041 |
| DHA% | 103,674 | European (Phase 2) | Raw | low-OFI | 0.115 | 0.001 | 1.00E-300 | 0.067 |
| Omega-3 | 127,714 | European (Phase 2) | Raw | high-OFI | 0.054 | 0.000 | 1.00E-300 | 0.091 |
| Omega-3 | 103,529 | European (Phase 2) | Raw | low-OFI | 0.057 | 0.000 | 1.00E-300 | 0.126 |
| Omega-3% | 126,840 | European (Phase 2) | Raw | high-OFI | 0.327 | 0.003 | 1.00E-300 | 0.074 |
| Omega-3% | 103,817 | European (Phase 2) | Raw | low-OFI | 0.372 | 0.003 | 1.00E-300 | 0.131 |
| DHA | 53,855 | European (additional set from Phase 3) | RINT | FOS | 0.227 | 0.004 | 1.00E-300 | 0.067 |
| DHA | 119,972 | European (additional set from Phase 3) | RINT | non-FOS | 0.251 | 0.002 | 1.00E-300 | 0.081 |
| DHA% | 54,208 | European (additional set from Phase 3) | RINT | FOS | 0.182 | 0.004 | 1.00E-300 | 0.044 |
| DHA% | 119,979 | European (additional set from Phase 3) | RINT | non-FOS | 0.206 | 0.002 | 1.00E-300 | 0.058 |
| Omega-3 | 54,072 | European (additional set from Phase 3) | RINT | FOS | 0.270 | 0.004 | 1.00E-300 | 0.090 |
| Omega-3 | 120,213 | European (additional set from Phase 3) | RINT | non-FOS | 0.312 | 0.002 | 1.00E-300 | 0.115 |
| Omega-3% | 53,729 | European (additional set from Phase 3) | RINT | FOS | 0.249 | 0.004 | 1.00E-300 | 0.083 |
| Omega-3% | 119,920 | European (additional set from Phase 3) | RINT | non-FOS | 0.301 | 0.002 | 1.00E-300 | 0.115 |
| DHA | 53,855 | European (additional set from Phase 3) | Raw | FOS | 0.017 | 0.000 | 1.00E-300 | 0.065 |
| DHA | 119,972 | European (additional set from Phase 3) | Raw | non-FOS | 0.017 | 0.000 | 1.00E-300 | 0.078 |

Continued

| DHA% | 54,208 | European (additional set from Phase 3) | Raw | FOS | 0.105 | 0.002 | 1.00E-300 | 0.042 |
| --- | --- | --- | --- | --- | --- | --- | --- | --- |
| DHA% | 119,979 | European (additional set from Phase 3) | Raw | non-FOS | 0.114 | 0.001 | 1.00E-300 | 0.055 |
| Omega-3 | 54,072 | European (additional set from Phase 3) | Raw | FOS | 0.053 | 0.001 | 1.00E-300 | 0.087 |
| Omega-3 | 120,213 | European (additional set from Phase 3) | Raw | non-FOS | 0.057 | 0.000 | 1.00E-300 | 0.109 |
| Omega-3% | 53,729 | European (additional set from Phase 3) | Raw | FOS | 0.314 | 0.005 | 1.00E-300 | 0.078 |
| Omega-3% | 119,920 | European (additional set from Phase 3) | Raw | non-FOS | 0.350 | 0.003 | 1.00E-300 | 0.105 |
| DHA | 94754 | European (additional set from Phase 3) | RINT | high-OFI | 0.230 | 0.003 | 1.00E-300 | 0.065 |
| DHA | 79073 | European (additional set from Phase 3) | RINT | low-OFI | 0.261 | 0.003 | 1.00E-300 | 0.094 |
| DHA% | 95035 | European (additional set from Phase 3) | RINT | high-OFI | 0.185 | 0.003 | 1.00E-300 | 0.042 |
| DHA% | 79152 | European (additional set from Phase 3) | RINT | low-OFI | 0.215 | 0.003 | 1.00E-300 | 0.071 |
| Omega-3 | 95283 | European (additional set from Phase 3) | RINT | high-OFI | 0.277 | 0.003 | 1.00E-300 | 0.091 |
| Omega-3 | 79002 | European (additional set from Phase 3) | RINT | low-OFI | 0.327 | 0.003 | 1.00E-300 | 0.129 |
| Omega-3% | 94426 | European (additional set from Phase 3) | RINT | high-OFI | 0.255 | 0.003 | 1.00E-300 | 0.079 |
| Omega-3% | 79223 | European (additional set from Phase 3) | RINT | low-OFI | 0.321 | 0.003 | 1.00E-300 | 0.143 |
| DHA | 94754 | European (additional set from Phase 3) | Raw | high-OFI | 0.017 | 0.000 | 1.00E-300 | 0.063 |
| DHA | 79073 | European (additional set from Phase 3) | Raw | low-OFI | 0.018 | 0.000 | 1.00E-300 | 0.091 |
| DHA% | 95035 | European (additional set from Phase 3) | Raw | high-OFI | 0.106 | 0.002 | 1.00E-300 | 0.040 |
| DHA% | 79152 | European (additional set from Phase 3) | Raw | low-OFI | 0.117 | 0.002 | 1.00E-300 | 0.068 |
| Omega-3 | 95283 | European (additional set from Phase 3) | Raw | high-OFI | 0.054 | 0.001 | 1.00E-300 | 0.087 |
| Omega-3 | 79002 | European (additional set from Phase 3) | Raw | low-OFI | 0.058 | 0.001 | 1.00E-300 | 0.123 |
| Omega-3% | 94426 | European (additional set from Phase 3) | Raw | high-OFI | 0.316 | 0.004 | 1.00E-300 | 0.074 |
| Omega-3% | 79223 | European (additional set from Phase 3) | Raw | low-OFI | 0.366 | 0.003 | 1.00E-300 | 0.133 |

FOS or OFI were defined as dietary exposure, respectively. Linear regression model excluding the PGS-by-diet interaction term was applied stratified by dietary status. Effect sizes (Beta and SE) and p-values testing the association between PGS and circulating concentrations of omega-3 fatty acids are reported. Abbreviations: Omega-3, omega-3 fatty acids; Omega-3%, omega-3 fatty acids to total fatty acids percentage; DHA, docosahexaenoic acid; DHA%, docosahexaenoic acid to total fatty acids percentage; FOS, fish oil supplementation; OFI, oily fish intake; PGS, polygenic score; SE, standard error; RINT, rank-based inverse normal transformation.

### **Supplementary Table 11.** Association of dietary exposures, PGS and PGS-by-diet with the observed circulating concentrations of omega-3 fatty acids in non-European ancestry

| Trait | # Sample | Population | Data Type | Dietary exposure | Beta (Dietary exposure) | SE (Dietary exposure) | P  (Dietary exposure) | R2 (Dietary exposure) | Beta (PGS) | SE (PGS) | P (PGS) | R2 (PGS) | Beta (PGS-by-diet) | SE (PGS-by-diet) | P (PGS-by-diet) | R2 (PGS-by-diet) |
| --- | --- | --- | --- | --- | --- | --- | --- | --- | --- | --- | --- | --- | --- | --- | --- | --- |
| DHA | 6,101 | AFR | RINT | FOS | 0.175 | 0.026 | 1.40E-11 | 0.007 | 0.058 | 0.015 | 9.34E-05 | 0.002 | -0.015 | 0.025 | 0.560 | -1.09E-04 |
| DHA | 6,101 | AFR | RINT | OFI | 0.414 | 0.026 | 4.85E-56 | 0.040 | 0.052 | 0.021 | 0.014 | 0.001 | 0.003 | 0.025 | 0.914 | -1.63E-04 |
| DHA% | 6,091 | AFR | RINT | FOS | 0.166 | 0.025 | 4.18E-11 | 0.007 | 0.022 | 0.014 | 0.131 | 0.000 | -0.038 | 0.024 | 0.117 | 2.40E-04 |
| DHA% | 6,091 | AFR | RINT | OFI | 0.541 | 0.025 | 4.88E-98 | 0.070 | 0.027 | 0.020 | 0.180 | 0.000 | -0.028 | 0.025 | 0.258 | 4.57E-05 |
| Omega-3 | 6,115 | AFR | RINT | FOS | 0.142 | 0.026 | 3.41E-08 | 0.005 | 0.075 | 0.015 | 4.86E-07 | 0.004 | -0.019 | 0.025 | 0.461 | -7.49E-05 |
| Omega-3 | 6,115 | AFR | RINT | OFI | 0.361 | 0.026 | 2.22E-43 | 0.031 | 0.074 | 0.021 | 0.000 | 0.002 | -0.008 | 0.026 | 0.752 | -1.48E-04 |
| Omega-3% | 6,072 | AFR | RINT | FOS | 0.165 | 0.025 | 6.90E-11 | 0.007 | 0.022 | 0.014 | 0.120 | 0.000 | -0.022 | 0.025 | 0.375 | -3.53E-05 |
| Omega-3% | 6,072 | AFR | RINT | OFI | 0.503 | 0.025 | 5.20E-85 | 0.061 | 0.011 | 0.020 | 0.564 | 0.000 | 0.005 | 0.025 | 0.828 | -1.58E-04 |
| DHA | 7,891 | CSA | RINT | FOS | 0.347 | 0.024 | 2.37E-45 | 0.025 | 0.144 | 0.012 | 5.52E-35 | 0.019 | -0.022 | 0.024 | 0.357 | -1.92E-05 |
| DHA | 7,891 | CSA | RINT | OFI | 0.495 | 0.021 | 2.81E-116 | 0.064 | 0.150 | 0.013 | 1.11E-29 | 0.016 | -0.026 | 0.020 | 0.205 | 7.68E-05 |
| DHA% | 7,908 | CSA | RINT | FOS | 0.352 | 0.024 | 1.13E-48 | 0.027 | 0.140 | 0.012 | 2.78E-32 | 0.017 | -0.056 | 0.024 | 0.017 | 5.95E-04 |
| DHA% | 7,908 | CSA | RINT | OFI | 0.485 | 0.021 | 1.80E-115 | 0.064 | 0.142 | 0.013 | 5.21E-26 | 0.014 | -0.035 | 0.020 | 0.079 | 2.64E-04 |
| Omega-3 | 7,964 | CSA | RINT | FOS | 0.313 | 0.024 | 2.98E-37 | 0.020 | 0.198 | 0.012 | 5.03E-65 | 0.036 | -0.021 | 0.024 | 0.388 | -3.20E-05 |
| Omega-3 | 7,964 | CSA | RINT | OFI | 0.446 | 0.021 | 3.10E-94 | 0.052 | 0.215 | 0.013 | 1.73E-59 | 0.033 | -0.054 | 0.020 | 0.007 | 7.78E-04 |
| Omega-3% | 7,929 | CSA | RINT | FOS | 0.393 | 0.023 | 3.55E-63 | 0.035 | 0.173 | 0.012 | 6.19E-50 | 0.027 | -0.023 | 0.023 | 0.308 | 5.09E-06 |
| Omega-3% | 7,929 | CSA | RINT | OFI | 0.518 | 0.020 | 2.47E-138 | 0.076 | 0.190 | 0.013 | 3.12E-47 | 0.026 | -0.053 | 0.019 | 0.006 | 8.39E-04 |
| DHA | 2,461 | EAS | RINT | FOS | 0.235 | 0.040 | 5.36E-09 | 0.013 | 0.047 | 0.023 | 0.043 | 0.001 | -0.007 | 0.040 | 0.853 | -3.96E-04 |
| DHA | 2,461 | EAS | RINT | OFI | 0.504 | 0.039 | 8.19E-38 | 0.065 | 0.016 | 0.030 | 0.589 | 0.000 | 0.045 | 0.037 | 0.228 | 1.86E-04 |

Continued

| DHA% | 2,491 | EAS | RINT | FOS | 0.215 | 0.040 | 6.74E-08 | 0.011 | 0.034 | 0.024 | 0.154 | 0.000 | 0.003 | 0.039 | 0.940 | -4.03E-04 |
| --- | --- | --- | --- | --- | --- | --- | --- | --- | --- | --- | --- | --- | --- | --- | --- | --- |
| DHA% | 2,491 | EAS | RINT | OFI | 0.479 | 0.038 | 4.19E-35 | 0.060 | 0.004 | 0.031 | 0.905 | 0.000 | 0.050 | 0.037 | 0.181 | 3.20E-04 |
| Omega-3 | 2,467 | EAS | RINT | FOS | 0.241 | 0.041 | 6.52E-09 | 0.013 | 0.084 | 0.024 | 0.000 | 0.005 | 0.026 | 0.041 | 0.525 | -2.44E-04 |
| Omega-3 | 2,467 | EAS | RINT | OFI | 0.447 | 0.040 | 9.97E-29 | 0.049 | 0.047 | 0.031 | 0.135 | 0.001 | 0.073 | 0.038 | 0.057 | 1.07E-03 |
| Omega-3% | 2,477 | EAS | RINT | FOS | 0.255 | 0.041 | 5.73E-10 | 0.015 | 0.050 | 0.025 | 0.042 | 0.001 | -0.020 | 0.040 | 0.622 | -3.08E-04 |
| Omega-3% | 2,477 | EAS | RINT | OFI | 0.517 | 0.039 | 3.70E-38 | 0.065 | 0.029 | 0.032 | 0.375 | 0.000 | 0.024 | 0.038 | 0.537 | -2.53E-04 |

Linear regression model including the PGS-by-FOS interaction term was applied in each ancestry group. When FOS is defined as the dietary exposure variable, all reported statistics, including the coefficient (Beta), standard error (SE), p-value, variance explained (R2), and interaction terms, pertain specifically to the effects of FOS. Conversely, when OFI is used as dietary exposure, these statistical parameters apply to OFI. Abbreviations: Omega-3, omega-3 fatty acids; Omega-3%, omega-3 fatty acids to total fatty acids percentage; DHA, docosahexaenoic acid; DHA%, docosahexaenoic acid to total fatty acids percentage; FOS, fish oil supplementation; OFI, oily fish intake; PGS, polygenic score; SE, standard error; RINT, rank-based inverse normal transformation; AFR, African; CSA, Central/South Asian; EAS, East Asian.

### **Supplementary Table 12.** Association of dietary exposures and the observed RINT-based circulating concentrations of omega-3 fatty acids stratified by PGS groups in CSA participants

| **Trait** | **Group** | **Dietary exposure** | **Beta (Dietary exposure)** | **SE (Dietary exposure)** | **P (Dietary exposure)** | **R2 (Dietary exposure)** |
| --- | --- | --- | --- | --- | --- | --- |
| DHA | all PGS | OFI | 0.495 | 0.021 | 3.52E-116 | 0.067 |
| DHA | 0-5% PGS | OFI | 0.746 | 0.093 | 1.78E-14 | 0.143 |
| DHA | 95-100% PGS | OFI | 0.476 | 0.099 | 2.22E-06 | 0.058 |
| DHA | 0-20% PGS | OFI | 0.542 | 0.049 | 5.60E-27 | 0.070 |
| DHA | 20-40% PGS | OFI | 0.474 | 0.049 | 1.04E-21 | 0.059 |
| DHA | 40-60% PGS | OFI | 0.448 | 0.047 | 4.10E-21 | 0.057 |
| DHA | 60-80% PGS | OFI | 0.508 | 0.047 | 1.80E-26 | 0.076 |
| DHA | 80-100% PGS | OFI | 0.501 | 0.047 | 1.32E-25 | 0.073 |
| DHA% | all PGS | OFI | 0.485 | 0.021 | 1.62E-115 | 0.065 |
| DHA% | 0-5% PGS | OFI | 0.554 | 0.099 | 4.65E-08 | 0.062 |
| DHA% | 95-100% PGS | OFI | 0.377 | 0.093 | 6.12E-05 | 0.046 |
| DHA% | 0-20% PGS | OFI | 0.506 | 0.049 | 7.22E-24 | 0.060 |
| DHA% | 20-40% PGS | OFI | 0.474 | 0.048 | 1.90E-22 | 0.066 |
| DHA% | 40-60% PGS | OFI | 0.521 | 0.048 | 4.83E-27 | 0.069 |
| DHA% | 60-80% PGS | OFI | 0.492 | 0.045 | 1.49E-26 | 0.071 |
| DHA% | 80-100% PGS | OFI | 0.415 | 0.044 | 2.25E-20 | 0.059 |
| Omega-3 | all PGS | OFI | 0.445 | 0.021 | 4.66E-94 | 0.054 |
| Omega-3 | 0-5% PGS | OFI | 0.555 | 0.103 | 1.12E-07 | 0.072 |
| Omega-3 | 95-100% PGS | OFI | 0.332 | 0.100 | 9.28E-04 | 0.034 |
| Omega-3 | 0-20% PGS | OFI | 0.515 | 0.050 | 3.91E-24 | 0.063 |
| Omega-3 | 20-40% PGS | OFI | 0.423 | 0.050 | 5.98E-17 | 0.046 |
| Omega-3 | 40-60% PGS | OFI | 0.494 | 0.046 | 1.54E-25 | 0.074 |
| Omega-3 | 60-80% PGS | OFI | 0.445 | 0.048 | 4.27E-20 | 0.054 |
| Omega-3 | 80-100% PGS | OFI | 0.338 | 0.045 | 1.43E-13 | 0.033 |
| Omega-3% | all PGS | OFI | 0.518 | 0.020 | 3.05E-138 | 0.077 |

Continued

| Omega-3% | 0-5% PGS | OFI | 0.657 | 0.100 | 1.57E-10 | 0.104 |
| --- | --- | --- | --- | --- | --- | --- |
| Omega-3% | 95-100% PGS | OFI | 0.455 | 0.088 | 4.06E-07 | 0.073 |
| Omega-3% | 0-20% PGS | OFI | 0.545 | 0.048 | 5.92E-29 | 0.077 |
| Omega-3% | 20-40% PGS | OFI | 0.584 | 0.047 | 2.28E-34 | 0.095 |
| Omega-3% | 40-60% PGS | OFI | 0.517 | 0.045 | 2.30E-29 | 0.079 |
| Omega-3% | 60-80% PGS | OFI | 0.492 | 0.046 | 4.21E-26 | 0.065 |
| Omega-3% | 80-100% PGS | OFI | 0.434 | 0.042 | 4.01E-24 | 0.066 |
| DHA | all PGS | FOS | 0.347 | 0.024 | 2.39E-45 | 0.025 |
| DHA | 0-5% PGS | FOS | 0.390 | 0.109 | 3.94E-04 | 0.030 |
| DHA | 95-100% PGS | FOS | 0.276 | 0.113 | 1.48E-02 | 0.013 |
| DHA | 0-20% PGS | FOS | 0.360 | 0.056 | 1.31E-10 | 0.026 |
| DHA | 20-40% PGS | FOS | 0.316 | 0.055 | 1.43E-08 | 0.020 |
| DHA | 40-60% PGS | FOS | 0.392 | 0.054 | 6.45E-13 | 0.032 |
| DHA | 60-80% PGS | FOS | 0.347 | 0.054 | 1.56E-10 | 0.025 |
| DHA | 80-100% PGS | FOS | 0.313 | 0.055 | 1.36E-08 | 0.020 |
| DHA% | all PGS | FOS | 0.353 | 0.024 | 6.86E-49 | 0.027 |
| DHA% | 0-5% PGS | FOS | 0.326 | 0.118 | 5.86E-03 | 0.018 |
| DHA% | 95-100% PGS | FOS | 0.317 | 0.108 | 3.46E-03 | 0.020 |
| DHA% | 0-20% PGS | FOS | 0.403 | 0.056 | 8.05E-13 | 0.032 |
| DHA% | 20-40% PGS | FOS | 0.446 | 0.055 | 1.60E-15 | 0.039 |
| DHA% | 40-60% PGS | FOS | 0.288 | 0.054 | 1.29E-07 | 0.017 |
| DHA% | 60-80% PGS | FOS | 0.315 | 0.051 | 8.05E-10 | 0.023 |
| DHA% | 80-100% PGS | FOS | 0.302 | 0.051 | 5.43E-09 | 0.021 |
| Omega-3 | all PGS | FOS | 0.313 | 0.024 | 3.26E-37 | 0.020 |
| Omega-3 | 0-5% PGS | FOS | 0.303 | 0.118 | 1.06E-02 | 0.015 |
| Omega-3 | 95-100% PGS | FOS | 0.310 | 0.107 | 4.11E-03 | 0.019 |
| Omega-3 | 0-20% PGS | FOS | 0.314 | 0.057 | 5.44E-08 | 0.018 |
| Omega-3 | 20-40% PGS | FOS | 0.294 | 0.056 | 2.09E-07 | 0.016 |

Continued

| Omega-3 | 40-60% PGS | FOS | 0.359 | 0.054 | 3.07E-11 | 0.027 |
| --- | --- | --- | --- | --- | --- | --- |
| Omega-3 | 60-80% PGS | FOS | 0.281 | 0.054 | 2.24E-07 | 0.016 |
| Omega-3 | 80-100% PGS | FOS | 0.322 | 0.052 | 1.07E-09 | 0.023 |
| Omega-3% | all PGS | FOS | 0.393 | 0.023 | 2.89E-63 | 0.035 |
| Omega-3% | 0-5% PGS | FOS | 0.527 | 0.117 | 8.58E-06 | 0.049 |
| Omega-3% | 95-100% PGS | FOS | 0.219 | 0.103 | 3.48E-02 | 0.009 |
| Omega-3% | 0-20% PGS | FOS | 0.396 | 0.054 | 2.28E-13 | 0.033 |
| Omega-3% | 20-40% PGS | FOS | 0.371 | 0.054 | 6.76E-12 | 0.029 |
| Omega-3% | 40-60% PGS | FOS | 0.385 | 0.052 | 1.36E-13 | 0.034 |
| Omega-3% | 60-80% PGS | FOS | 0.476 | 0.052 | 2.71E-19 | 0.049 |
| Omega-3% | 80-100% PGS | FOS | 0.349 | 0.049 | 1.37E-12 | 0.031 |

The PGS were stratified into seven groups based on their distribution: the bottom 5% (0–5%), five quintile-based bins (0–20%, 20–40%, 40–60%, 60–80%, and 80–100%), and the top 5% (95–100%). Linear regression model excluding the PGS-by-diet interaction term was applied in each PGS group. When FOS is defined as the dietary exposure variable, all reported statistics, including the coefficient (Beta), standard error (SE), p-value, variance explained (R2), and interaction terms, pertain specifically to the effects of FOS. Conversely, when OFI is used as dietary exposure, these statistical parameters apply to OFI. Abbreviations: Omega-3, omega-3 fatty acids; Omega-3%, omega-3 fatty acids to total fatty acids percentage; DHA, docosahexaenoic acid; DHA%, docosahexaenoic acid to total fatty acids percentage; FOS, fish oil supplementation; OFI, oily fish intake; PGS, polygenic score; SE, standard error; RINT, rank-based inverse normal transformation; CSA, Central/South Asian.

### **Supplementary Table 13.** Association of PGS with the observed circulating concentrations of omega-3 fatty acids stratified by dietary status in CSA participants

| **Trait** | **Sample Size** | **Data Type** | **Dietary group** | **Beta (PGS)** | **SE (PGS)** | **P (PGS)** | **R2 (PGS)** |
| --- | --- | --- | --- | --- | --- | --- | --- |
| DHA | 1,713 | RINT | FOS | 0.1136 | 0.0227 | 6.38E-07 | 0.0140 |
| DHA | 6,178 | RINT | non-FOS | 0.1448 | 0.0117 | 1.27E-34 | 0.0240 |
| DHA% | 1,733 | RINT | FOS | 0.0637 | 0.0236 | 6.95E-03 | 0.0037 |
| DHA% | 6,175 | RINT | non-FOS | 0.1460 | 0.0119 | 3.61E-34 | 0.0237 |
| Omega-3 | 1,753 | RINT | FOS | 0.1704 | 0.0222 | 2.50E-14 | 0.0325 |
| Omega-3 | 6,211 | RINT | non-FOS | 0.1978 | 0.0115 | 1.81E-64 | 0.0452 |
| Omega-3% | 1,745 | RINT | FOS | 0.1350 | 0.0227 | 3.13E-09 | 0.0196 |
| Omega-3% | 6,184 | RINT | non-FOS | 0.1765 | 0.0117 | 1.89E-50 | 0.0354 |
| DHA | 1,713 | Raw | FOS | 0.0074 | 0.0015 | 7.53E-07 | 0.0138 |
| DHA | 6,178 | Raw | non-FOS | 0.0089 | 0.0007 | 1.83E-34 | 0.0239 |
| DHA% | 1,733 | Raw | FOS | 0.0320 | 0.0128 | 1.24E-02 | 0.0031 |
| DHA% | 6,175 | Raw | non-FOS | 0.0750 | 0.0062 | 1.23E-33 | 0.0233 |
| Omega-3 | 1,753 | Raw | FOS | 0.0314 | 0.0042 | 1.09E-13 | 0.0308 |
| Omega-3 | 6,211 | Raw | non-FOS | 0.0338 | 0.0020 | 2.84E-60 | 0.0423 |
| Omega-3% | 1,745 | Raw | FOS | 0.1662 | 0.0286 | 7.80E-09 | 0.0186 |
| Omega-3% | 6,184 | Raw | non-FOS | 0.2041 | 0.0141 | 5.57E-47 | 0.0329 |
| DHA | 3,093 | RINT | high-OFI | 0.1311 | 0.0172 | 2.87E-14 | 0.0183 |
| DHA | 4,798 | RINT | low-OFI | 0.1432 | 0.0131 | 1.11E-27 | 0.0244 |
| DHA% | 3,111 | RINT | high-OFI | 0.1124 | 0.0174 | 1.19E-10 | 0.0130 |
| DHA% | 4,797 | RINT | low-OFI | 0.1407 | 0.0133 | 1.07E-25 | 0.0225 |
| Omega-3 | 3,162 | RINT | high-OFI | 0.1682 | 0.0162 | 5.33E-25 | 0.0331 |
| Omega-3 | 4,802 | RINT | low-OFI | 0.2088 | 0.0132 | 3.27E-55 | 0.0498 |
| Omega-3% | 3,122 | RINT | high-OFI | 0.1469 | 0.0167 | 2.70E-18 | 0.0239 |
| Omega-3% | 4,807 | RINT | low-OFI | 0.1839 | 0.0132 | 2.90E-43 | 0.0388 |
| DHA | 3,093 | Raw | high-OFI | 0.0084 | 0.0011 | 4.88E-14 | 0.0180 |
| DHA | 4,798 | Raw | low-OFI | 0.0086 | 0.0008 | 6.26E-28 | 0.0246 |

Continued

| DHA% | 3,111 | Raw | high-OFI | 0.0598 | 0.0094 | 2.03E-10 | 0.0127 |
| --- | --- | --- | --- | --- | --- | --- | --- |
| DHA% | 4,797 | Raw | low-OFI | 0.0707 | 0.0068 | 7.60E-25 | 0.0217 |
| Omega-3 | 3,162 | Raw | high-OFI | 0.0306 | 0.0031 | 6.40E-23 | 0.0302 |
| Omega-3 | 4,802 | Raw | low-OFI | 0.0353 | 0.0023 | 1.59E-53 | 0.0482 |
| Omega-3% | 3,122 | Raw | high-OFI | 0.1776 | 0.0213 | 1.02E-16 | 0.0217 |
| Omega-3% | 4,807 | Raw | low-OFI | 0.2120 | 0.0155 | 7.83E-42 | 0.0375 |

FOS or OFI were defined as dietary exposure, respectively. Linear regression model excluding the PGS-by-diet interaction term was applied stratified by dietary status. Effect sizes (Beta and SE) and p-values testing the association between PGS and circulating concentrations of omega-3 fatty acids are reported. Abbreviations: Omega-3, omega-3 fatty acids; Omega-3%, omega-3 fatty acids to total fatty acids percentage; DHA, docosahexaenoic acid; DHA%, docosahexaenoic acid to total fatty acids percentage; FOS, fish oil supplementation; OFI, oily fish intake; PGS, polygenic score; SE, standard error; RINT, rank-based inverse normal transformation; CSA, Central/South Asian.

### **Supplementary Table 14.** Association of FOS, PGS and PGS-by-FOS with the observed circulating concentrations of omega-3 fatty acids in EUR participants (sensitivity analysis)

| **Trait** | **DHA** | **DHA%** | **Omega-3** | **Omega-3%** |
| --- | --- | --- | --- | --- |
| Setting: European (Phase 2), P+T-derived PGS, raw phenotype, touchscreen questionnaire | | | | |
| Sample Size | 230,691 | 231,052 | 231,243 | 230,657 |
| Beta (FOS) | 0.0267 | 0.2081 | 0.0681 | 0.5386 |
| SE (FOS) | 0.0003 | 0.0022 | 0.0008 | 0.0050 |
| P (FOS) | 1.00E-300 | 1.00E-300 | 1.00E-300 | 1.00E-300 |
| R2 (FOS) | 0.0404 | 0.0374 | 0.0342 | 0.0483 |
| Beta (PGS) | 0.0156 | 0.1004 | 0.0496 | 0.3198 |
| SE (PGS) | 0.0001 | 0.0012 | 0.0004 | 0.0027 |
| P (PGS) | 1.00E-300 | 1.00E-300 | 1.00E-300 | 1.00E-300 |
| R2 (PGS) | 0.0461 | 0.0294 | 0.0592 | 0.0567 |
| Beta (PGS-by-FOS) | -0.0005 | -0.0064 | -0.0029 | -0.0245 |
| SE (PGS-by-FOS) | 0.0003 | 0.0022 | 0.0007 | 0.0049 |
| P (PGS-by-FOS) | 4.55E-02 | 2.82E-03 | 9.37E-05 | 5.21E-07 |
| R2 (PGS-by-FOS) | 1.30E-05 | 3.43E-05 | 6.17E-05 | 1.05E-04 |
| Setting: European (Phase 2), P+T-derived PGS, RINT-based phenotype, touchscreen questionnaire | | | | |
| Sample Size | 230,691 | 231,052 | 231,243 | 230,657 |
| Beta (FOS) | 0.3765 | 0.3607 | 0.3588 | 0.4153 |
| SE (FOS) | 0.0039 | 0.0038 | 0.0040 | 0.0039 |
| P (FOS) | 1.00E-300 | 1.00E-300 | 1.00E-300 | 1.00E-300 |
| R2 (FOS) | 0.0395 | 0.0366 | 0.0340 | 0.0478 |
| Beta (PGS) | 0.2347 | 0.1847 | 0.2796 | 0.2667 |
| SE (PGS) | 0.0021 | 0.0021 | 0.0022 | 0.0021 |
| P (PGS) | 1.00E-300 | 1.00E-300 | 1.00E-300 | 1.00E-300 |
| R2 (PGS) | 0.0509 | 0.0323 | 0.0668 | 0.0650 |
| Beta (PGS-by-FOS) | -0.0239 | -0.0198 | -0.0384 | -0.0395 |

Continued

| SE (PGS-by-FOS) | 0.0038 | 0.0038 | 0.0039 | 0.0038 |
| --- | --- | --- | --- | --- |
| P (PGS-by-FOS) | 2.63E-10 | 1.41E-07 | 5.50E-23 | 1.30E-25 |
| R2 (PGS-by-FOS) | 0.00017 | 0.00012 | 0.00042 | 0.00047 |
| Setting: European (all phases), SBayesRC-derived PGS, raw phenotype, 24-hour dietary recall questionnaire | | | | |
| Sample Size | 175,291 | 175,512 | 175,562 | 175,035 |
| Beta (FOS) | 0.0248 | 0.1945 | 0.0618 | 0.4871 |
| SE (FOS) | 0.0003 | 0.0028 | 0.0009 | 0.0062 |
| P (FOS) | 1.00E-300 | 1.00E-300 | 1.00E-300 | 1.00E-300 |
| R2 (FOS) | 0.0291 | 0.0273 | 0.0252 | 0.0340 |
| Beta (PGS) | 0.0177 | 0.1151 | 0.0572 | 0.3534 |
| SE (PGS) | 0.0002 | 0.0015 | 0.0005 | 0.0033 |
| P (PGS) | 1.00E-300 | 1.00E-300 | 1.00E-300 | 1.00E-300 |
| R2 (PGS) | 0.0518 | 0.0341 | 0.0736 | 0.0625 |
| Beta (PGS-by-FOS) | -0.0007 | -0.0088 | -0.0024 | -0.0303 |
| SE (PGS-by-FOS) | 0.0003 | 0.0027 | 0.0009 | 0.0061 |
| P (PGS-by-FOS) | 2.63E-02 | 1.26E-03 | 0.00815238 | 6.64E-07 |
| R2 (PGS-by-FOS) | 2.25E-05 | 5.36E-05 | 3.42E-05 | 1.36E-04 |
| Setting: European (all phases), SBayesRC-derived PGS, RINT-based phenotype, 24-hour dietary recall questionnaire | | | | |
| Sample Size | 175,291 | 175,512 | 175,562 | 175,035 |
| Beta (FOS) | 0.3418 | 0.3330 | 0.3221 | 0.3754 |
| SE (FOS) | 0.0048 | 0.0048 | 0.0048 | 0.0048 |
| P (FOS) | 1.00E-300 | 1.00E-300 | 1.00E-300 | 1.00E-300 |
| R2 (FOS) | 0.0286 | 0.0271 | 0.0252 | 0.0339 |
| Beta (PGS) | 0.2552 | 0.2066 | 0.3133 | 0.2924 |
| SE (PGS) | 0.0025 | 0.0025 | 0.0025 | 0.0025 |
| P (PGS) | 1.00E-300 | 1.00E-300 | 1.00E-300 | 1.00E-300 |
| R2 (PGS) | 0.0557 | 0.0370 | 0.0808 | 0.0711 |
| Beta (PGS-by-FOS) | -0.0247 | -0.0235 | -0.0330 | -0.0423 |

Continued

| SE (PGS-by-FOS) | 0.0047 | 0.0047 | 0.0047 | 0.0047 |
| --- | --- | --- | --- | --- |
| P (PGS-by-FOS) | 1.27E-07 | 5.49E-07 | 2.36E-12 | 2.65E-19 |
| R2 (PGS-by-FOS) | 0.0002 | 0.0001 | 0.0003 | 0.0005 |

Linear regression model including the PGS-by-FOS interaction term was applied. Effect sizes (Beta and SE) and p-values testing the association between PGS-by-FOS and circulating concentrations of omega-3 fatty acids are reported. Abbreviations: Omega-3, omega-3 fatty acids; Omega-3%, omega-3 fatty acids to total fatty acids percentage; DHA, docosahexaenoic acid; DHA%, docosahexaenoic acid to total fatty acids percentage; FOS, fish oil supplementation; PGS, polygenic score; SE, standard error; RINT, rank-based inverse normal transformation.

### **Supplementary Table 15.** Association of FOS and the observed RINT-based circulating concentrations of omega-3 fatty acids stratified by PGS groups in EUR participants (all phases) using a 24-hour dietary recall questionnaire (sensitivity analysis)

| **Trait** | **Group** | **Beta (FOS)** | **SE (FOS)** | **P (FOS)** | **R2 (FOS)** |
| --- | --- | --- | --- | --- | --- |
| DHA | all PGS | 0.3409 | 0.0049 | 1.00E-300 | 0.0264 |
| DHA | 0-5% PGS | 0.3542 | 0.0225 | 2.65E-55 | 0.0272 |
| DHA | 95-100% PGS | 0.2638 | 0.0236 | 1.03E-28 | 0.0186 |
| DHA | 0-20% PGS | 0.3819 | 0.0110 | 7.33E-260 | 0.0328 |
| DHA | 20-40% PGS | 0.3442 | 0.0106 | 8.46E-227 | 0.0287 |
| DHA | 40-60% PGS | 0.3423 | 0.0106 | 6.53E-227 | 0.0287 |
| DHA | 60-80% PGS | 0.3324 | 0.0104 | 4.10E-219 | 0.0277 |
| DHA | 80-100% PGS | 0.3091 | 0.0107 | 5.41E-182 | 0.0246 |
| DHA% | all PGS | 0.3340 | 0.0049 | 1.00E-300 | 0.0259 |
| DHA% | 0-5% PGS | 0.3642 | 0.0234 | 9.73E-54 | 0.0264 |
| DHA% | 95-100% PGS | 0.2891 | 0.0231 | 1.55E-35 | 0.0232 |
| DHA% | 0-20% PGS | 0.3570 | 0.0114 | 9.71E-213 | 0.0269 |
| DHA% | 20-40% PGS | 0.3599 | 0.0108 | 6.97E-241 | 0.0304 |
| DHA% | 40-60% PGS | 0.3193 | 0.0105 | 8.16E-200 | 0.0252 |
| DHA% | 60-80% PGS | 0.3257 | 0.0103 | 2.17E-218 | 0.0276 |
| DHA% | 80-100% PGS | 0.2982 | 0.0104 | 2.76E-180 | 0.0243 |
| Omega-3 | all PGS | 0.3224 | 0.0051 | 1.00E-300 | 0.0227 |
| Omega-3 | 0-5% PGS | 0.3740 | 0.0230 | 9.36E-59 | 0.0289 |
| Omega-3 | 95-100% PGS | 0.2439 | 0.0238 | 1.77E-24 | 0.0156 |
| Omega-3 | 0-20% PGS | 0.3672 | 0.0112 | 8.96E-231 | 0.0291 |
| Omega-3 | 20-40% PGS | 0.3340 | 0.0108 | 8.30E-209 | 0.0264 |
| Omega-3 | 40-60% PGS | 0.3169 | 0.0105 | 1.24E-197 | 0.0249 |
| Omega-3 | 60-80% PGS | 0.3163 | 0.0105 | 2.41E-196 | 0.0248 |
| Omega-3 | 80-100% PGS | 0.2776 | 0.0107 | 9.89E-146 | 0.0197 |
| Omega-3% | all PGS | 0.3781 | 0.0050 | 1.00E-300 | 0.0313 |
| Omega-3% | 0-5% PGS | 0.4520 | 0.0246 | 5.50E-74 | 0.0366 |

Continued

| Omega-3% | 95-100% PGS | 0.3038 | 0.0227 | 2.92E-40 | 0.0265 |
| --- | --- | --- | --- | --- | --- |
| Omega-3% | 0-20% PGS | 0.4202 | 0.0119 | 1.80E-269 | 0.0341 |
| Omega-3% | 20-40% PGS | 0.4077 | 0.0109 | 3.27E-300 | 0.0379 |
| Omega-3% | 40-60% PGS | 0.3776 | 0.0105 | 2.03E-280 | 0.0354 |
| Omega-3% | 60-80% PGS | 0.3414 | 0.0102 | 1.22E-242 | 0.0307 |
| Omega-3% | 80-100% PGS | 0.3258 | 0.0102 | 1.09E-221 | 0.0300 |

The PGS were stratified into seven groups based on their distribution: the bottom 5% (0–5%), five quintile-based bins (0–20%, 20–40%, 40–60%, 60–80%, and 80–100%), and the top 5% (95–100%). Linear regression model excluding the PGS-by-FOS interaction term was applied in each PGS group. Effect sizes (Beta and SE) and p-values testing the association between FOS and circulating concentration of omega-3 fatty acids are reported. Abbreviations: Omega-3, omega-3 fatty acids; Omega-3%, omega-3 fatty acids to total fatty acids percentage; DHA, docosahexaenoic acid; DHA%, docosahexaenoic acid to total fatty acids percentage; FOS, fish oil supplementation; PGS, polygenic score; SE, standard error; RINT, rank-based inverse normal transformation.

### **Supplementary Table 16.** Association of PGS with the observed circulating concentrations of omega-3 fatty acids stratified by FOS status in EUR participants (sensitivity analysis)

| **Trait** | **Sample Size** | **Population** | **PGS approach** | **Data Type** | **Questionnaire** | **Dietary group** | **Beta (PGS)** | **SE (PGS)** | **P (PGS)** | **R2 (PGS)** |
| --- | --- | --- | --- | --- | --- | --- | --- | --- | --- | --- |
| DHA | 50,689 | European (all phases) | SBayesRC | Raw | 24-hour dietary recall | FOS | 0.0170 | 0.0003 | 1.00E-300 | 0.1848 |
| DHA | 124,602 | European (all phases) | SBayesRC | Raw | 24-hour dietary recall | non-FOS | 0.0176 | 0.0002 | 1.00E-300 | 0.1761 |
| DHA | 50,689 | European (all phases) | SBayesRC | RINT | 24-hour dietary recall | FOS | 0.2316 | 0.0039 | 1.00E-300 | 0.1878 |
| DHA | 124,602 | European (all phases) | SBayesRC | RINT | 24-hour dietary recall | non-FOS | 0.2547 | 0.0025 | 1.00E-300 | 0.1813 |
| DHA% | 50,938 | European (all phases) | SBayesRC | Raw | 24-hour dietary recall | FOS | 0.1060 | 0.0023 | 1.00E-300 | 0.1724 |
| DHA% | 124,574 | European (all phases) | SBayesRC | Raw | 24-hour dietary recall | non-FOS | 0.1152 | 0.0014 | 1.00E-300 | 0.1778 |
| DHA% | 50,938 | European (all phases) | SBayesRC | RINT | 24-hour dietary recall | FOS | 0.1829 | 0.0040 | 1.00E-300 | 0.1764 |
| DHA% | 124,574 | European (all phases) | SBayesRC | RINT | 24-hour dietary recall | non-FOS | 0.2067 | 0.0025 | 1.00E-300 | 0.1842 |
| Omega-3 | 50,746 | European (all phases) | SBayesRC | Raw | 24-hour dietary recall | FOS | 0.0551 | 0.0008 | 1.00E-300 | 0.1685 |
| Omega-3 | 124,816 | European (all phases) | SBayesRC | Raw | 24-hour dietary recall | non-FOS | 0.0570 | 0.0005 | 1.00E-300 | 0.1712 |
| Omega-3 | 50,746 | European (all phases) | SBayesRC | RINT | 24-hour dietary recall | FOS | 0.2815 | 0.0039 | 1.00E-300 | 0.1725 |
| Omega-3 | 124,816 | European (all phases) | SBayesRC | RINT | 24-hour dietary recall | non-FOS | 0.3127 | 0.0025 | 1.00E-300 | 0.1780 |
| Omega-3% | 50,537 | European (all phases) | SBayesRC | Raw | 24-hour dietary recall | FOS | 0.3225 | 0.0052 | 1.00E-300 | 0.1586 |
| Omega-3% | 124,498 | European (all phases) | SBayesRC | Raw | 24-hour dietary recall | non-FOS | 0.3536 | 0.0032 | 1.00E-300 | 0.1617 |
| Omega-3% | 50,537 | European (all phases) | SBayesRC | RINT | 24-hour dietary recall | FOS | 0.2496 | 0.0039 | 1.00E-300 | 0.1635 |
| Omega-3% | 124,498 | European (all phases) | SBayesRC | RINT | 24-hour dietary recall | non-FOS | 0.2925 | 0.0025 | 1.00E-300 | 0.1699 |
| DHA | 72,276 | European (Phase 2) | P+T | Raw | Touchscreen | FOS | 0.0150 | 0.0002 | 1.00E-300 | 0.0522 |
| DHA | 159,516 | European (Phase 2) | P+T | Raw | Touchscreen | non-FOS | 0.0156 | 0.0002 | 1.00E-300 | 0.0621 |
| DHA | 72,276 | European (Phase 2) | P+T | RINT | Touchscreen | FOS | 0.2099 | 0.0033 | 1.00E-300 | 0.0541 |
| DHA | 159,516 | European (Phase 2) | P+T | RINT | Touchscreen | non-FOS | 0.2344 | 0.0022 | 1.00E-300 | 0.0661 |
| DHA% | 72,651 | European (Phase 2) | P+T | Raw | Touchscreen | FOS | 0.0930 | 0.0019 | 1.00E-300 | 0.0310 |
| DHA% | 159,501 | European (Phase 2) | P+T | Raw | Touchscreen | non-FOS | 0.1004 | 0.0012 | 1.00E-300 | 0.0393 |
| DHA% | 72,651 | European (Phase 2) | P+T | RINT | Touchscreen | FOS | 0.1632 | 0.0033 | 1.00E-300 | 0.0328 |
| DHA% | 159,501 | European (Phase 2) | P+T | RINT | Touchscreen | non-FOS | 0.1848 | 0.0022 | 1.00E-300 | 0.0423 |
| Omega-3 | 72,565 | European (Phase 2) | P+T | Raw | Touchscreen | FOS | 0.0467 | 0.0006 | 1.00E-300 | 0.0666 |

Continued

| Omega-3 | 159,780 | European (Phase 2) | P+T | Raw | Touchscreen | non-FOS | 0.0495 | 0.0004 | 1.00E-300 | 0.0811 |
| --- | --- | --- | --- | --- | --- | --- | --- | --- | --- | --- |
| Omega-3 | 72,565 | European (Phase 2) | P+T | RINT | Touchscreen | FOS | 0.2410 | 0.0033 | 1.00E-300 | 0.0697 |
| Omega-3 | 159,780 | European (Phase 2) | P+T | RINT | Touchscreen | non-FOS | 0.2792 | 0.0023 | 1.00E-300 | 0.0874 |
| Omega-3% | 72,221 | European (Phase 2) | P+T | Raw | Touchscreen | FOS | 0.2919 | 0.0043 | 1.00E-300 | 0.0589 |
| Omega-3% | 159,536 | European (Phase 2) | P+T | Raw | Touchscreen | non-FOS | 0.3191 | 0.0028 | 1.00E-300 | 0.0742 |
| Omega-3% | 72,221 | European (Phase 2) | P+T | RINT | Touchscreen | FOS | 0.2246 | 0.0032 | 1.00E-300 | 0.0628 |
| Omega-3% | 159,536 | European (Phase 2) | P+T | RINT | Touchscreen | non-FOS | 0.2661 | 0.0022 | 1.00E-300 | 0.0818 |

Linear regression model excluding the PGS-by-FOS interaction term was applied stratified by FOS status (yes vs no). Effect sizes (Beta and SE) and p-values testing the association between PGS and circulating concentrations of omega-3 fatty acids are reported. Abbreviations: Omega-3, omega-3 fatty acids; Omega-3%, omega-3 fatty acids to total fatty acids percentage; DHA, docosahexaenoic acid; DHA%, docosahexaenoic acid to total fatty acids percentage; FOS, fish oil supplementation; PGS, polygenic score; SE, standard error; RINT, rank-based inverse normal transformation.
